# Validation of an AI for Skin Diseases in Korea and Global Usage Statistics

**DOI:** 10.1101/2025.01.27.25321219

**Authors:** Seung Seog Han, Soo Ick Cho, Sung Eun Chang, Seong Hwan Kim, Jung-Im Na

**Affiliations:** Department of Dermatology, I Dermatology Clinic, Seoul, Korea; IDerma Inc., Seoul, Korea; Lunit Inc., Seoul, Korea; Department of Dermatology, Asan Medical Center, Ulsan University College of Medicine, Seoul, Korea; Department of Plastic and Reconstructive Surgery, Kangnam Sacred Hospital, Hallym University College of Medicine, Seoul, Korea; Department of Dermatology, Seoul National University College of Medicine, Seoul National University Bundang Hospital, Seoul, Korea

## Abstract

**Background:** To address the diversity of skin conditions and the low prevalence of skin cancers, we curated a large dataset and collected real-world data, to evaluate the real-world performance.

**Methods:** We evaluated a neural network model using the NIA dataset (70 diseases; 152,443 images from 92,004 cases from 9 hospitals). The NIA dataset was used to calculate the algorithm’s sensitivity for detecting skin cancer. Global usage statistics (1,690,849 queries) were analyzed to estimate the algorithm’s specificity for malignancy predictions, assuming all predictions were false positives.

A global reader test (61,066 questionnaires) was performed using the SNU dataset (80 diseases; 240 images).

**Results:** For malignancy diagnosis (NIA dataset), the sensitivity of the algorithm was 78.2% based on the three predicted differentials. In global usage statistics, among predictions for tumorous conditions, the malignancy prediction rates from the three differentials were 12.0% in Korea and 10.0% globally. The algorithm’s predictions for benign tumors were most prevalent in Asia (55.5%), Oceania (46.8%), and Europe (46.5%), while predictions for infectious diseases were more common in Africa (17.1%)

In the reader test (SNU dataset), sensitivity / specificity of the algorithm (87.5% / 91.0%) outperformed those of participants (Korea = 56.9% / 87.2%, global = 55.5% / 84.3%).

**Conclusion:** For skin cancer diagnosis in Korea, the sensitivity and specificity of the algorithm were estimated to be 78.2% and 88.0%. Further research on appropriate indications is required for screening use, taking overdiagnosis into consideration.

## INTRODUCTION

Artificial intelligence (AI) has demonstrated remarkable performance in dermatology, often surpassing dermatologists in certain controlled conditions. Those studies usually encompassing skin cancer and utilizing clinical photographs and dermoscopy images, have shown that AI can outperform dermatologists when diagnoses are based solely on image data. ^1-4^ Validation of various AI model performance has been made possible by the release of a variety of publicly available dermatology datasets. ^5-9^

Unlike radiology, where x-ray diagnoses rely entirely on image data, dermatological diagnosis in clinical practice incorporates various contextual and patient-specific information beyond images. Consequently, while AI may outperform dermatologists in purely image-based diagnosis, clinicians in real practice are much more accurate. ^10^ Historical examples, such as the MelaFind algorithm, which received FDA approval but ultimately disappeared from the market due to high false-positive rates, highlight how algorithms that perform well under controlled conditions can struggle in real-world applications, where they may exhibit uncertainty across a wide range of skin conditions.

Although it is crucial to accurately evaluate sensitivity and specificity in real-world usage, the low prevalence of skin cancer and the lack of serial data linking AI use to biopsy outcomes hinder comprehensive evaluation. Additionally, for general skin disorders, the sheer number of conditions makes it difficult to perform adequate external validation across diseases.

In this study, we suggested an evaluation method to calculate the sensitivity and specificity in real-world usage. To precisely assess sensitivity, we created a dataset larger than the annual number of skin cancer cases in South Korea. To evaluate specificity, we analyzed the malignancy predictions made by the algorithm in actual use. Additionally, to assess multiclass performance, we analyzed 150k images across 70 diseases. Finally, we analyzed global disease prediction patterns using the algorithm, aiming to get insights into user interests and disease prevalence.

## METHODS

This study adheres to the STARD 2015 reporting guidelines for diagnostic accuracy studies and was approved by the Kangnam Sacred Hospital Institutional Review Board (IRB number 2021-10-011, 2024-02-028-004).

The National Information Society Agency (NIA) dataset consists of 152,443 clinical photographs from 92,004 cases, covering 70 distinct skin diseases. (Table 1) Among these, 4,250 cases are malignancies, including 734 melanoma, 1,804 basal cell carcinoma (BCC), 1,601 squamous cell carcinoma (SCC), and 111 Bowen’s disease cases. The number of malignancies in the NIA dataset is comparable to Korea’s annual average number of skin cancer cases (4,498.3) based on 20 years of statistics. ^11^

**Table 1.**
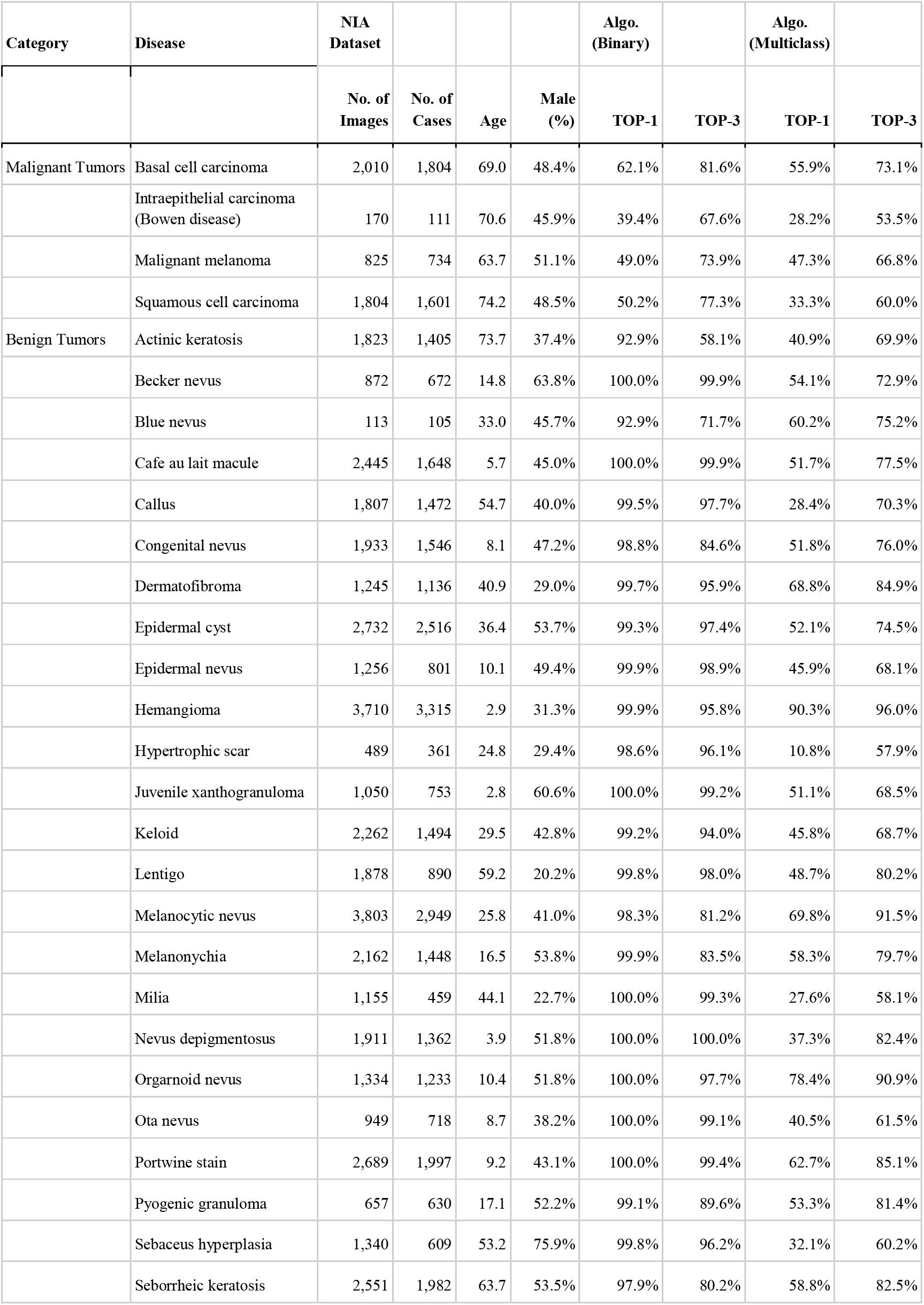
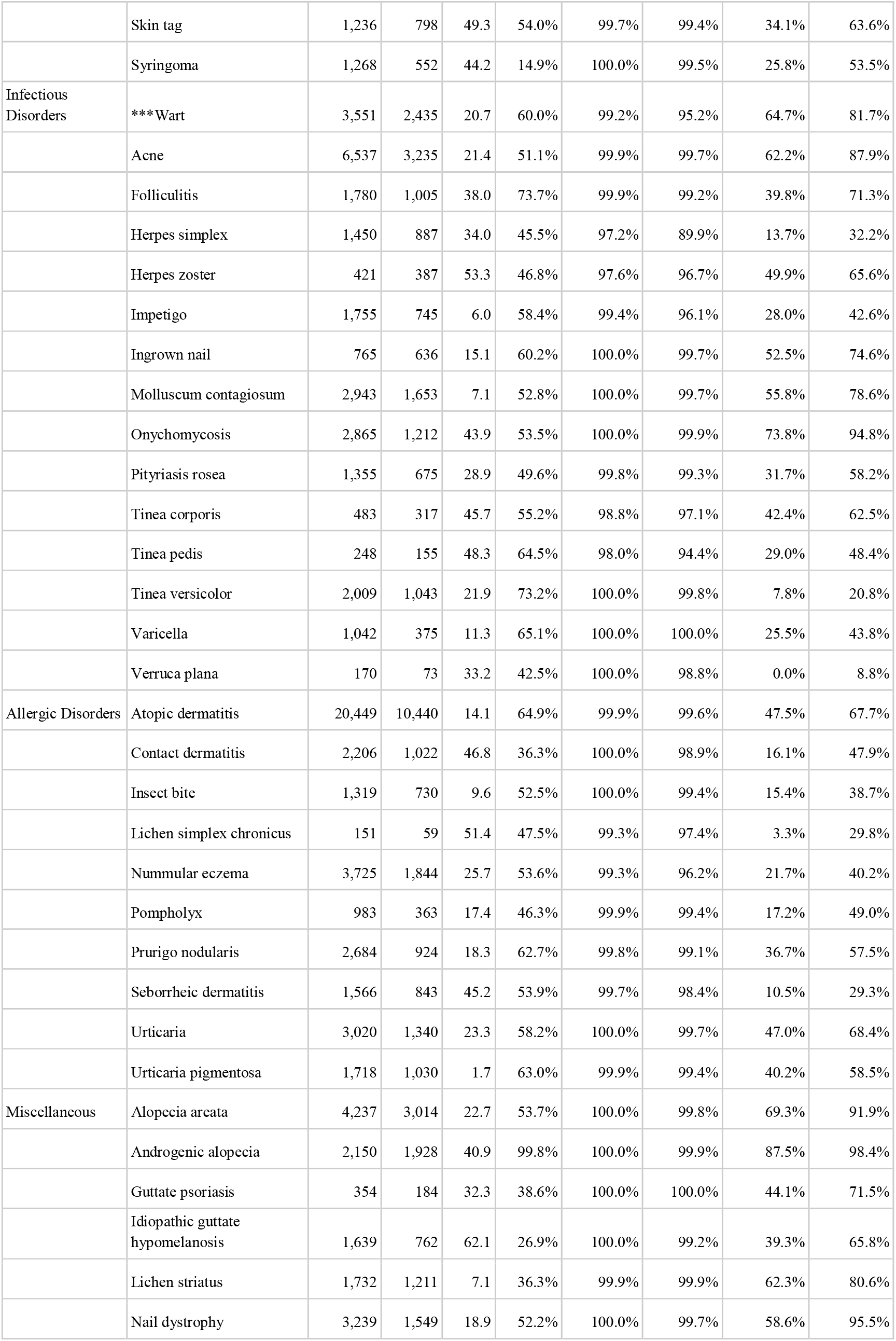
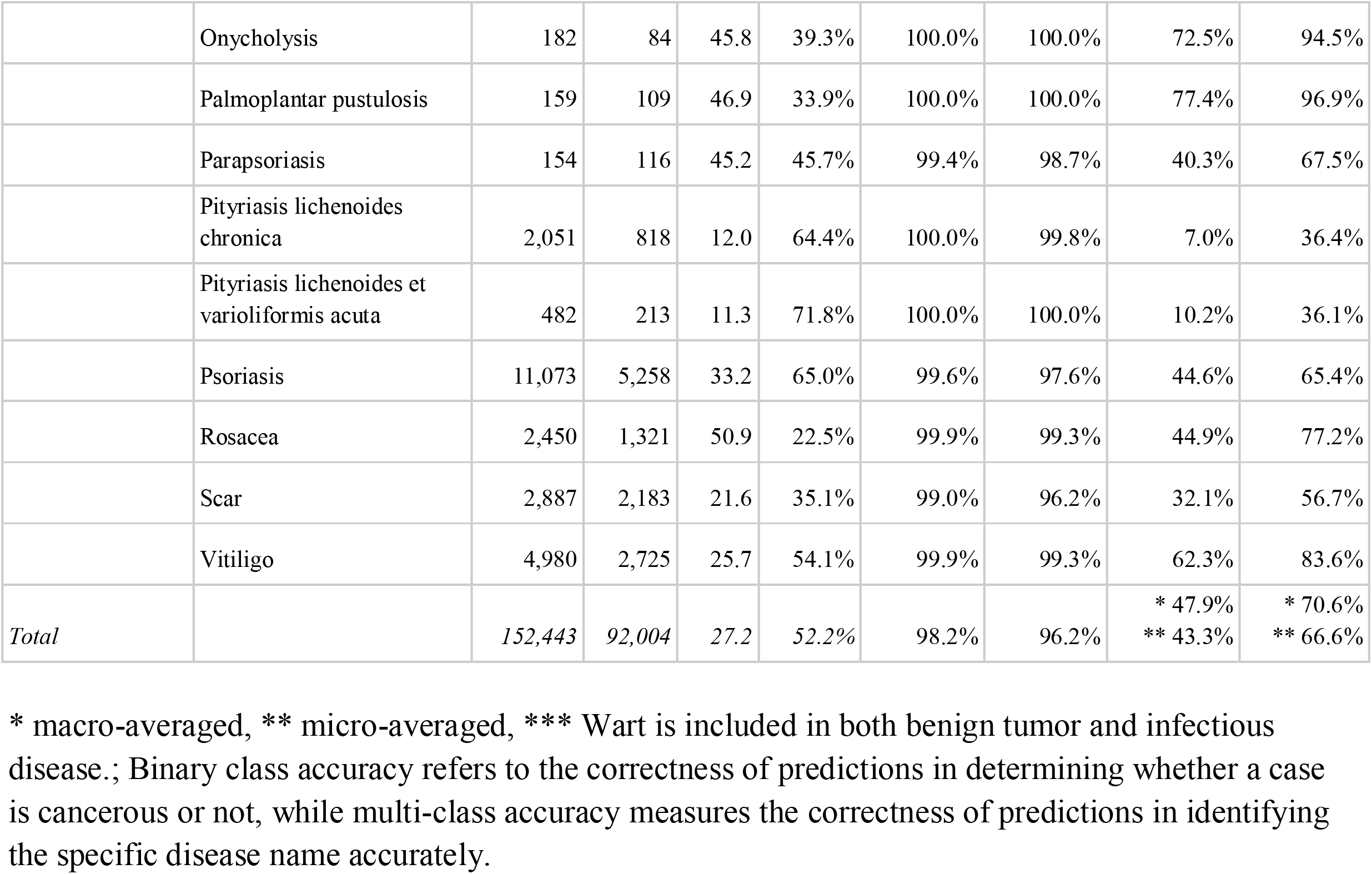
Demographics of the NIA Dataset and Top-1/3 Accuracies of the Algorithm.

Diagnoses were validated based on clinical diagnoses or pathology reports in cases of malignancy. The dataset was compiled by 63 board-certified dermatologists and 36 dermatology residents over 2 years. All photographs were manually cropped at the lesion level, and all images were reviewed by 2 dermatologists.

The SNU test dataset consists of 240 clinical photographs, covering 80 distinct diseases. (Table S1) The dataset includes 40 malignancy cases, comprising 7 Bowen’s disease, 3 keratoacanthoma, 8 BCC, 8 SCC, and 14 melanoma cases. This public dataset was compiled in a previous study that includes the reader test results from 21 dermatologists. ^3^ The Onychomycosis test dataset contains 225 clinical photographs and is also a public dataset with reader test results from 34 dermatologists in a previous study. ^12^ In this study, a global reader test was conducted using the SNU test dataset and the Onychomycosis test dataset in a mobile environment (https://www.modelderm.com/#quiz, https://www.modelderm.com/#quiz2).

We analyzed 1,690,849 queries submitted over 2.5 years (2022.7.7–2025.1.18) by 371,757 global users (Table 3, Table S4), of 6 continents (North America, South America, Asia, Europe, Oceania, and Africa). Algorithm’s predictions were categorized into four groups: benign tumors, malignant tumors, infectious diseases, and allergic diseases. (Table 1) Wart was counted under both benign tumors and infectious diseases because they are often misdiagnosed as tumors.

All datasets (NIA, SNU test, and Onychomycosis test) were tested using ModelDerm Build2024 (Supplementary Method). For global usage statistics, Build2024 was used during 2024–2025, while Build2021 ^13^ was used during 2022–2023.

### Statistics

Model performance was evaluated using sensitivity, specificity, and top-n accuracy metrics. The differences in sensitivity and specificity between the algorithm and users were tested for the paired questionnaires, using the McNemar method using R software (https://www.r-project.org; ver 4.4.2). The ROC curve was drawn by MatPlotLib (https://matplotlib.org) using the malignancy score which is composed of the output for malignant and premalignant diseases. ^3^

## RESULT

For binary classification of malignancy or not, the algorithm achieved an AUC of 0.946 (95% CI, 0.942–0.950) in the NIA dataset, utilizing only the data on benign and malignant tumors (n=51,038 images, 33 diseases) from the NIA dataset. (Fig 1a) The sensitivity and specificity of the algorithm were calculated through the three differential diagnoses suggested by the algorithm. The sensitivity / specificity of the algorithm were 78.2% / 93.0% for the NIA dataset.

**Fig 1.**
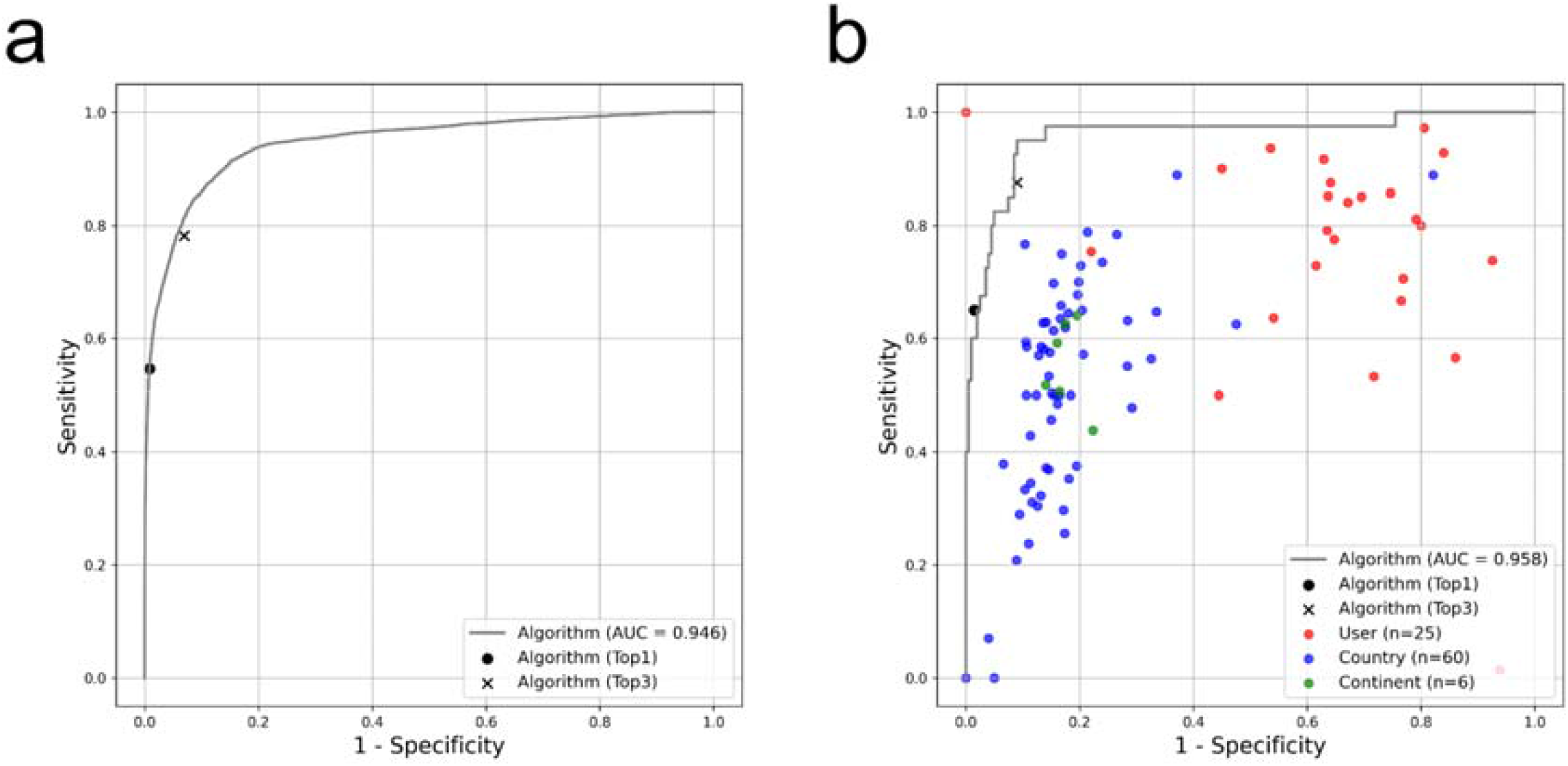
ROC curve for determining malignancy using the NIA dataset and SNU test dataset. a) TEST = Benign and malignant tumors in the NIA dataset (n=51,038 images, 33 diseases) b) TEST = SNU Test Dataset (n=240 images; 80 diseases); Results from 135 users across 60 countries and 6 continents, each with over 100 responses. Detailed data by continent and country can be found in Table S2.

For the SNU test dataset, the AUC was 0.958 (95% CI, 0.919–0.997). (Fig 1b) When calculated from three predictions of the algorithm, sensitivity / specificity were 87.5% / 91.0% for the SNU test dataset.

For multi-class classification of matching exact diagnosis, the algorithm achieved mean Top-1 / Top-3 accuracies of 47.9% / 70.6% on the NIA dataset. (Table 1) The micro-averaged Top-3 / Top-1 accuracies showed 63.3% / 83.2% on the SNU test dataset, which were comparable to the 49.9 ± 7.0% / 67.2 ± 5.4% performance reported for the SNU dataset involving 2 dermatology specialists and 2 dermatology residents. ^3^

In the global reader test, involving 61,066 assessments for determining malignancy or not, the mean sensitivity / specificity of global users were 55.2% / 84.3%, respectively, which were significantly lower than the performance of the algorithm for the same 61,066 cases (86.8% / 91.0%; p<0.0001 / p<0.0001, McNemar). (Table S2) For onychomycosis diagnosis, involving 69,931 assessments, much larger discrepancies were observed, indicating the algorithm’s superiority in diagnostic accuracy compared to general users (users = 51.9% / 57.6% vs algorithm’s Top-1 prediction = 87.9% / 78.0%). (Fig S1, Table S3)

Global usage statistics from 1,690,849 queries notable regional variations. The distribution of queries by region was as follows: Europe with 810,181, Asia with 533,106, North America with 170,948, South America with 96,373, Africa with 48,300, and Oceania with 31,941 queries. The top five countries by usage were Korea, Republic of (382,602), Italy (151,574), United States (105,233), Germany (100,285), and France (68,408). (Fig 2a, Table S4)

**Fig 2.**
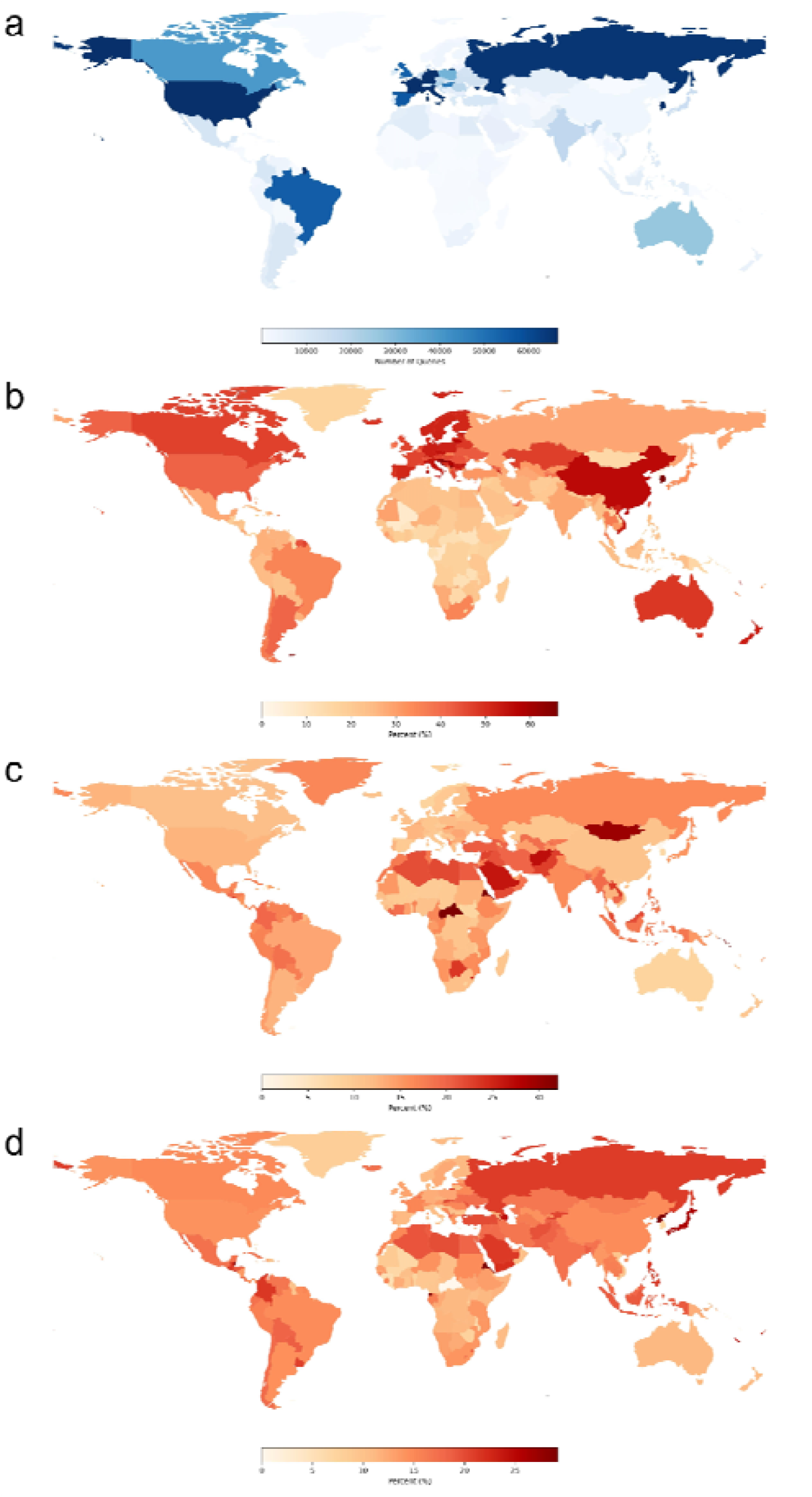
Global Distribution of Algorithm Usage and Prediction Differences by Disease Group. A worldwide map illustrating the differences in the algorithm’s predictions and usage across various disease groups. Country-specific top-1 information can be accessed at https://whria78.github.io/nia/demo a) Usage of the algorithm b) Algorithm’s prediction of either benign or malignant tumors c) Algorithm’s prediction of infectious diseases d) Algorithm’s prediction of allergic diseases

The algorithm’s predictions demonstrated distinct diagnostic trends across regions. The highest proportion of algorithm’s Top-1 predictions were for benign tumors, which accounted for 47.0%. (Table 2) In terms of benign tumor predictions, the ranking by region was as follows (all p<0.0001, McNemar): Asia (55.5%) > Oceania (46.8%) > Europe (46.5%) > North America (38.0%) > South America (32.3%) > Africa (22.4%).

**Table 2.**
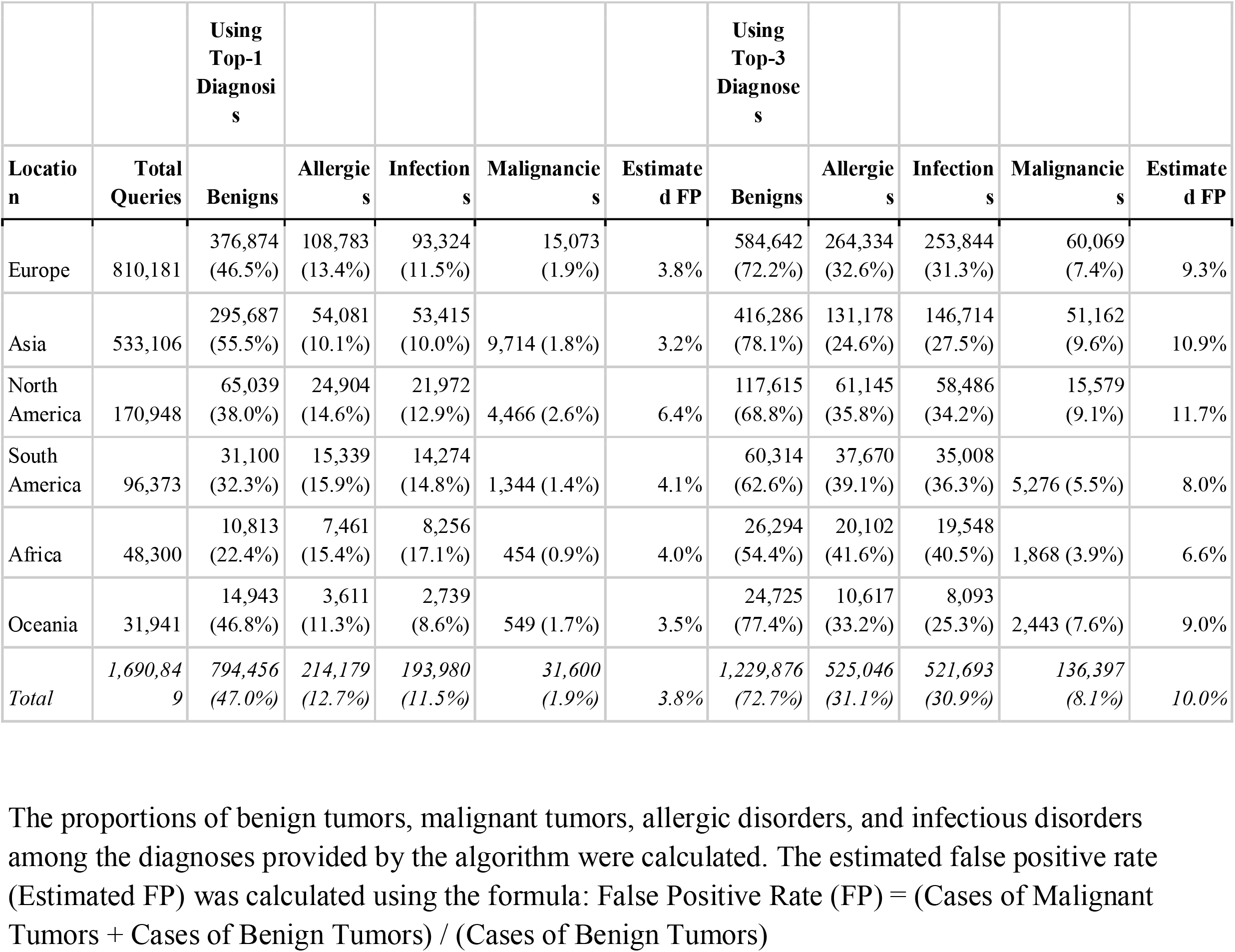
Proportions of Disease Groups Obtained from the Algorithm’s Top-1 and Top-3 Predictions (by Continent) The proportions of benign tumors, malignant tumors, allergic disorders, and infectious disorders among the diagnoses provided by the algorithm were calculated. The estimated false positive rate (Estimated FP) was calculated using the formula: False Positive Rate (FP) = (Cases of Malignant Tumors + Cases of Benign Tumors) / (Cases of Benign Tumors)

Regarding malignancy predictions, the rate was 3.8% for Top-1 predictions and 10.0% for Top-3 predictions in Korea. For the Top-3 predictions, the regional distribution of malignancy was as follows (all p<0.0001, McNemar): North America (11.7%) > Asia (10.9%) > Europe (9.3%) > Oceania (9.0%) > South America (8.0%) > Africa (6.6%). (Table 2)

Infectious conditions were predominantly predicted in Africa (17.1%) and South America (14.8%) than other regions like North America (12.9%), Europe (11.5%), Asia (10.0%), and Oceania (8.6%), with higher frequencies observed in the Middle East and Northern Africa on the country map (Fig 2c). Allergic diseases showed relatively consistent trends across regions (Fig 2d).

## DISCUSSION

There is a significant performance gap between diagnosing based on images alone and diagnosing in actual clinical practice. For the diagnosis of 43 tumors, attending physicians achieved Top-1 / Top-3 accuracies of 68.1% / 77.3%, while physicians in the reader test had only 37.7% / 53.4%. ^10^ Although AI has demonstrated exceptional performance in controlled settings, it must prove its effectiveness in real-world environments to be truly meaningful in clinical practice. ^14,15^ Despite the large number of retrospective studies conducted on AI in healthcare, real-world evidence remains still insufficient. As of 2024, only 86 medical AI systems have reported RCT-level evidence, and among these, 70 achieved successful outcomes. ^16^

Evaluating the real-world performance of AI in dermatology is highly challenging. To assess the real-world performance of AI in skin cancer diagnosis, a continuous dataset linking AI results to biopsy outcomes conducted in hospitals is required. However, due to the low prevalence of skin cancer and the current lack of digitalization in clinical practice, obtaining large datasets with serial results is virtually impossible. Additionally, evaluating specificity is problematic, as curated hospital datasets fail to capture the wide range of out-of-distribution conditions encountered in real-world settings.

In this study, we curated a large dataset representing cancer cases in Korea to estimate the sensitivity of the AI for cancer diagnosis. To assess specificity, we analyzed usage statistics under the assumption that all AI-determined malignancy predictions were false positives. This approach enabled us to estimate the algorithm’s real-world performance in terms of sensitivity and specificity for the binary classification of malignancy.

Furthermore, to evaluate multiclass accuracy, where the algorithm correctly matches the disease to its specific name, we constructed a dataset containing sufficient cases for 70 common dermatological diseases. This allowed us to provide an analysis of the AI’s diagnostic capabilities across detailed data for each disease.

First, regarding the sensitivity, the NIA dataset is large enough to represent skin cancer cases in Korea. Therefore, the sensitivity of 78.2%, predicted by three differentials (Top-3 diagnoses), was presented as the ideal maximum value achievable by the algorithm in real-world settings. if users capture only low-quality images, the sensitivity could be lower. ^13,17^ However, in actual use, users may test a single lesion multiple times over several days, which could result in higher sensitivity.

Next, regarding specificity, the analysis of usage records in Korea showed that the algorithm predicted malignancy at a rate of 12.0% based on Top-3 predictions. Given the very low prevalence of cancer, if we assume all malignancy predictions by the model are false positives, the specificity was estimated at 88.0% from Top-3 predictions. This figure (88.0%) is lower than the specificity measured on the NIA dataset (93.0%; Table 1). At first glance, we may expect that benign tumors significant enough to warrant a hospital visit would be harder to diagnose than those observed in everyday life. However, unlike the curated benign cases from hospitals, real-world usage involves various out-of-distribution conditions, which likely contributed to this difference in AI performance. However, the actual specificity may be slightly higher than our estimate of 88.0%, as users often confirm results labeled as malignant through repeated tests before visiting the hospital. In addition, the assumption that all malignancy predictions are false positives suggests the actual specificity of the algorithm was likely somewhat higher.

From the perspective of disease screening, the WHO guidelines for tuberculosis diagnosis set minimal requirements, including a sensitivity of 90% and a specificity of 70%. ^18^ Similarly, the Breast Cancer Surveillance Consortium benchmarks highlight a sensitivity of 86.9% at a specificity of 88.9% for breast cancer screening. ^19^ However, for skin cancer screening, no precise guidelines for the sensitivity and specificity currently exist. Furthermore, there are concerns about overdiagnosis even when dermatologists conduct skin cancer screenings, which raises the need for further discussion on how algorithms with performance levels still below that of dermatologists could be utilized for screening purposes. ^20^ However, unlike human physicians, algorithms can be adjusted to operate with either very high specificity or sensitivity by modifying the threshold. For example, in scenarios where concerns about overdiagnosis necessitate more specific screening, the algorithm’s threshold could be raised to identify only cancers with the highest diagnostic certainty, ensuring that only the most definitive cases are flagged, particularly in populations with limited access to healthcare. From this perspective, it is necessary to identify proper settings and demonstrate clinical outcomes (mortality or morbidity) in the future. In the reader test, the algorithm performed much better than the general population in diagnosing onychomycosis. (Fig S1) Since onychomycosis carries a lower burden of misdiagnosis, it would be beneficial to first evaluate the strengths and limitations of AI screening in such common benign conditions before gradually expanding its application to more critical areas like skin cancer screening.

The usage statistics revealed significant regional differences in the types of diseases predicted by the algorithm. (Fig 2, Fig S2, Table 3, Table S4) Tumorous disorders were more commonly predicted in Asia, Europe, and the USA. (Fig 2a) On the other hand, premalignant conditions such as actinic keratosis were observed at higher rates in Austria and the USA. (Fig S2I) In contrast, infectious diseases were more frequently observed in regions such as Northern Africa and the Middle East. (Fig 2c) These regional differences reflect variations in disease prevalence, the age demographics of users, and the diseases users are particularly concerned about in each region. These data, collected before patients even visit the clinic, can offer more accurate insights into the prevalence and interest in specific skin conditions. This indicates that further research using AI-based big data analytics could significantly contribute to understanding skin disease trends globally.

**Table 3.**
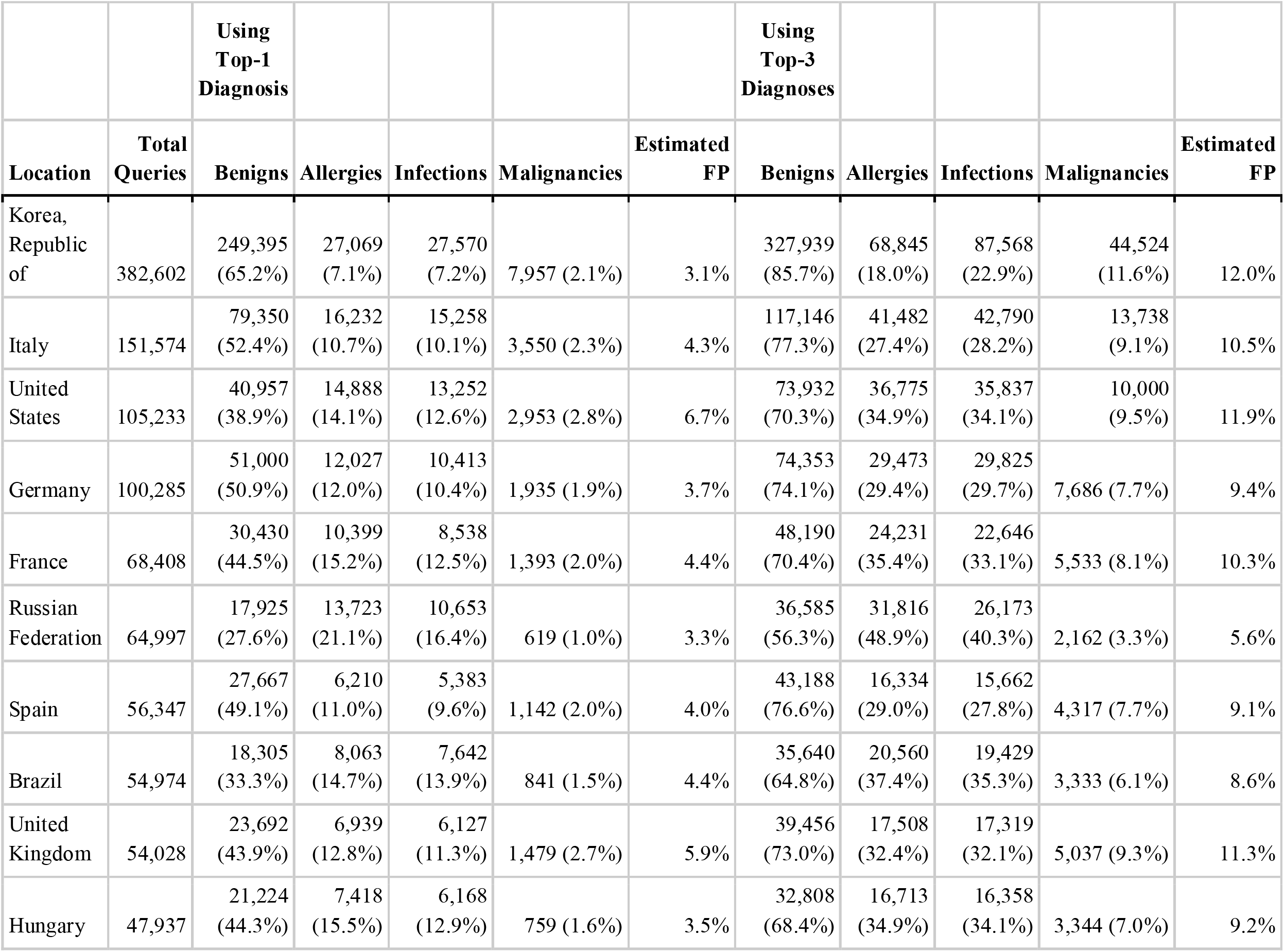
Proportions of Disease Groups Obtained from the Algorithm’s Top-1 and Top-3 Predictions (by Country) This table shows statistics for the 10 countries with the highest usage, calculated using the same method as in Table 2.

## Limitation

First, this study was limited to patients with skin types III and IV. Additional studies are required to evaluate the sensitivity of the algorithm for skin cancer in White and Black populations.

Second, the NIA dataset does not include less common skin cancers beyond the four major types. According to 20 years of incidence statistics in Korea, these other types of skin cancers account for approximately 11.3% of all skin cancers ^11^.

Third, the sensitivity and specificity calculated in this study will need to be re-evaluated through further digital transformation (DX) efforts. Such efforts should be conducted separately for each indication where the algorithm is applied.

Finally, in terms of multi-class results, the algorithm achieved Top-1 and Top-3 micro-averaged accuracies of 43.3% and 66.6% on the multi-class NIA dataset. This replicates the performance observed in previous studies and proves that the algorithm has reached the level of dermatologists in the multi-class classification. ^3^ Although the multiclass performance of the algorithm was validated using the large curated NIA dataset, its real-world performance will also need to be reassessed in future studies in real-world settings.

## Conclusion

Using a large curated dataset and usage data, the real-world performance of AI in diagnosing skin cancer in Korea could be estimated at 78.2% sensitivity and 88.0% specificity. In multi-class classification, our algorithm achieved Top-1 and Top-3 accuracies of 43.3% and 66.6%, respectively, replicating the dermatology-level performance. ^3^ Furthermore, this study highlights the potential of AI algorithms to provide a global perspective on skin diseases, offering a quantitative reflection of regional variations. Future research is needed to determine the specific clinical settings in which AI can effectively improve clinical outcomes.

## Abbreviations

AI: artificial intelligence
AUC: area under the curve
BCC: basal cell carcinoma
CNN: convolutional neural network
RCT: randomized controlled trial
ROC curve: receiver operating characteristic curve
SCC: squamous cell carcinoma
DX: digital transformation

## DATA AVAILABILITY

Interactive demo showing the global distribution of the Top-1 disease:

https://whria78.github.io/nia/demo

The SQL dump of usage logs is available at:

https://whria78.github.io/nia

The NIA dataset was created under the leadership of the National Information Society Agency, and offline access is only available with IRB approval in the offline zone of the AI Hub (https://aihub.or.kr).

## ACKNOWLEDGEMENTS

We would like to thank the global participants in the reader test. Han and Cho have full access to all the data used in the study and take responsibility for the integrity of the data and accuracy of the data analysis.

## AUTHOR CONTRIBUTIONS

Han, Na, and Cho designed the experiments.

Cho, Na, Kim, and Han prepared the datasets and performed the experiments.

Han, Cho, Na, and Kim interpreted the results.

Han, Cho, Na, and Kim wrote the manuscript and prepared the figures.

Han, Na, and Cho supervised the entire experiment.

All authors approved the final version of the manuscript.

## Notes

Conflict of interest: SS Han is the founder and CEO of IDerma, Inc.; SI CHo is an employee of Lunit. All other authors have no conflict of interest.

### Competing Interest Statement

SS Han is the founder and CEO of IDerma, Inc.; SI CHo is an employee of Lunit. All other authors have no conflict of interest.

### Funding Statement

None

### Author Declarations

Approved by the Kangnam Sacred Hospital Institutional Review Board (IRB number 2021-10-011, 2024-02-028-004).

