## Supplementary Method for "Validation of an AI for Skin Diseases in Korea and Global Usage Statistics"

**SUPPLEMENTARY APPENDIX**

**Supplementary Method**

ModelDerm (<https://modelderm.com>) has been developed and improved since 2017 through several studies. In a study ^1^ , it was confirmed that the model outperformed specialists in classifying 10 major diseases and over 200 secondary diseases after training using CNN (convolutional neural network). However, when the hospital dataset was used for training without proper handling, inappropriate predictions were observed in the demo version, and Cristian and Kostantinos reported the issues of the algorithm in a letter article. ^2^ Another algorithm for onychomycosis, developed around the same time, demonstrated that using a region-based CNN for predicting lesions, combined with annotating lesions across a large dataset, resulted in much more accurate outcomes. ^3^ A similar approach was applied recursively to general disorders, resulting in the validation of 134 diseases, with performance equivalent to that of dermatology residents in terms of binary and multiclass accuracy. ^4^ In addition to disease diagnosis (classification), the algorithm also showed similar results to dermatologists in detecting skin cancer (object detection). ^5^

The architecture of the algorithm is an ensemble of ResNet variants. ^6^ By 2020, over a million lesional crops, including images collected from the internet alongside publicly available datasets, were used to further improve the model (Build2020). Since Build2020, the training dataset has undergone minor revisions, which included correcting label accuracy. The performance of Build2020, Build2021, and Build2024 on public datasets (Edinburgh dermofit, <https://licensing.edinburgh-innovations.ed.ac.uk/product/dermofit-image-library>) has shown slight improvement via data-centric approach. (AUC, Build2020 = 0.937; Build2021 = 0.935, Build2024 = 0.947)

Recently, we demonstrated that annotating many images from the internet produced better results than using hospital datasets. ^7^ The algorithm’s performance in real-world usage was proven through an RCT, showing that it could improve the diagnostic rate of non-specialists. ^8^ The algorithm marked the CE MDR class I (May 28, 2021), but the app is currently positioned as an information search tool.

**Studies related Model Development**

Assessment of Deep Neural Networks for the Diagnosis of Benign and Malignant Skin Neoplasms in Comparison with Dermatologists: A Retrospective Validation Study. PLOS Medicine, 2020

Performance of a deep neural network in teledermatology: a single‐center prospective diagnostic study. J Eur Acad Dermatol Venereol. 2020

Keratinocytic Skin Cancer Detection on the Face using Region-based Convolutional Neural Network. JAMA Dermatol. 2019

Seems to be low, but is it really poor? : Need for Cohort and Comparative studies to Clarify Performance of Deep Neural Networks. J Invest Dermatol. 2020

Multiclass Artificial Intelligence in Dermatology: Progress but Still Room for Improvement. J Invest Dermatol. 2020

Augment Intelligence Dermatology : Deep Neural Networks Empower Medical Professionals in Diagnosing Skin Cancer and Predicting Treatment Options for 134 Skin Disorders. J Invest Dermatol. 2020

Interpretation of the Outputs of Deep Learning Model trained with Skin Cancer Dataset. J Invest Dermatol. 2018

Automated Dermatological Diagnosis: Hype or Reality? J Invest Dermatol. 2018

Classification of the Clinical Images for Benign and Malignant Cutaneous Tumors Using a Deep Learning Algorithm. J Invest Dermatol. 2018

Augmenting the Accuracy of Trainee Doctors in Diagnosing Skin Lesions Suspected of Skin Neoplasms in a Real-World Setting: A Prospective Controlled Before and After Study. PLOS One, 2022

Evaluation of Artificial Intelligence-assisted Diagnosis of Skin Neoplasms – a single-center, paralleled, unmasked, randomized controlled trial. J Invest Dermatol. 2022

The Degradation of Performance of a State-of-the-art Skin Image Classifier When Applied to Patient-driven Internet Search. Scientific Report 2022

Generation of a melanoma and nevus data set from unstandardized clinical photographs on the internet. JAMA dermatology 2023

Melanoma detection: Evaluating the classification performance of a deep convolutional neural network and dermatologist assessment via a mobile app in an Italian real-world setting. J Eur Acad Dermatol Venereol. 2024

**Fig S1 - ROC curve for determining malignancy using the Onychomycosis test dataset**

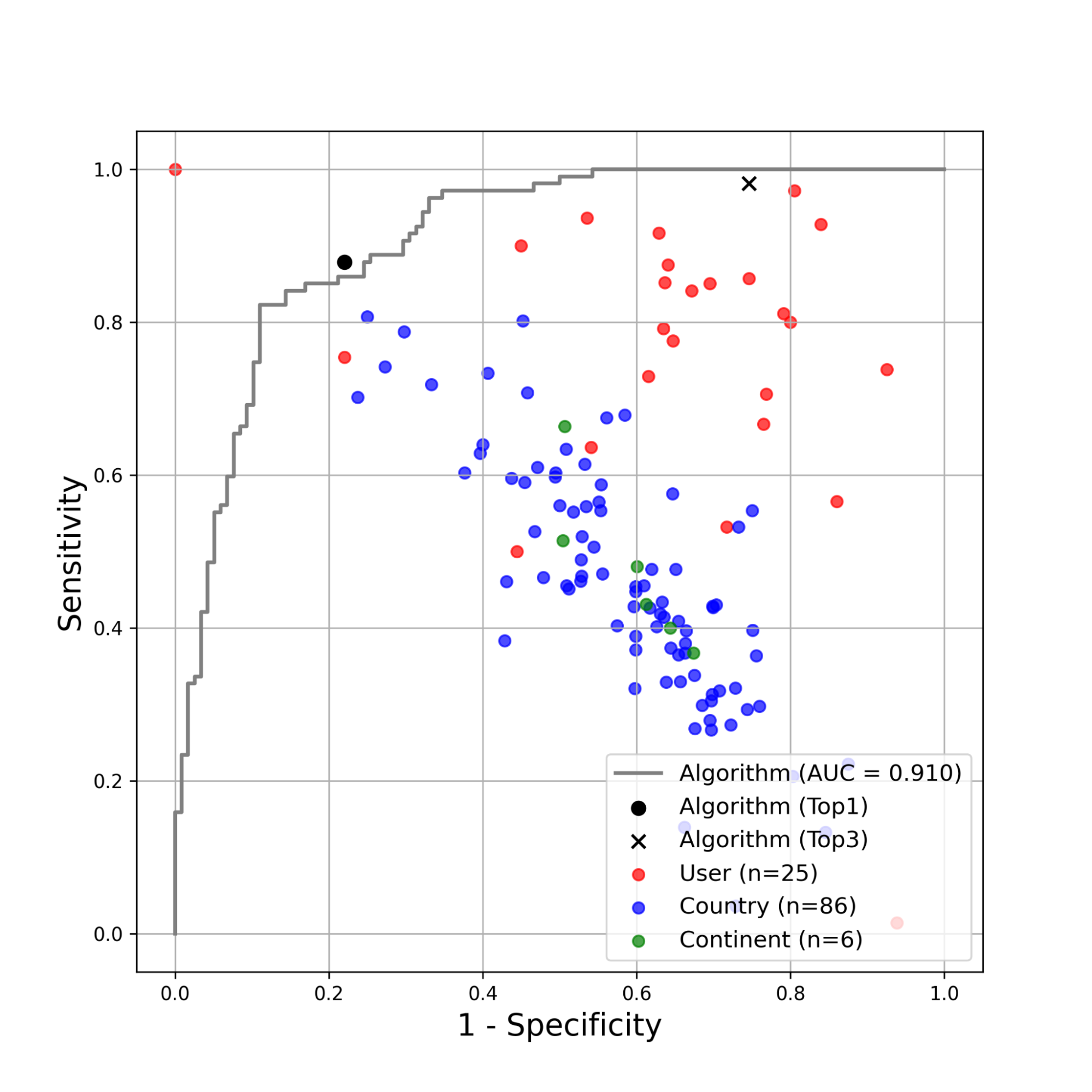

TEST = Onychomycosis Dataset (225 images; Onychomycosis and Nail dystrophy)

Results from 86 countries with over 100 responses, 25 users, and 6 continents. The test dataset (B1+C) is available for download at :

<https://figshare.com/articles/dataset/Model_Onychomycosis_Training_Datasets_JPG_thumbnails_and_Validation_Datasets_JPG_images_/5398573>

**Fig S2 - Global Distribution of Algorithm Prediction Differences by Disease**

A worldwide map illustrating how the algorithm's predictions and usage vary across common diseases. Globally, the algorithm's usage by disease is as follows: Seborrheic keratosis (77,257), Cherry Hemangioma (36,906), Dermatofibroma (35,215), Folliculitis (34,246), Wart (33,982), Lentigo (33,440), Hemangioma (33,174), Urticaria (32,763), and Actinic Keratosis (32,354). Country-specific top-1 information can be accessed at : <https://whria78.github.io/nia/demo>

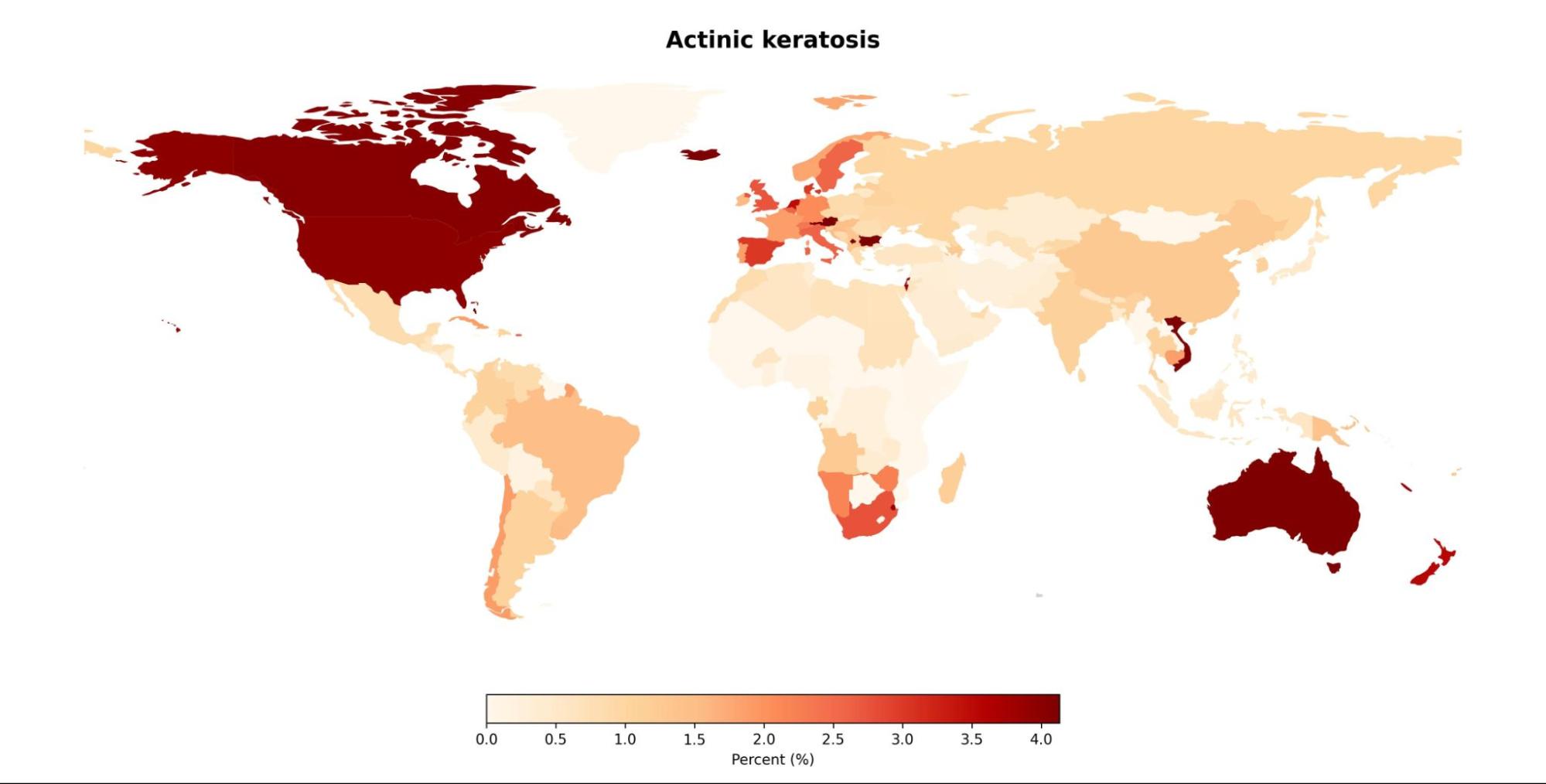

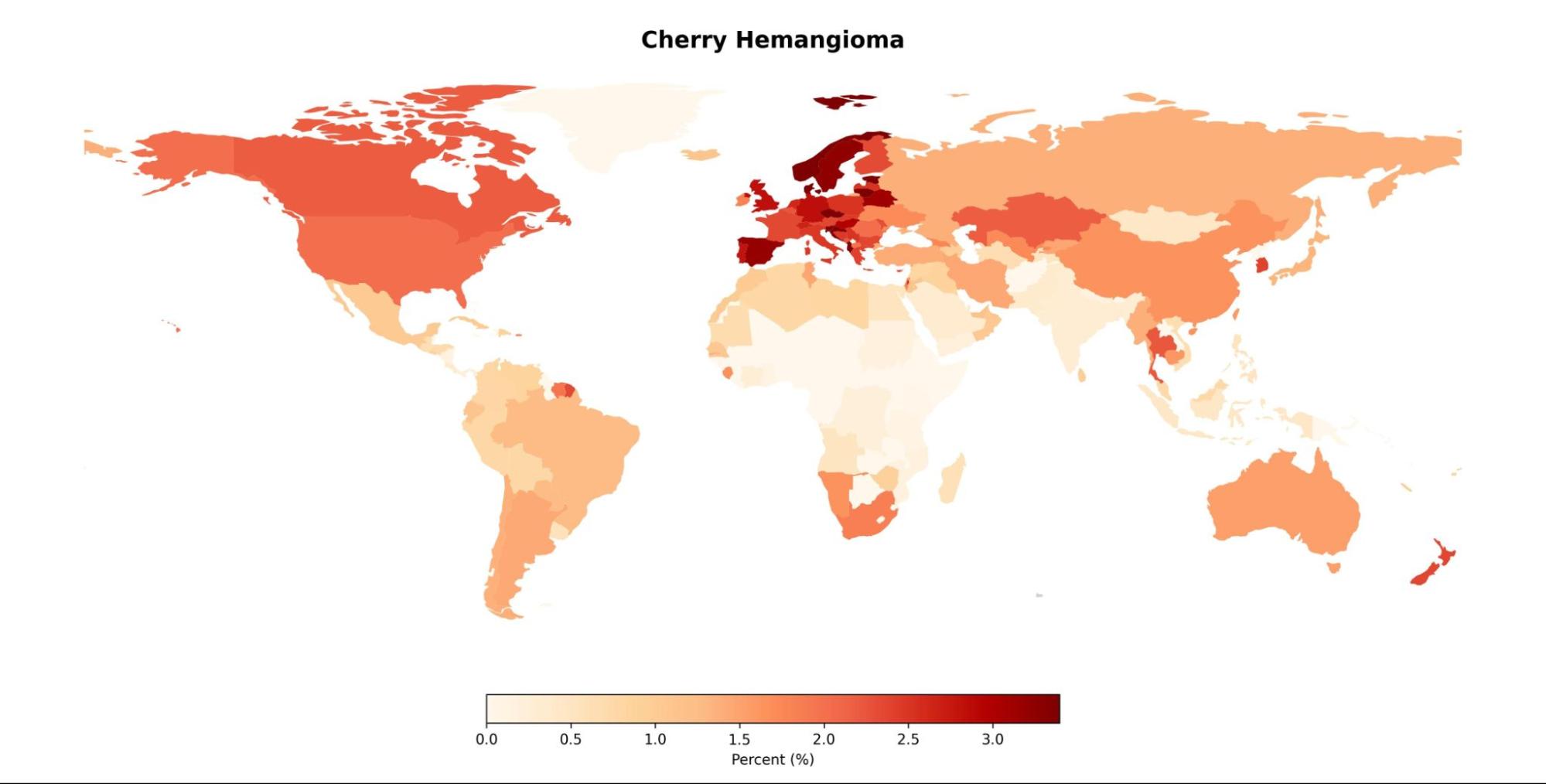

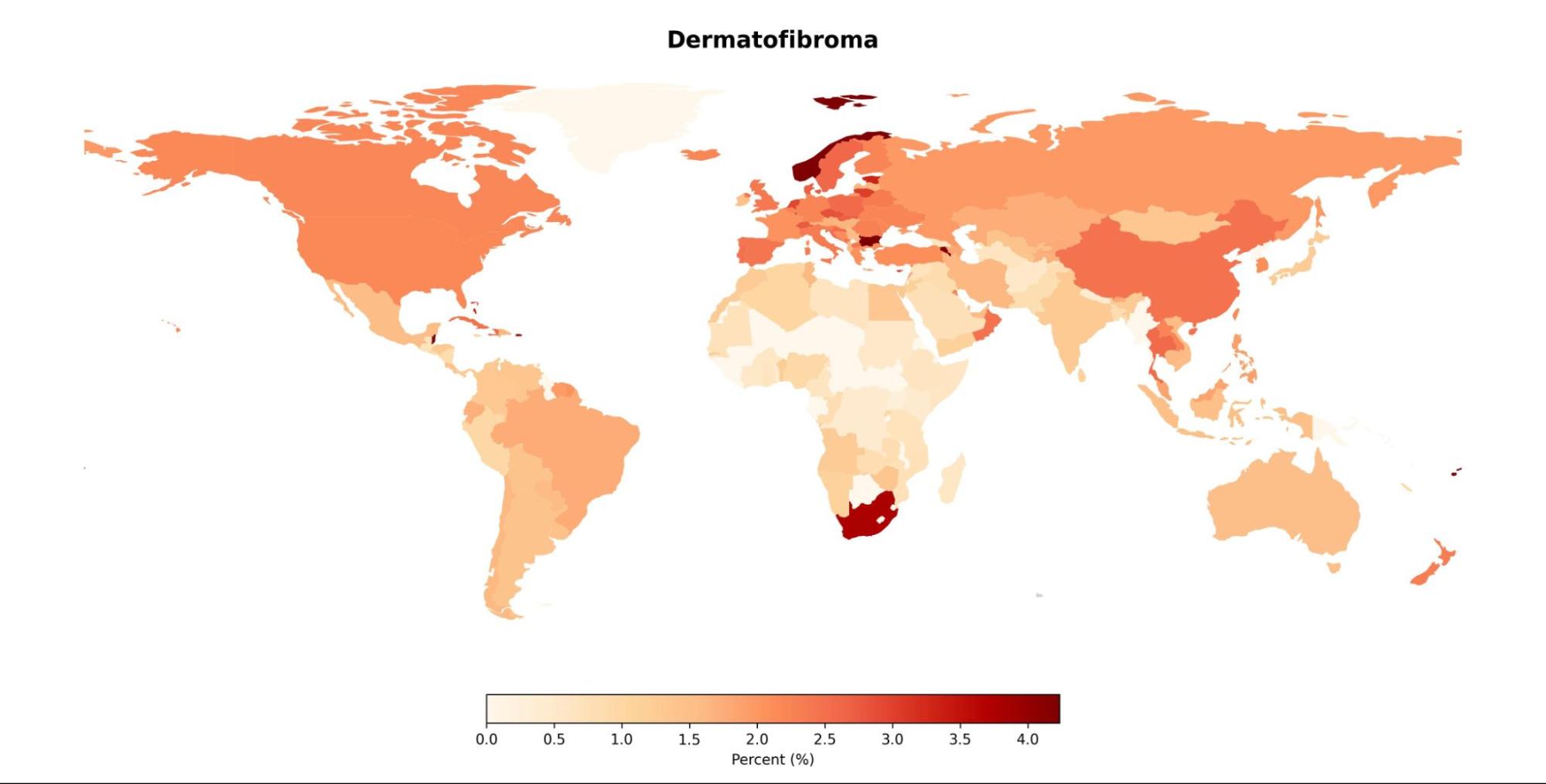

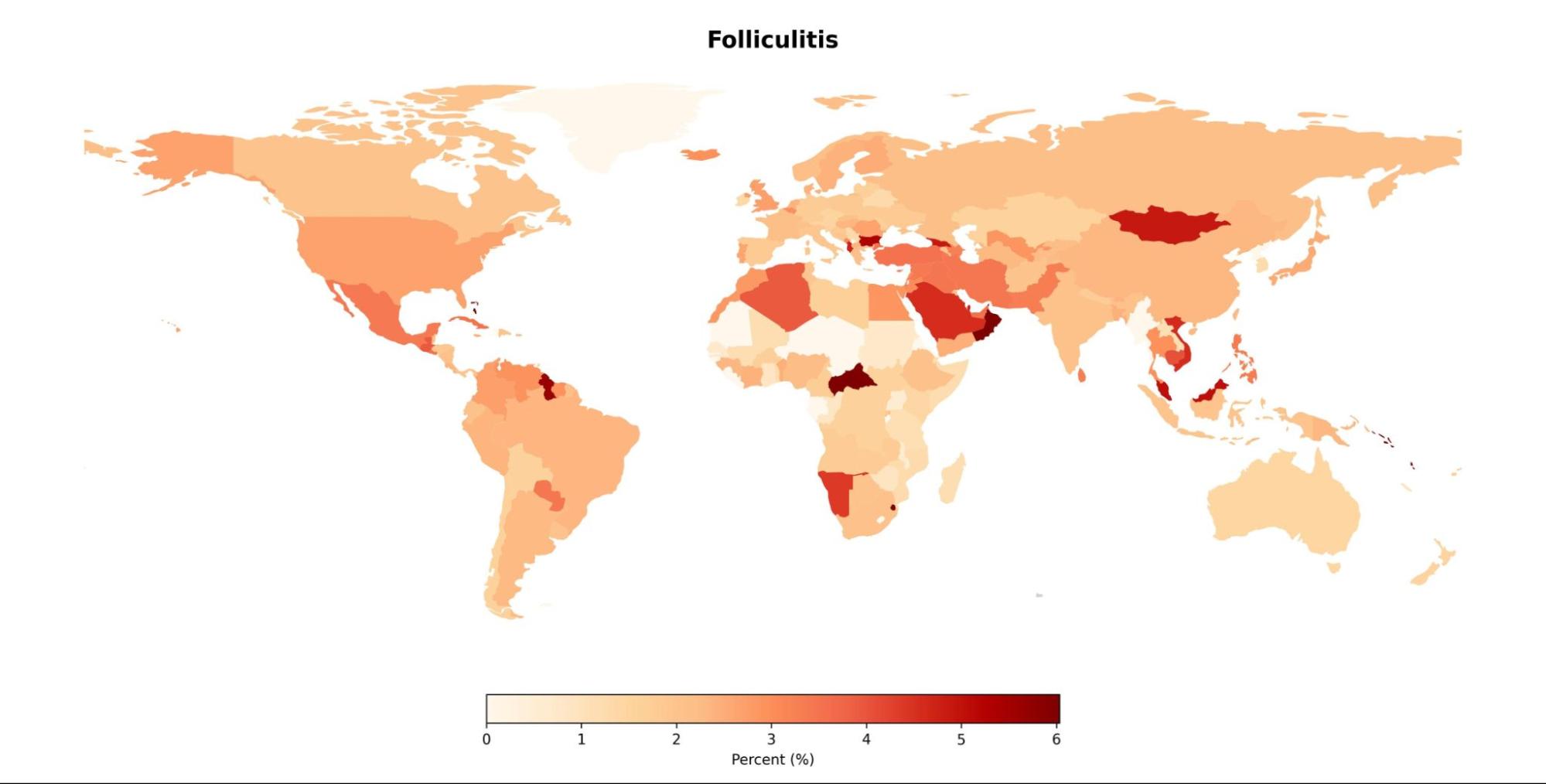

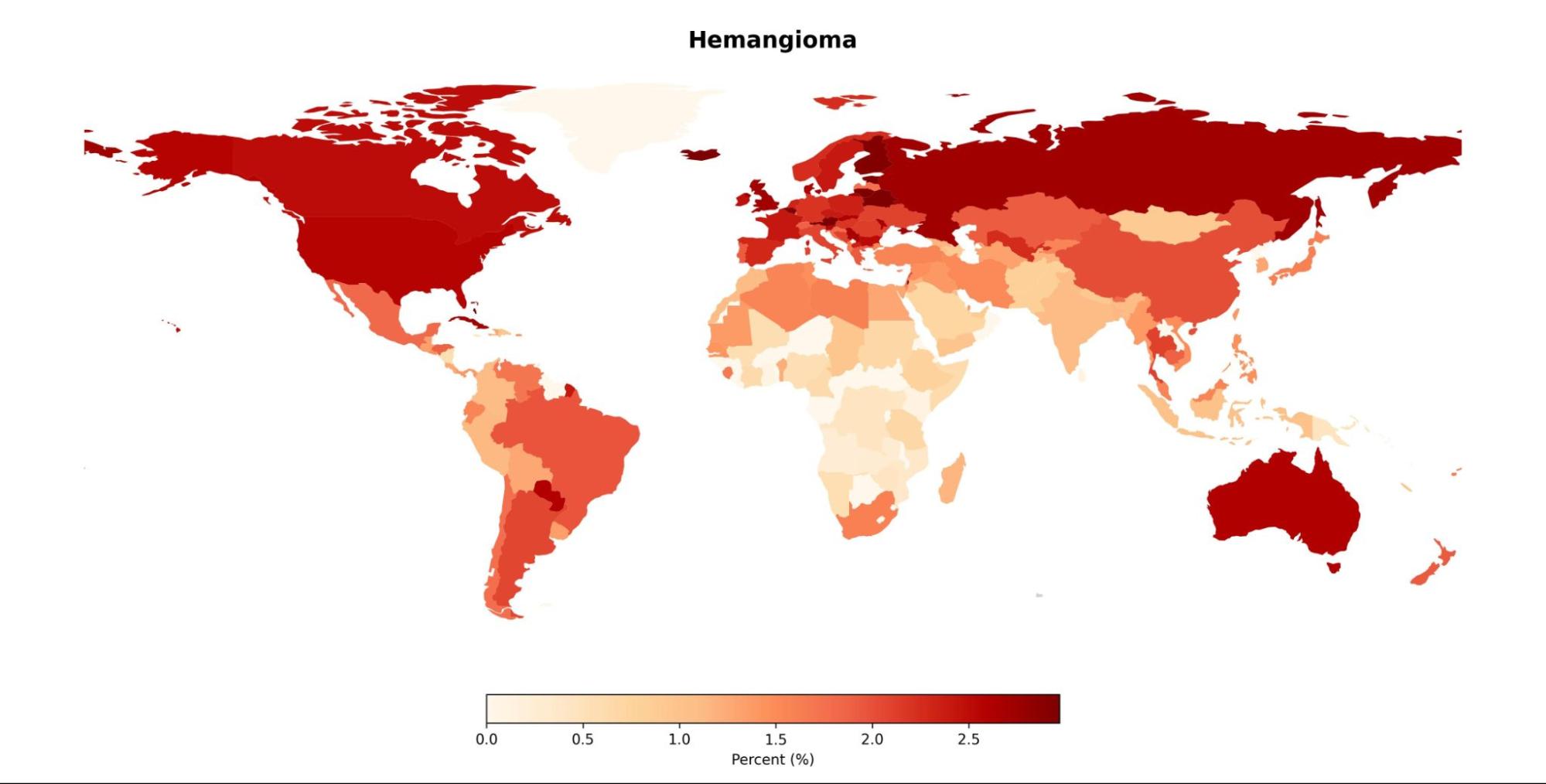

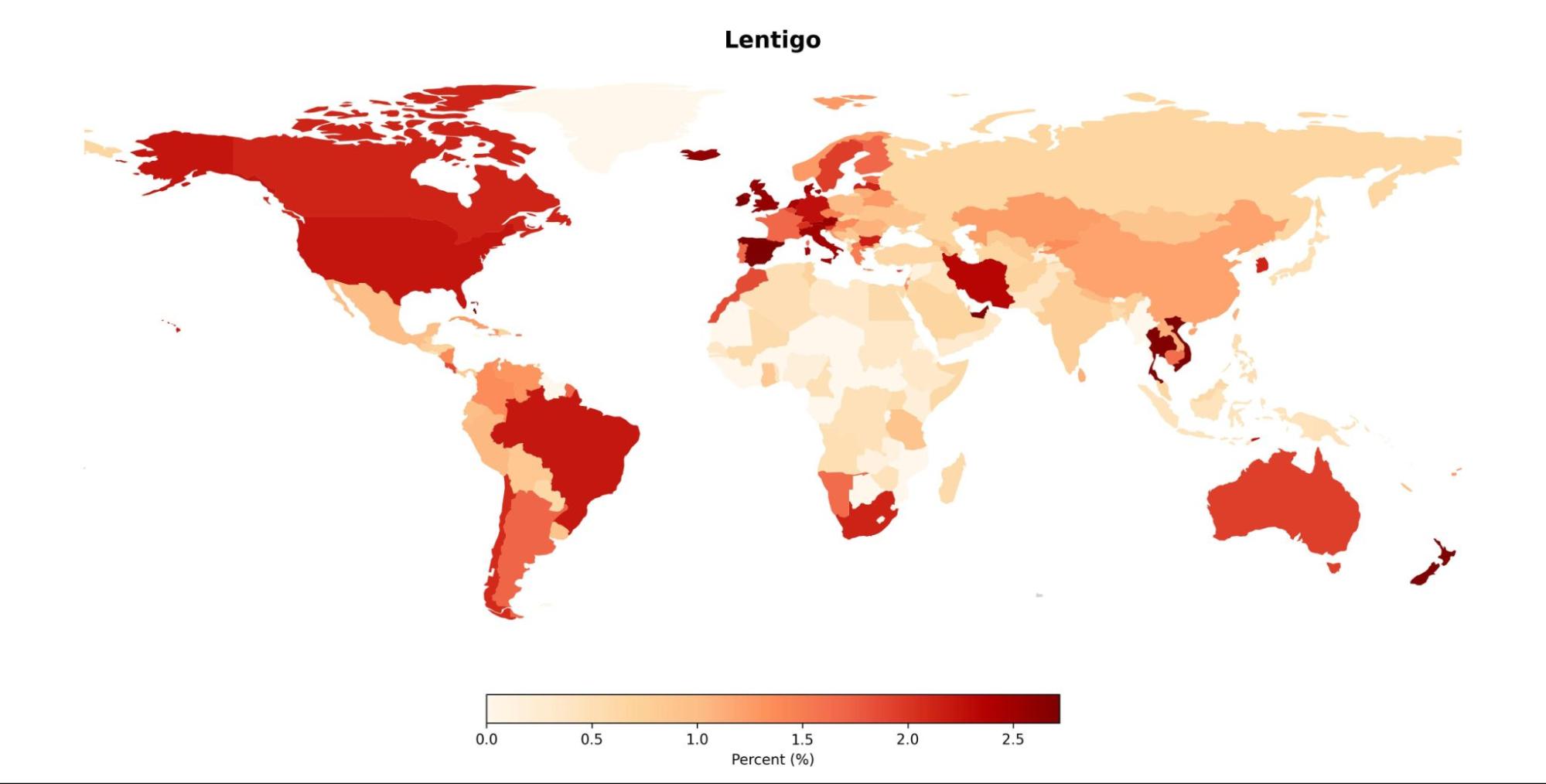

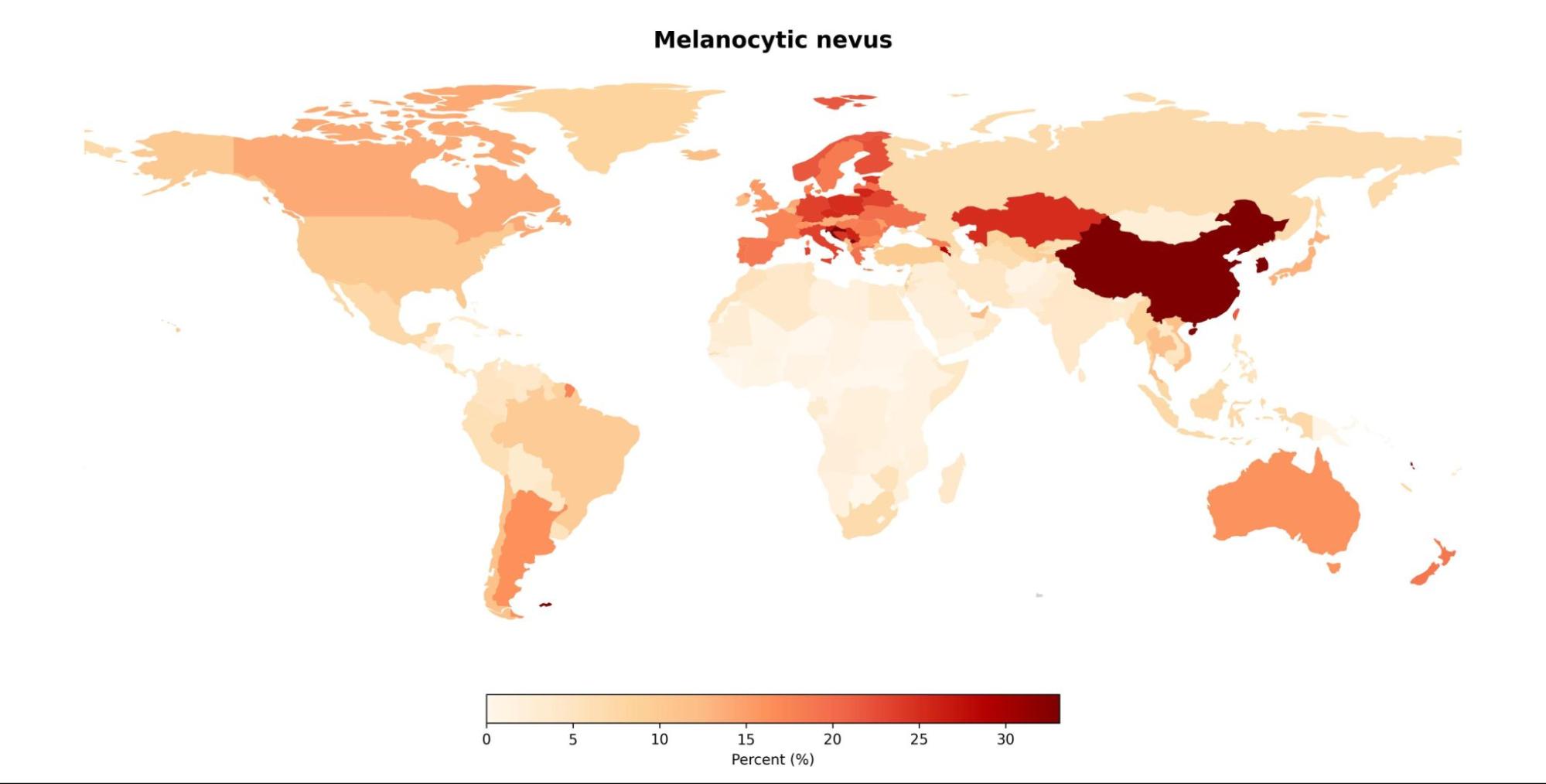

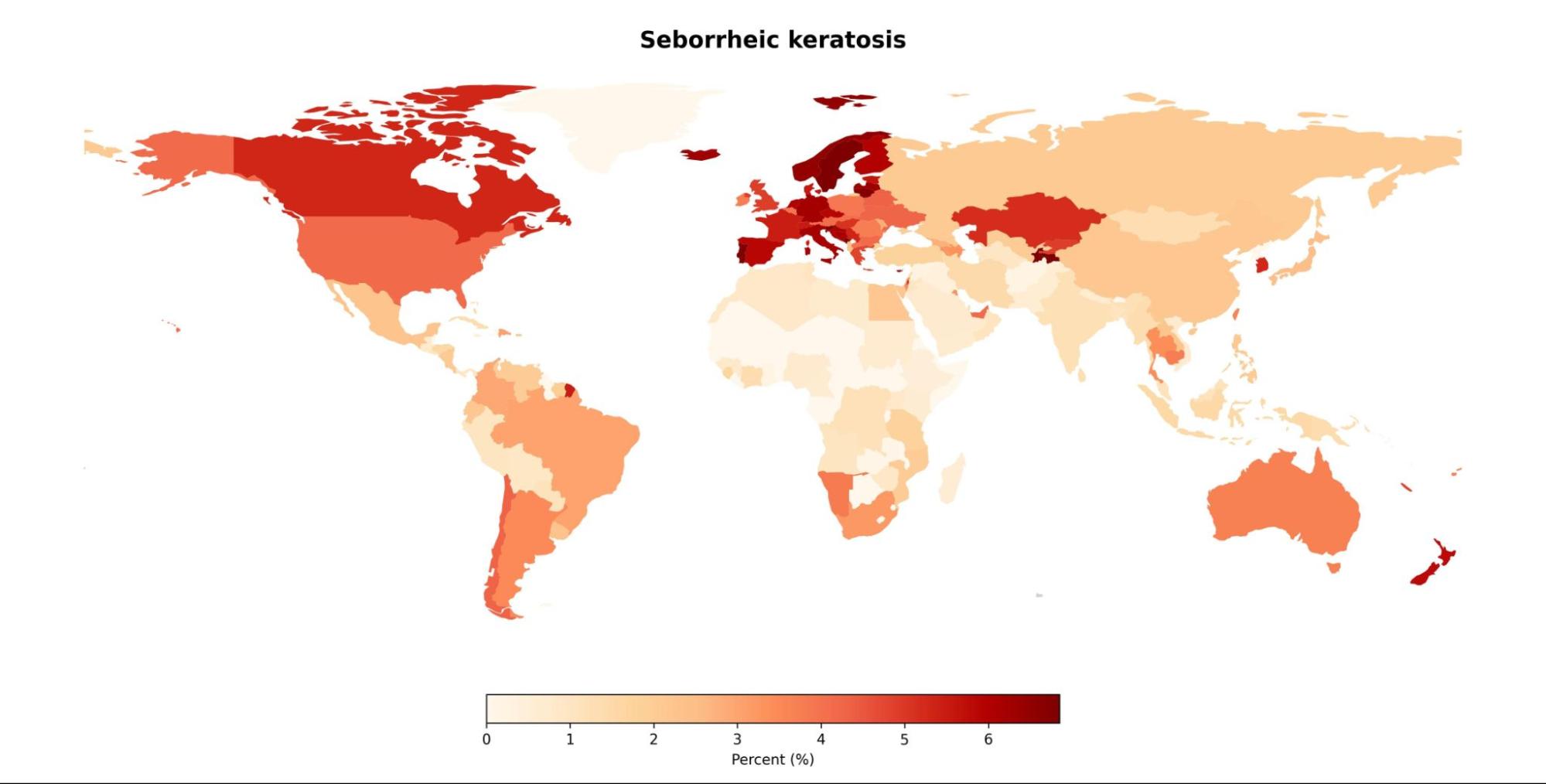

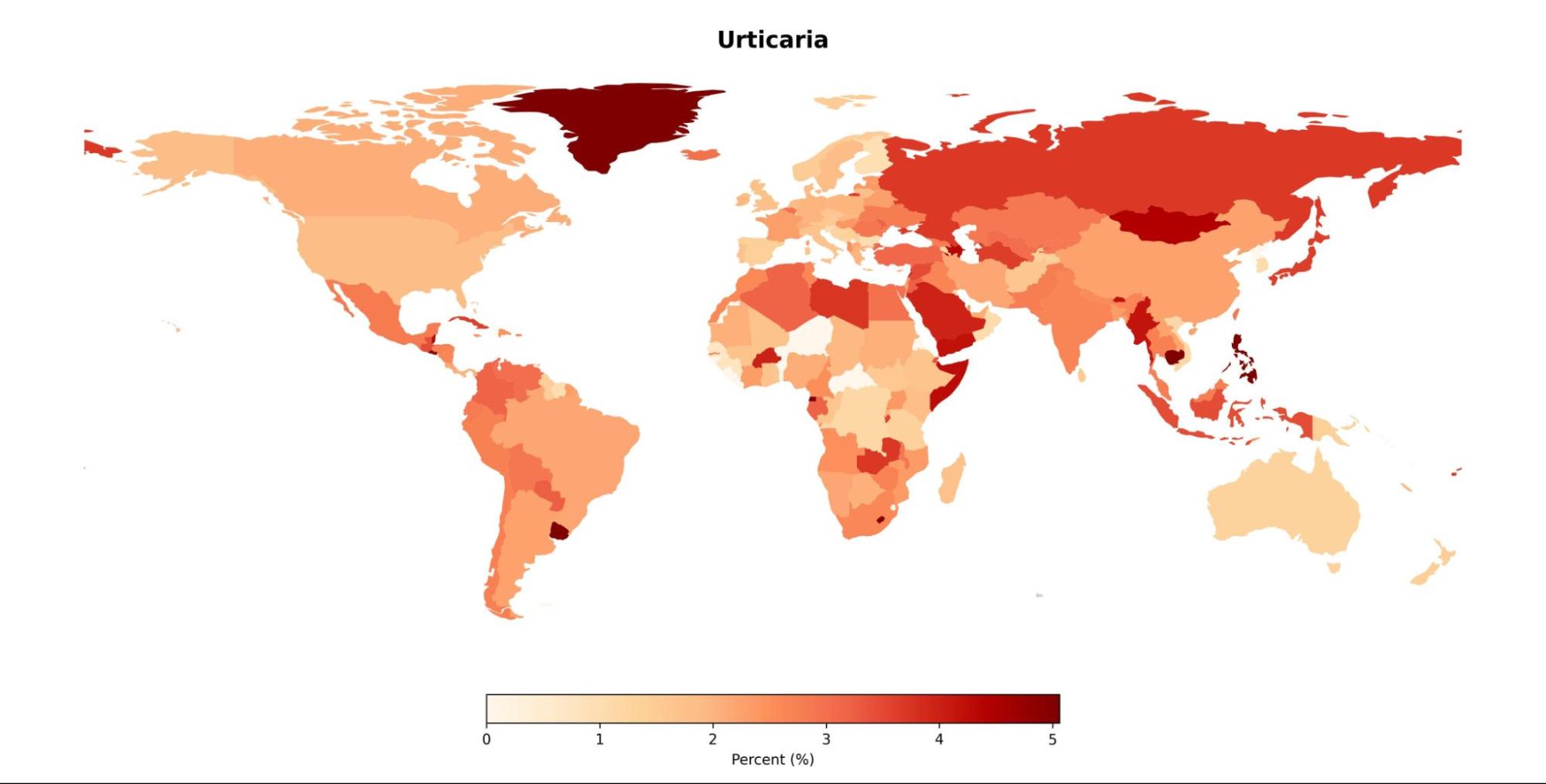

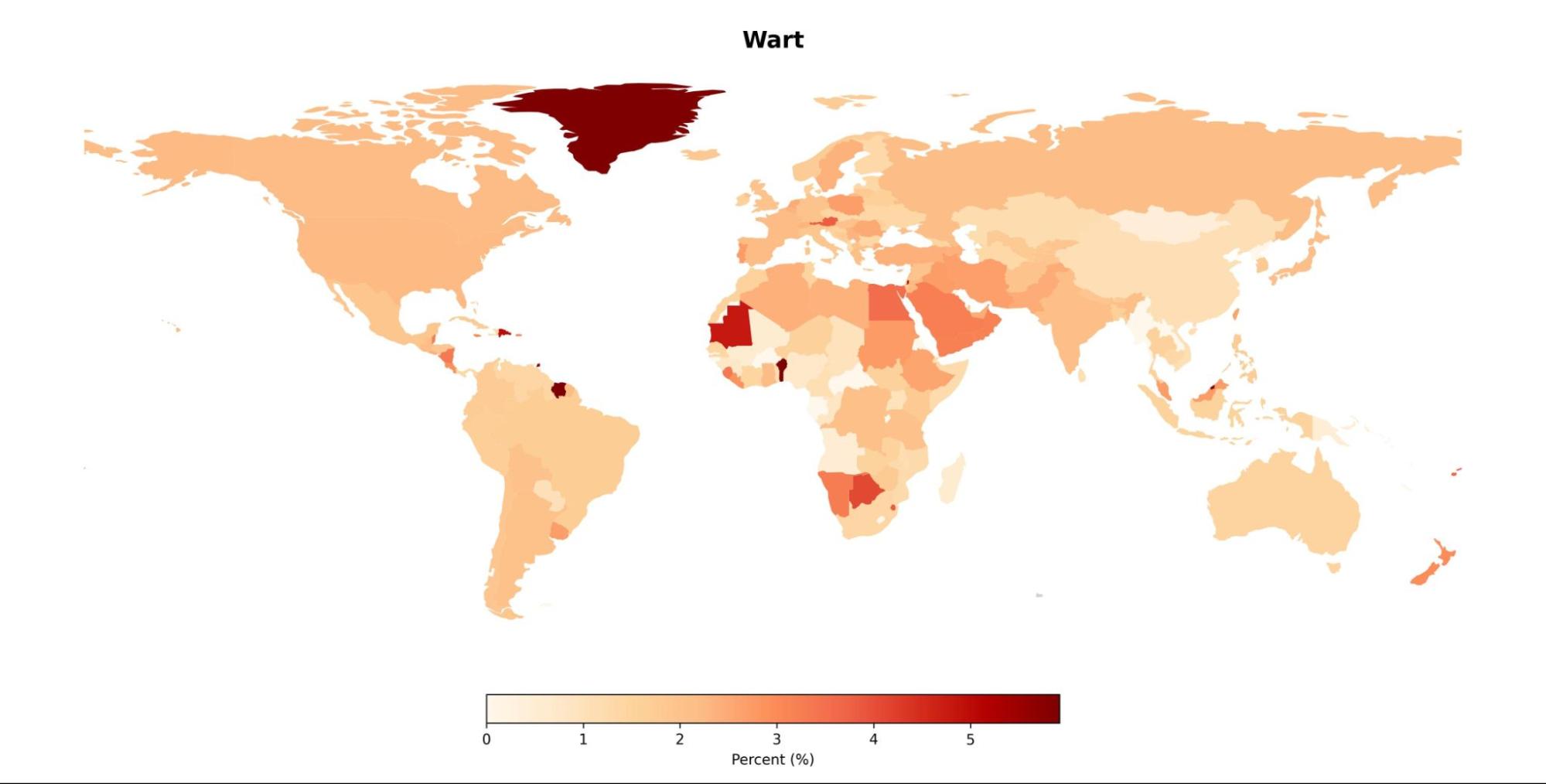

**Table S1 - Demographics of the SNU test Dataset and Top-1/3 Accuracies of the Algorithm**

|  |  | Binary |  | Multi-Class |  |
| --- | --- | --- | --- | --- | --- |
| Disease | No. of  Images | TOP-1 | TOP-3 | TOP-1 | TOP-3 |
| Abscess | 0 | 100.0% | 100.0% | 0.0% | 100.0% |
| Acne | 2 | 100.0% | 100.0% | 50.0% | 75.0% |
| Actinic keratosis | 4 | 100.0% | 0.0% | 100.0% | 100.0% |
| Acute generalized exanthematous pustulosis | 0 | 100.0% | 100.0% | 0.0% | 0.0% |
| Alopecia areata | 1 | 100.0% | 100.0% | 100.0% | 100.0% |
| Amyloidosis | 2 | 100.0% | 100.0% | 66.7% | 100.0% |
| Angiofibroma | 1 | 100.0% | 100.0% | 100.0% | 100.0% |
| Angiokeratoma | 1 | 100.0% | 100.0% | 100.0% | 100.0% |
| Basal cell carcinoma | 7 | 87.5% | 100.0% | 87.5% | 100.0% |
| Becker nevus | 0 | 100.0% | 100.0% | 0.0% | 100.0% |
| Blue nevus | 1 | 100.0% | 0.0% | 100.0% | 100.0% |
| Intraepithelial carcinoma (Bowen disease) | 3 | 57.1% | 85.7% | 42.9% | 57.1% |
| Cellulitis | 1 | 75.0% | 75.0% | 25.0% | 25.0% |
| Confluent reticulated papillomatosis | 3 | 100.0% | 100.0% | 100.0% | 100.0% |
| Congenital nevus | 1 | 100.0% | 100.0% | 100.0% | 100.0% |
| Contact dermatitis | 1 | 100.0% | 100.0% | 100.0% | 100.0% |
| Dermatofibroma | 1 | 100.0% | 100.0% | 100.0% | 100.0% |
| Drug eruption | 1 | 100.0% | 100.0% | 100.0% | 100.0% |
| Eczema herpeticum | 4 | 100.0% | 100.0% | 66.7% | 83.3% |
| Epidermal cyst | 1 | 100.0% | 100.0% | 100.0% | 100.0% |
| Epidermal nevus | 1 | 100.0% | 100.0% | 100.0% | 100.0% |
| Erythema annulare centrifugum | 1 | 100.0% | 100.0% | 100.0% | 100.0% |
| Erythema multiforme | 2 | 100.0% | 100.0% | 100.0% | 100.0% |
| Exfoliative dermatitis | 1 | 100.0% | 100.0% | 100.0% | 100.0% |
| Folliculitis | 1 | 100.0% | 100.0% | 25.0% | 50.0% |
| Furuncle | 0 | 100.0% | 100.0% | 0.0% | 25.0% |
| Granuloma annulare | 1 | 100.0% | 100.0% | 100.0% | 100.0% |
| Hand eczema | 0 | 100.0% | 100.0% | 0.0% | 0.0% |
| Hemangioma | 1 | 100.0% | 100.0% | 100.0% | 100.0% |
| Herpes simplex | 5 | 100.0% | 83.3% | 83.3% | 100.0% |
| Herpes zoster | 13 | 100.0% | 94.4% | 72.2% | 72.2% |
| Impetigo | 2 | 100.0% | 100.0% | 40.0% | 40.0% |
| Inflammed cyst | 0 | 100.0% | 50.0% | 0.0% | 50.0% |
| Insect bite | 1 | 66.7% | 66.7% | 33.3% | 100.0% |
| Juvenile xanthogranuloma | 2 | 100.0% | 100.0% | 100.0% | 100.0% |
| Keratoacanthoma | 1 | 33.3% | 100.0% | 33.3% | 33.3% |
| Lentigo | 2 | 100.0% | 100.0% | 100.0% | 100.0% |
| Lichen planus | 1 | 100.0% | 100.0% | 100.0% | 100.0% |
| Lichen simplex chronicus | 0 | 100.0% | 100.0% | 0.0% | 0.0% |
| Lupus erythematosus | 1 | 100.0% | 100.0% | 50.0% | 50.0% |
| Lymphangioma | 0 | 100.0% | 0.0% | 0.0% | 0.0% |
| Malignant melanoma | 7 | 50.0% | 71.4% | 50.0% | 71.4% |
| Melanocytic nevus | 2 | 66.7% | 66.7% | 66.7% | 100.0% |
| Morphea | 0 | 100.0% | 100.0% | 0.0% | 0.0% |
| Neurofibroma | 0 | 100.0% | 100.0% | 0.0% | 100.0% |
| Neurofibromatosis | 0 | 100.0% | 100.0% | 0.0% | 100.0% |
| Nevus spilus | 0 | 100.0% | 100.0% | 0.0% | 100.0% |
| Nummular eczema | 1 | 100.0% | 100.0% | 50.0% | 100.0% |
| onychomycosis | 6 | 100.0% | 100.0% | 75.0% | 87.5% |
| Orgarnoid nevus | 1 | 100.0% | 100.0% | 100.0% | 100.0% |
| Palmoplantar pustulosis | 1 | 100.0% | 100.0% | 100.0% | 100.0% |
| Paronychia | 1 | 100.0% | 100.0% | 25.0% | 100.0% |
| Pompholyx | 0 | 100.0% | 100.0% | 0.0% | 100.0% |
| Prurigo pigmentosa | 4 | 100.0% | 100.0% | 100.0% | 100.0% |
| Psoriasis | 2 | 100.0% | 100.0% | 100.0% | 100.0% |
| Pustular psoriasis | 0 | 100.0% | 100.0% | 0.0% | 100.0% |
| Pyoderma gangrenosum | 1 | 100.0% | 0.0% | 100.0% | 100.0% |
| Pyogenic granuloma | 1 | 100.0% | 100.0% | 100.0% | 100.0% |
| Sebaceus hyperplasia | 1 | 100.0% | 0.0% | 50.0% | 100.0% |
| Seborrheic dermatitis | 1 | 100.0% | 100.0% | 100.0% | 100.0% |
| Seborrheic keratosis | 4 | 100.0% | 50.0% | 100.0% | 100.0% |
| Skin tag | 1 | 100.0% | 100.0% | 100.0% | 100.0% |
| Squamous cell carcinoma | 4 | 87.5% | 100.0% | 50.0% | 100.0% |
| Staphylococcal scalded skin syndrome | 2 | 100.0% | 100.0% | 100.0% | 100.0% |
| Steatocystoma multiplex | 1 | 100.0% | 100.0% | 100.0% | 100.0% |
| Subungual hematoma | 0 | 100.0% | 100.0% | 0.0% | 100.0% |
| Tinea corporis | 4 | 100.0% | 100.0% | 44.4% | 66.7% |
| Tinea cruris | 2 | 100.0% | 100.0% | 66.7% | 66.7% |
| Tinea faciei | 3 | 100.0% | 100.0% | 60.0% | 100.0% |
| Tinea pedis | 5 | 100.0% | 100.0% | 62.5% | 87.5% |
| Tinea versicolor | 4 | 100.0% | 100.0% | 57.1% | 85.7% |
| Urticaria | 1 | 100.0% | 100.0% | 100.0% | 100.0% |
| Urticaria pigmentosa | 2 | 100.0% | 100.0% | 66.7% | 100.0% |
| Varicella | 6 | 100.0% | 100.0% | 60.0% | 80.0% |
| Vasculitis | 3 | 100.0% | 100.0% | 100.0% | 100.0% |
| Verruca plana | 1 | 100.0% | 100.0% | 100.0% | 100.0% |
| Vitiligo | 1 | 100.0% | 100.0% | 100.0% | 100.0% |
| Wart | 0 | 100.0% | 100.0% | 0.0% | 50.0% |
| Xanthelasma | 1 | 100.0% | 100.0% | 100.0% | 100.0% |
| Xerotic eczema | 0 | 100.0% | 100.0% | 0.0% | 0.0% |
| *Total* | *145* | ** 92.9%*  *** 96.5%* | ** 90.4%*  *** 90.5%* | ** 60.4%*  *** 63.3%* | ** 80.8%*  *** 83.2%* |

* macro-averaged, ** micro-averaged; Binary class accuracy refers to the correctness of predictions in determining whether a case is cancerous or not, while multi-class accuracy measures the correctness of predictions in identifying the specific disease name accurately.

**Table S2 - Results of the Reader Test on a Global Scale Using the SNU Dataset**

| Location | Sensitivity | Specificity | Accuracy |
| --- | --- | --- | --- |
| Africa | 43.8% (174/397) | 77.7% (468/2,096) | 72.3% (1,802/2,493) |
| Asia | 51.7% (2,212/4,277) | 85.9% (2,998/21,330) | 80.2% (20,544/25,607) |
| Europe | 59.2% (2,598/4,389) | 83.9% (3,436/21,379) | 79.7% (20,541/25,768) |
| North America | 64.0% (310/484) | 80.4% (447/2,284) | 77.6% (2,147/2,768) |
| Oceania | 62.6% (57/91) | 82.5% (76/435) | 79.1% (416/526) |
| South America | 50.6% (335/662) | 83.6% (533/3,242) | 78.0% (3,044/3,904) |
| *Total* | *55.2% (5,686/10,300)* | *84.3% (7,958/50,766)* | *79.4% (48,494/61,066)* |
| Afghanistan | 16.7% (1/6) | 52.2% (11/23) | 44.8% (13/29) |
| Albania | 90.0% (9/10) | 61.1% (7/18) | 71.4% (20/28) |
| Algeria | 56.3% (71/126) | 67.5% (215/661) | 65.7% (517/787) |
| Angola | 0.0% (0/1) | 55.6% (4/9) | 50.0% (5/10) |
| Argentina | 25.6% (11/43) | 82.7% (39/225) | 73.5% (197/268) |
| Armenia | 58.5% (24/41) | 89.4% (23/216) | 84.4% (217/257) |
| Australia | 63.2% (24/38) | 71.6% (48/169) | 70.0% (145/207) |
| Austria | 78.4% (40/51) | 73.5% (65/245) | 74.3% (220/296) |
| Azerbaijan | 71.4% (5/7) | 53.4% (27/58) | 55.4% (36/65) |
| Bangladesh | 23.8% (19/80) | 89.0% (36/326) | 76.1% (309/406) |
| Belarus | 69.7% (53/76) | 84.6% (58/376) | 82.1% (371/452) |
| Belgium | 50.0% (26/52) | 81.6% (46/250) | 76.2% (230/302) |
| Belize | 0.0% (0/7) | 82.6% (4/23) | 63.3% (19/30) |
| Bolivia, Plurinational State of | 100.0% (2/2) | 36.4% (7/11) | 46.2% (6/13) |
| Bosnia and Herzegovina | 61.9% (13/21) | 82.5% (18/103) | 79.0% (98/124) |
| Brazil | 50.3% (245/487) | 84.9% (351/2,320) | 78.9% (2,214/2,807) |
| Bulgaria | 63.5% (40/63) | 83.4% (58/350) | 80.4% (332/413) |
| Burkina Faso | 100.0% (5/5) | 18.2% (9/11) | 43.8% (7/16) |
| Cambodia | 100.0% (1/1) | 75.0% (3/12) | 76.9% (10/13) |
| Cameroon | 13.3% (2/15) | 92.1% (6/76) | 79.1% (72/91) |
| Canada | 72.9% (78/107) | 79.8% (102/505) | 78.6% (481/612) |
| Central African Republic | 100.0% (1/1) | 0.0% (9/9) | 10.0% (1/10) |
| Chad | 100.0% (3/3) | 10.0% (9/10) | 30.8% (4/13) |
| Chile | 88.9% (16/18) | 62.9% (36/97) | 67.0% (77/115) |
| China | 32.3% (10/31) | 86.9% (23/175) | 78.6% (162/206) |
| Colombia | 75.0% (48/64) | 83.2% (61/363) | 82.0% (350/427) |
| Congo, The Democratic Republic of the | 66.7% (4/6) | 57.7% (11/26) | 59.4% (19/32) |
| Côte d'Ivoire | 66.7% (2/3) | 37.9% (18/29) | 40.6% (13/32) |
| Croatia | 59.4% (41/69) | 89.4% (32/303) | 83.9% (312/372) |
| Cuba | 50.0% (4/8) | 78.2% (12/55) | 74.6% (47/63) |
| Cyprus | 75.0% (3/4) | 56.7% (13/30) | 58.8% (20/34) |
| Czechia | 48.5% (95/196) | 83.9% (154/957) | 77.9% (898/1,153) |
| Denmark | 42.9% (12/28) | 88.7% (11/97) | 78.4% (98/125) |
| Ecuador | 54.5% (6/11) | 72.7% (12/44) | 69.1% (38/55) |
| Egypt | 47.8% (11/23) | 70.8% (28/96) | 66.4% (79/119) |
| El Salvador | 50.0% (5/10) | 86.8% (5/38) | 79.2% (38/48) |
| Estonia | 66.7% (4/6) | 96.4% (1/28) | 91.2% (31/34) |
| Ethiopia | 66.7% (2/3) | 80.0% (5/25) | 78.6% (22/28) |
| Fiji | 0.0% (0/2) | 100.0% (0/9) | 81.8% (9/11) |
| Finland | 62.9% (22/35) | 85.9% (21/149) | 81.5% (150/184) |
| France | 64.5% (225/349) | 82.0% (303/1,685) | 79.0% (1,607/2,034) |
| Gabon | 0.0% (0/40) | 95.0% (10/200) | 79.2% (190/240) |
| Gambia | 0.0% (0/1) | 100.0% (0/19) | 95.0% (19/20) |
| Georgia | 88.9% (32/36) | 17.9% (147/179) | 29.8% (64/215) |
| Germany | 65.8% (578/878) | 83.3% (707/4,236) | 80.3% (4,107/5,114) |
| Ghana | 100.0% (1/1) | 75.0% (1/4) | 80.0% (4/5) |
| Greece | 36.8% (7/19) | 85.4% (15/103) | 77.9% (95/122) |
| Guadeloupe | 100.0% (1/1) | 40.0% (3/5) | 50.0% (3/6) |
| Haiti | 33.3% (1/3) | 58.3% (5/12) | 53.3% (8/15) |
| Honduras | 0.0% (0/2) | 100.0% (0/8) | 80.0% (8/10) |
| Hong Kong | 83.3% (5/6) | 71.4% (8/28) | 73.5% (25/34) |
| Hungary | 53.3% (130/244) | 85.4% (187/1,282) | 80.3% (1,225/1,526) |
| Iceland | - | 85.7% (1/7) | 85.7% (6/7) |
| India | 29.7% (11/37) | 82.8% (27/157) | 72.7% (141/194) |
| Indonesia | 33.3% (57/171) | 89.6% (92/885) | 80.5% (850/1,056) |
| Iran, Islamic Republic of | 31.1% (32/103) | 88.4% (59/508) | 78.7% (481/611) |
| Iraq | 35.3% (49/139) | 81.9% (135/746) | 74.6% (660/885) |
| Ireland | 28.6% (2/7) | 86.7% (6/45) | 78.8% (41/52) |
| Israel | 78.6% (11/14) | 76.6% (11/47) | 77.0% (47/61) |
| Italy | 61.3% (373/608) | 84.6% (462/3,005) | 80.7% (2,916/3,613) |
| Jamaica | 0.0% (0/2) | 88.9% (1/9) | 72.7% (8/11) |
| Japan | 50.0% (32/64) | 83.5% (47/285) | 77.4% (270/349) |
| Jordan | 66.7% (2/3) | 94.4% (2/36) | 92.3% (36/39) |
| Kazakhstan | 65.0% (13/20) | 79.6% (20/98) | 77.1% (91/118) |
| Kenya | 0.0% (0/1) | - | 0.0% (0/1) |
| Korea, Republic of | 57.0% (1,689/2,965) | 87.2% (1,895/14,805) | 82.2% (14,599/17,770) |
| Kuwait | 0.0% (0/4) | 100.0% (0/9) | 69.2% (9/13) |
| Kyrgyzstan | 50.0% (2/4) | 76.2% (5/21) | 72.0% (18/25) |
| Latvia | 100.0% (1/1) | 66.7% (3/9) | 70.0% (7/10) |
| Lebanon | 28.6% (2/7) | 95.5% (1/22) | 79.3% (23/29) |
| Libya | 27.3% (3/11) | 81.4% (11/59) | 72.9% (51/70) |
| Lithuania | 50.0% (3/6) | 77.8% (6/27) | 72.7% (24/33) |
| Luxembourg | 100.0% (2/2) | 71.4% (4/14) | 75.0% (12/16) |
| Madagascar | - | 0.0% (5/5) | 0.0% (0/5) |
| Malaysia | 57.1% (12/21) | 79.4% (21/102) | 75.6% (93/123) |
| Maldives | 0.0% (0/1) | - | 0.0% (0/1) |
| Mali | 0.0% (0/3) | 94.4% (1/18) | 81.0% (17/21) |
| Mauritania | - | 0.0% (10/10) | 0.0% (0/10) |
| Mexico | 37.5% (9/24) | 80.5% (24/123) | 73.5% (108/147) |
| Moldova, Republic of | 50.0% (1/2) | 73.3% (4/15) | 70.6% (12/17) |
| Monaco | 100.0% (1/1) | - | 100.0% (1/1) |
| Morocco | 50.0% (36/72) | 89.4% (39/367) | 82.9% (364/439) |
| Mozambique | 66.7% (2/3) | 53.8% (6/13) | 56.3% (9/16) |
| Myanmar | 0.0% (0/14) | 100.0% (0/66) | 82.5% (66/80) |
| Nepal | 87.5% (7/8) | 96.3% (1/27) | 94.3% (33/35) |
| Netherlands | 73.5% (25/34) | 76.0% (35/146) | 75.6% (136/180) |
| New Zealand | 76.7% (33/43) | 89.6% (25/241) | 87.7% (249/284) |
| Niger | - | 50.0% (3/6) | 50.0% (3/6) |
| Nigeria | 50.0% (2/4) | 92.3% (2/26) | 86.7% (26/30) |
| North Macedonia | 100.0% (1/1) | 81.8% (2/11) | 83.3% (10/12) |
| Norway | 0.0% (0/1) | 90.9% (1/11) | 83.3% (10/12) |
| Oman | - | 100.0% (0/2) | 100.0% (2/2) |
| Pakistan | 37.1% (13/35) | 85.9% (30/213) | 79.0% (196/248) |
| Panama | 0.0% (0/7) | 94.4% (2/36) | 79.1% (34/43) |
| Papua New Guinea | 0.0% (0/8) | 92.9% (1/14) | 59.1% (13/22) |
| Paraguay | 0.0% (0/4) | 92.3% (1/13) | 70.6% (12/17) |
| Peru | 12.5% (2/16) | 75.4% (15/61) | 62.3% (48/77) |
| Philippines | 38.5% (5/13) | 91.4% (5/58) | 81.7% (58/71) |
| Poland | 58.1% (155/267) | 86.2% (173/1,251) | 81.2% (1,233/1,518) |
| Portugal | 58.5% (31/53) | 86.8% (43/325) | 82.8% (313/378) |
| Qatar | 0.0% (0/41) | 100.0% (0/200) | 83.0% (200/241) |
| Réunion | 0.0% (0/2) | 100.0% (0/4) | 66.7% (4/6) |
| Romania | 50.0% (54/108) | 84.4% (73/468) | 78.0% (449/576) |
| Russian Federation | 45.7% (148/324) | 85.0% (235/1,570) | 78.3% (1,483/1,894) |
| Rwanda | 0.0% (0/2) | 100.0% (0/12) | 85.7% (12/14) |
| Sao Tome and Principe | 100.0% (1/1) | 28.6% (5/7) | 37.5% (3/8) |
| Saudi Arabia | 70.0% (14/20) | 80.2% (23/116) | 78.7% (107/136) |
| Senegal | 66.7% (2/3) | 100.0% (0/7) | 90.0% (9/10) |
| Serbia | 62.5% (20/32) | 52.5% (57/120) | 54.6% (83/152) |
| Seychelles | 100.0% (1/1) | 11.1% (8/9) | 20.0% (2/10) |
| Singapore | 0.0% (0/1) | - | 0.0% (0/1) |
| Slovakia | 55.1% (38/69) | 71.7% (100/353) | 69.0% (291/422) |
| Slovenia | 85.7% (6/7) | 76.3% (14/59) | 77.3% (51/66) |
| South Africa | 56.3% (9/16) | 71.4% (22/77) | 68.8% (64/93) |
| Spain | 62.8% (241/384) | 86.4% (251/1,848) | 82.3% (1,838/2,232) |
| Sri Lanka | - | 100.0% (0/3) | 100.0% (3/3) |
| Sudan | 9.1% (1/11) | 83.8% (13/80) | 74.7% (68/91) |
| Sweden | 88.9% (8/9) | 61.4% (17/44) | 66.0% (35/53) |
| Switzerland | 50.0% (20/40) | 87.6% (28/226) | 82.0% (218/266) |
| Syrian Arab Republic | 6.3% (1/16) | 95.9% (2/49) | 73.8% (48/65) |
| Taiwan, Province of China | 34.5% (10/29) | 88.6% (20/176) | 81.0% (166/205) |
| Tajikistan | 30.0% (6/20) | 76.7% (17/73) | 66.7% (62/93) |
| Tanzania, United Republic of | 100.0% (1/1) | 33.3% (4/6) | 42.9% (3/7) |
| Thailand | 28.6% (4/14) | 84.2% (9/57) | 73.2% (52/71) |
| Togo | 0.0% (0/1) | 100.0% (0/3) | 75.0% (3/4) |
| Tunisia | 37.8% (14/37) | 93.4% (14/212) | 85.1% (212/249) |
| Türkiye | 57.5% (46/80) | 85.3% (50/339) | 80.0% (335/419) |
| Turkmenistan | 40.0% (2/5) | 75.8% (16/66) | 73.2% (52/71) |
| Ukraine | 7.0% (9/128) | 96.0% (26/651) | 81.4% (634/779) |
| United Arab Emirates | 28.9% (11/38) | 90.6% (21/223) | 81.6% (213/261) |
| United Kingdom | 78.8% (164/208) | 78.6% (212/992) | 78.7% (944/1,200) |
| United States | 67.7% (212/313) | 80.3% (289/1,470) | 78.1% (1,393/1,783) |
| United States Minor Outlying Islands | - | 0.0% (2/2) | 0.0% (0/2) |
| Uruguay | 30.0% (3/10) | 84.0% (8/50) | 75.0% (45/60) |
| Uzbekistan | 30.4% (21/69) | 87.4% (43/340) | 77.8% (318/409) |
| Venezuela, Bolivarian Republic of | 28.6% (2/7) | 94.8% (3/58) | 87.7% (57/65) |
| Viet Nam | 20.8% (5/24) | 91.1% (11/124) | 79.7% (118/148) |
| Yemen | 64.7% (55/85) | 66.5% (144/430) | 66.2% (341/515) |

**Table S3 - Results of the Reader Test on a Global Scale Using the Onychomycosis Dataset**

| Location | Sensitivity | Specificity | Accuracy |
| --- | --- | --- | --- |
| Africa | 51.4% (1,836/3,570) | 50.4% (2,260/4,553) | 50.8% (4,129/8,123) |
| Asia | 66.4% (8,685/13,082) | 50.6% (7,691/15,582) | 57.8% (16,576/28,664) |
| Europe | 36.7% (3,827/10,415) | 67.4% (4,046/12,413) | 53.4% (12,194/22,828) |
| North America | 48.0% (1,009/2,100) | 60.1% (1,034/2,590) | 54.7% (2,565/4,690) |
| Oceania | 40.0% (108/270) | 64.4% (114/320) | 53.2% (314/590) |
| South America | 43.1% (985/2,285) | 61.3% (1,066/2,751) | 53.0% (2,670/5,036) |
| *Total* | *51.9% (16,450/31,722)* | *57.6% (16,211/38,209)* | *55.0% (38,448/69,931)* |
| Afghanistan | 52.6% (80/152) | 46.7% (90/169) | 49.5% (159/321) |
| Albania | 80.7% (138/171) | 25.0% (156/208) | 50.1% (190/379) |
| Algeria | 48.9% (272/556) | 52.8% (339/718) | 51.1% (651/1,274) |
| Angola | 62.9% (83/132) | 39.7% (105/174) | 49.7% (152/306) |
| Argentina | 42.7% (70/164) | 69.9% (49/163) | 56.3% (184/327) |
| Armenia | 33.3% (2/6) | 66.7% (2/6) | 50.0% (6/12) |
| Aruba | - | 100.0% (0/1) | 100.0% (1/1) |
| Australia | 36.7% (58/158) | 66.3% (54/160) | 51.6% (164/318) |
| Austria | 31.3% (46/147) | 69.8% (51/169) | 51.9% (164/316) |
| Azerbaijan | 64.0% (48/75) | 40.0% (45/75) | 52.0% (78/150) |
| Bahrain | 90.0% (9/10) | 66.7% (3/9) | 78.9% (15/19) |
| Bangladesh | 45.6% (123/270) | 50.9% (141/287) | 48.3% (269/557) |
| Barbados | 0.0% (0/4) | 100.0% (0/6) | 60.0% (6/10) |
| Belarus | 29.4% (94/320) | 74.4% (90/351) | 52.9% (355/671) |
| Belgium | 55.4% (72/130) | 55.3% (80/179) | 55.3% (171/309) |
| Belize | 0.0% (0/7) | 100.0% (0/3) | 30.0% (3/10) |
| Benin | 41.2% (7/17) | 43.8% (18/32) | 42.9% (21/49) |
| Bhutan | 0.0% (0/5) | 88.9% (1/9) | 57.1% (8/14) |
| Bolivia, Plurinational State of | 74.2% (46/62) | 27.3% (80/110) | 44.2% (76/172) |
| Bosnia and Herzegovina | 29.9% (32/107) | 68.5% (46/146) | 52.2% (132/253) |
| Botswana | 100.0% (1/1) | - | 100.0% (1/1) |
| Brazil | 38.0% (494/1,301) | 66.3% (531/1,576) | 53.5% (1,539/2,877) |
| Bulgaria | 27.9% (19/68) | 69.5% (25/82) | 50.7% (76/150) |
| Burkina Faso | 14.0% (6/43) | 66.2% (26/77) | 47.5% (57/120) |
| Burundi | 58.8% (10/17) | 45.8% (13/24) | 51.2% (21/41) |
| Cabo Verde | 40.0% (2/5) | 100.0% (0/5) | 70.0% (7/10) |
| Cambodia | 48.0% (12/25) | 44.1% (19/34) | 45.8% (27/59) |
| Cameroon | 46.1% (47/102) | 43.1% (70/123) | 44.4% (100/225) |
| Canada | 43.4% (134/309) | 63.3% (135/368) | 54.2% (367/677) |
| Central African Republic | 80.0% (8/10) | 5.9% (16/17) | 33.3% (9/27) |
| Chad | 78.7% (74/94) | 29.8% (85/121) | 51.2% (110/215) |
| Chile | 51.9% (80/154) | 52.9% (113/240) | 52.5% (207/394) |
| China | 45.4% (74/163) | 59.9% (69/172) | 52.8% (177/335) |
| Colombia | 38.9% (79/203) | 59.9% (85/212) | 49.6% (206/415) |
| Congo | 64.1% (25/39) | 37.7% (33/53) | 48.9% (45/92) |
| Congo, The Democratic Republic of the | 59.6% (165/277) | 43.8% (180/320) | 51.1% (305/597) |
| Costa Rica | 75.0% (9/12) | 30.0% (7/10) | 54.5% (12/22) |
| Côte d'Ivoire | 47.1% (73/155) | 55.6% (84/189) | 51.7% (178/344) |
| Croatia | 13.2% (31/234) | 84.6% (40/259) | 50.7% (250/493) |
| Cuba | 46.6% (75/161) | 47.9% (111/213) | 47.3% (177/374) |
| Cyprus | 50.0% (15/30) | 64.9% (13/37) | 58.2% (39/67) |
| Czechia | 30.5% (100/328) | 69.7% (111/366) | 51.2% (355/694) |
| Denmark | 63.4% (26/41) | 50.8% (30/61) | 55.9% (57/102) |
| Djibouti | 100.0% (4/4) | 0.0% (6/6) | 40.0% (4/10) |
| Dominican Republic | 23.1% (3/13) | 91.7% (1/12) | 56.0% (14/25) |
| Ecuador | 58.7% (37/63) | 55.4% (29/65) | 57.0% (73/128) |
| Egypt | 45.1% (184/408) | 51.2% (254/520) | 48.5% (450/928) |
| El Salvador | 50.0% (11/22) | 60.9% (9/23) | 55.6% (25/45) |
| Equatorial Guinea | 100.0% (6/6) | 100.0% (0/4) | 100.0% (10/10) |
| Estonia | 73.0% (27/37) | 28.3% (38/53) | 46.7% (42/90) |
| Ethiopia | 44.8% (64/143) | 59.9% (81/202) | 53.6% (185/345) |
| Fiji | 0.0% (0/3) | 100.0% (0/4) | 57.1% (4/7) |
| Finland | 10.3% (3/29) | 97.6% (1/41) | 61.4% (43/70) |
| France | 36.5% (184/504) | 65.4% (226/654) | 52.8% (612/1,158) |
| French Guiana | 0.0% (0/1) | - | 0.0% (0/1) |
| Gabon | 100.0% (10/10) | 0.0% (18/18) | 35.7% (10/28) |
| Gambia | 100.0% (4/4) | 0.0% (6/6) | 40.0% (4/10) |
| Georgia | 53.2% (33/62) | 73.2% (19/71) | 63.9% (85/133) |
| Germany | 29.8% (407/1,367) | 76.0% (402/1,674) | 55.2% (1,679/3,041) |
| Ghana | 44.4% (16/36) | 34.6% (34/52) | 38.6% (34/88) |
| Greece | 37.1% (49/132) | 59.9% (79/197) | 50.8% (167/329) |
| Guadeloupe | 100.0% (1/1) | 0.0% (1/1) | 50.0% (1/2) |
| Guatemala | 41.2% (7/17) | 82.4% (3/17) | 61.8% (21/34) |
| Guinea | 76.7% (23/30) | 21.7% (36/46) | 43.4% (33/76) |
| Guinea-Bissau | 12.5% (1/8) | 72.7% (3/11) | 47.4% (9/19) |
| Guyana | 38.9% (7/18) | 68.8% (5/16) | 52.9% (18/34) |
| Haiti | 56.0% (28/50) | 50.0% (27/54) | 52.9% (55/104) |
| Honduras | 55.6% (10/18) | 70.8% (7/24) | 64.3% (27/42) |
| Hong Kong | 58.3% (7/12) | 53.3% (7/15) | 55.6% (15/27) |
| Hungary | 73.3% (508/693) | 40.6% (514/866) | 55.2% (860/1,559) |
| India | 33.8% (48/142) | 67.5% (63/194) | 53.3% (179/336) |
| Indonesia | 46.8% (161/344) | 52.9% (206/437) | 50.2% (392/781) |
| Iran, Islamic Republic of | 37.4% (257/687) | 64.4% (278/781) | 51.8% (760/1,468) |
| Iraq | 55.1% (407/738) | 51.8% (416/863) | 53.3% (854/1,601) |
| Ireland | 41.8% (23/55) | 63.1% (24/65) | 53.3% (64/120) |
| Israel | 59.7% (46/77) | 49.4% (43/85) | 54.3% (88/162) |
| Italy | 31.8% (481/1,513) | 70.8% (494/1,689) | 52.3% (1,676/3,202) |
| Jamaica | 23.5% (4/17) | 62.5% (9/24) | 46.3% (19/41) |
| Japan | 61.4% (137/223) | 53.2% (116/248) | 57.1% (269/471) |
| Jordan | 40.9% (27/66) | 65.5% (19/55) | 52.1% (63/121) |
| Kazakhstan | 42.9% (33/77) | 69.9% (28/93) | 57.6% (98/170) |
| Kenya | 45.5% (20/44) | 56.8% (19/44) | 51.1% (45/88) |
| Korea, Republic of | 80.2% (5,809/7,243) | 45.2% (4,673/8,533) | 61.3% (9,669/15,776) |
| Kuwait | 15.0% (3/20) | 84.6% (2/13) | 42.4% (14/33) |
| Kyrgyzstan | 36.6% (15/41) | 70.5% (13/44) | 54.1% (46/85) |
| Latvia | 66.7% (16/24) | 40.0% (21/35) | 50.8% (30/59) |
| Lebanon | 77.4% (24/31) | 21.2% (41/52) | 42.2% (35/83) |
| Lesotho | 100.0% (1/1) | - | 100.0% (1/1) |
| Liberia | - | 0.0% (1/1) | 0.0% (0/1) |
| Libya | 42.8% (71/166) | 59.7% (73/181) | 51.6% (179/347) |
| Lithuania | 77.8% (7/9) | 47.1% (9/17) | 57.7% (15/26) |
| Luxembourg | 0.0% (0/9) | 100.0% (0/22) | 71.0% (22/31) |
| Macao | 75.0% (6/8) | 16.7% (10/12) | 40.0% (8/20) |
| Madagascar | 80.0% (12/15) | 31.8% (15/22) | 51.4% (19/37) |
| Malawi | 100.0% (6/6) | 0.0% (7/7) | 46.2% (6/13) |
| Malaysia | 58.6% (17/29) | 55.6% (20/45) | 56.8% (42/74) |
| Maldives | 18.2% (2/11) | 80.0% (2/10) | 47.6% (10/21) |
| Mali | 70.1% (47/67) | 23.8% (61/80) | 44.9% (66/147) |
| Malta | 16.7% (1/6) | 57.1% (3/7) | 38.5% (5/13) |
| Mauritania | 75.9% (22/29) | 42.4% (19/33) | 58.1% (36/62) |
| Mayotte | 100.0% (2/2) | 0.0% (2/2) | 50.0% (2/4) |
| Mexico | 56.5% (166/294) | 55.1% (176/392) | 55.7% (382/686) |
| Moldova, Republic of | 3.6% (2/55) | 72.9% (16/59) | 39.5% (45/114) |
| Mongolia | 0.0% (0/4) | 100.0% (0/3) | 42.9% (3/7) |
| Montenegro | 18.2% (4/22) | 63.2% (7/19) | 39.0% (16/41) |
| Morocco | 43.0% (170/395) | 70.3% (148/499) | 58.3% (521/894) |
| Mozambique | 71.8% (74/103) | 33.3% (94/141) | 49.6% (121/244) |
| Myanmar | 0.0% (0/16) | 88.2% (2/17) | 45.5% (15/33) |
| Namibia | 0.0% (0/1) | 0.0% (2/2) | 0.0% (0/3) |
| Nepal | 69.4% (25/36) | 34.9% (28/43) | 50.6% (40/79) |
| Netherlands | 60.3% (117/194) | 49.5% (97/192) | 54.9% (212/386) |
| New Caledonia | 28.6% (2/7) | 76.9% (3/13) | 60.0% (12/20) |
| New Zealand | 36.4% (24/66) | 75.6% (22/90) | 59.0% (92/156) |
| Nicaragua | 51.6% (16/31) | 65.9% (14/41) | 59.7% (43/72) |
| Niger | 60.0% (15/25) | 27.3% (32/44) | 39.1% (27/69) |
| Nigeria | 60.3% (38/63) | 37.6% (53/85) | 47.3% (70/148) |
| North Macedonia | 64.5% (20/31) | 53.8% (12/26) | 59.6% (34/57) |
| Norway | 42.9% (6/14) | 53.8% (6/13) | 48.1% (13/27) |
| Oman | 55.6% (5/9) | 63.6% (4/11) | 60.0% (12/20) |
| Pakistan | 33.0% (121/367) | 65.7% (159/463) | 51.2% (425/830) |
| Palestine, State of | 81.8% (18/22) | 68.4% (6/19) | 75.6% (31/41) |
| Panama | 38.9% (7/18) | 81.5% (5/27) | 64.4% (29/45) |
| Papua New Guinea | 73.7% (14/19) | 26.5% (25/34) | 43.4% (23/53) |
| Paraguay | 33.3% (3/9) | 80.0% (1/5) | 50.0% (7/14) |
| Peru | 55.9% (76/136) | 53.4% (68/146) | 54.6% (154/282) |
| Philippines | 73.3% (22/30) | 39.5% (23/38) | 54.4% (37/68) |
| Poland | 20.6% (109/530) | 80.3% (132/670) | 53.9% (647/1,200) |
| Portugal | 32.1% (68/212) | 59.7% (93/231) | 46.5% (206/443) |
| Puerto Rico | 25.0% (4/16) | 71.4% (6/21) | 51.4% (19/37) |
| Qatar | 0.0% (0/15) | 95.2% (1/21) | 55.6% (20/36) |
| Réunion | 50.0% (5/10) | 50.0% (7/14) | 50.0% (12/24) |
| Romania | 40.2% (131/326) | 62.6% (148/396) | 52.5% (379/722) |
| Russian Federation | 27.3% (327/1,197) | 72.2% (384/1,383) | 51.4% (1,326/2,580) |
| Rwanda | 37.5% (3/8) | 50.0% (5/10) | 44.4% (8/18) |
| Saint Kitts and Nevis | 85.7% (6/7) | 38.5% (8/13) | 55.0% (11/20) |
| Samoa | 100.0% (4/4) | 33.3% (4/6) | 60.0% (6/10) |
| Sao Tome and Principe | 42.9% (3/7) | 64.3% (5/14) | 57.1% (12/21) |
| Saudi Arabia | 67.9% (114/168) | 58.5% (86/207) | 62.7% (235/375) |
| Senegal | 53.8% (21/39) | 44.4% (15/27) | 50.0% (33/66) |
| Serbia | 70.7% (104/147) | 45.8% (97/179) | 57.1% (186/326) |
| Seychelles | 50.0% (1/2) | 28.6% (5/7) | 33.3% (3/9) |
| Singapore | 55.6% (10/18) | 47.1% (9/17) | 51.4% (18/35) |
| Slovakia | 26.7% (24/90) | 69.7% (37/122) | 51.4% (109/212) |
| Slovenia | 52.0% (26/50) | 62.2% (17/45) | 56.8% (54/95) |
| Solomon Islands | 25.0% (1/4) | 66.7% (2/6) | 50.0% (5/10) |
| Somalia | 72.7% (8/11) | 38.5% (8/13) | 54.2% (13/24) |
| South Africa | 38.3% (23/60) | 42.9% (44/77) | 40.9% (56/137) |
| South Sudan | 72.7% (8/11) | 33.3% (8/12) | 52.2% (12/23) |
| Spain | 39.7% (278/700) | 75.1% (198/794) | 58.5% (874/1,494) |
| Sri Lanka | 53.5% (23/43) | 65.8% (13/38) | 59.3% (48/81) |
| Sudan | 61.0% (111/182) | 47.1% (128/242) | 53.1% (225/424) |
| Sweden | 47.7% (31/65) | 65.1% (29/83) | 57.4% (85/148) |
| Switzerland | 41.4% (65/157) | 63.5% (70/192) | 53.6% (187/349) |
| Syrian Arab Republic | 40.3% (56/139) | 57.5% (77/181) | 50.0% (160/320) |
| Taiwan, Province of China | 39.6% (44/111) | 66.4% (48/143) | 54.7% (139/254) |
| Tajikistan | 42.6% (26/61) | 61.7% (36/94) | 54.2% (84/155) |
| Tanzania, United Republic of | 47.5% (19/40) | 66.7% (17/51) | 58.2% (53/91) |
| Thailand | 55.4% (31/56) | 75.0% (14/56) | 65.2% (73/112) |
| Timor-Leste | 42.9% (3/7) | 60.0% (2/5) | 50.0% (6/12) |
| Togo | 38.9% (7/18) | 72.0% (7/25) | 58.1% (25/43) |
| Trinidad and Tobago | 61.5% (8/13) | 46.9% (17/32) | 51.1% (23/45) |
| Tunisia | 26.8% (22/82) | 67.5% (37/114) | 50.5% (99/196) |
| Türkiye | 67.5% (291/431) | 56.1% (250/569) | 61.0% (610/1,000) |
| Turkmenistan | 32.9% (79/240) | 63.8% (98/271) | 49.3% (252/511) |
| Uganda | 46.2% (24/52) | 52.7% (26/55) | 49.5% (53/107) |
| Ukraine | 45.5% (87/191) | 61.0% (89/228) | 53.9% (226/419) |
| United Arab Emirates | 22.2% (10/45) | 87.5% (7/56) | 58.4% (59/101) |
| United Kingdom | 32.2% (164/510) | 72.8% (174/640) | 54.8% (630/1,150) |
| United States | 47.7% (520/1,090) | 61.9% (498/1,308) | 55.5% (1,330/2,398) |
| United States Minor Outlying Islands | 100.0% (2/2) | 0.0% (2/2) | 50.0% (2/4) |
| Uruguay | 91.7% (11/12) | 14.3% (12/14) | 50.0% (13/26) |
| Uzbekistan | 57.6% (95/165) | 64.7% (71/201) | 61.5% (225/366) |
| Venezuela, Bolivarian Republic of | 50.6% (82/162) | 54.4% (93/204) | 52.7% (193/366) |
| Viet Nam | 37.0% (17/46) | 62.0% (19/50) | 50.0% (48/96) |
| Yemen | 59.1% (303/513) | 45.4% (401/735) | 51.0% (637/1,248) |
| Zambia | 63.6% (7/11) | 56.3% (7/16) | 59.3% (16/27) |
| Zimbabwe | 47.8% (11/23) | 52.9% (8/17) | 50.0% (20/40) |

**Table S4 - Proportions of Disease Groups Obtained from the Algorithm's Top-1 and Top-3 Predictions (by Country)**

|  |  | **Using Top-1 Diagnosis** |  |  |  |  | **Using Top-3 Diagnoses** |  |  |  |  |
| --- | --- | --- | --- | --- | --- | --- | --- | --- | --- | --- | --- |
| **Location** | **Total Queries** | **Benigns** | **Allergies** | **Infections** | **Malignancies** | **Estimated FP** | **Benigns** | **Allergies** | **Infections** | **Malignancies** | **Estimated FP** |
| Afghanistan | 995 | 178 (17.9%) | 195 (19.6%) | 268 (26.9%) | 16 (1.6%) | 8.2% | 465 (46.7%) | 492 (49.4%) | 510 (51.3%) | 36 (3.6%) | 7.2% |
| Åland Islands | 2 | 0 (0.0%) | 1 (50.0%) | 0 (0.0%) | 0 (0.0%) | - | 0 (0.0%) | 1 (50.0%) | 2 (100.0%) | 0 (0.0%) | - |
| Albania | 1,799 | 579 (32.2%) | 317 (17.6%) | 306 (17.0%) | 27 (1.5%) | 4.5% | 1,077 (59.9%) | 748 (41.6%) | 681 (37.9%) | 64 (3.6%) | 5.6% |
| Algeria | 7,611 | 1,659 (21.8%) | 1,473 (19.4%) | 1,655 (21.7%) | 66 (0.9%) | 3.8% | 3,852 (50.6%) | 3,555 (46.7%) | 3,547 (46.6%) | 237 (3.1%) | 5.8% |
| American Samoa | 4 | 1 (25.0%) | 2 (50.0%) | 1 (25.0%) | 0 (0.0%) | 0.0% | 2 (50.0%) | 4 (100.0%) | 2 (50.0%) | 0 (0.0%) | 0.0% |
| Andorra | 43 | 29 (67.4%) | 2 (4.7%) | 5 (11.6%) | 0 (0.0%) | 0.0% | 39 (90.7%) | 4 (9.3%) | 14 (32.6%) | 0 (0.0%) | 0.0% |
| Angola | 399 | 69 (17.3%) | 46 (11.5%) | 53 (13.3%) | 5 (1.3%) | 6.8% | 219 (54.9%) | 142 (35.6%) | 140 (35.1%) | 19 (4.8%) | 8.0% |
| Anguilla | 24 | 7 (29.2%) | 1 (4.2%) | 13 (54.2%) | 0 (0.0%) | 0.0% | 17 (70.8%) | 9 (37.5%) | 16 (66.7%) | 1 (4.2%) | 5.6% |
| Antigua and Barbuda | 64 | 12 (18.8%) | 15 (23.4%) | 6 (9.4%) | 0 (0.0%) | 0.0% | 30 (46.9%) | 34 (53.1%) | 16 (25.0%) | 2 (3.1%) | 6.3% |
| Argentina | 10,233 | 4,111 (40.2%) | 1,466 (14.3%) | 1,305 (12.8%) | 109 (1.1%) | 2.6% | 6,981 (68.2%) | 3,625 (35.4%) | 3,313 (32.4%) | 574 (5.6%) | 7.6% |
| Armenia | 581 | 308 (53.0%) | 61 (10.5%) | 67 (11.5%) | 1 (0.2%) | 0.3% | 418 (71.9%) | 185 (31.8%) | 153 (26.3%) | 18 (3.1%) | 4.1% |
| Aruba | 58 | 24 (41.4%) | 7 (12.1%) | 5 (8.6%) | 1 (1.7%) | 4.0% | 41 (70.7%) | 20 (34.5%) | 19 (32.8%) | 2 (3.4%) | 4.7% |
| Australia | 26,028 | 12,188 (46.8%) | 2,894 (11.1%) | 2,103 (8.1%) | 426 (1.6%) | 3.4% | 20,348 (78.2%) | 8,780 (33.7%) | 6,313 (24.3%) | 1,971 (7.6%) | 8.8% |
| Austria | 21,560 | 9,697 (45.0%) | 2,930 (13.6%) | 2,349 (10.9%) | 359 (1.7%) | 3.6% | 16,028 (74.3%) | 7,095 (32.9%) | 6,932 (32.2%) | 1,516 (7.0%) | 8.6% |
| Azerbaijan | 1,583 | 437 (27.6%) | 385 (24.3%) | 318 (20.1%) | 11 (0.7%) | 2.5% | 812 (51.3%) | 777 (49.1%) | 709 (44.8%) | 53 (3.3%) | 6.1% |
| Bahamas | 62 | 18 (29.0%) | 4 (6.5%) | 9 (14.5%) | 0 (0.0%) | 0.0% | 41 (66.1%) | 25 (40.3%) | 18 (29.0%) | 0 (0.0%) | 0.0% |
| Bahrain | 278 | 62 (22.3%) | 48 (17.3%) | 72 (25.9%) | 0 (0.0%) | 0.0% | 150 (54.0%) | 136 (48.9%) | 147 (52.9%) | 3 (1.1%) | 2.0% |
| Bangladesh | 5,514 | 1,192 (21.6%) | 1,033 (18.7%) | 1,061 (19.2%) | 42 (0.8%) | 3.4% | 2,778 (50.4%) | 2,476 (44.9%) | 2,382 (43.2%) | 166 (3.0%) | 5.6% |
| Barbados | 93 | 24 (25.8%) | 21 (22.6%) | 15 (16.1%) | 1 (1.1%) | 4.0% | 58 (62.4%) | 42 (45.2%) | 36 (38.7%) | 2 (2.2%) | 3.3% |
| Belarus | 12,240 | 5,802 (47.4%) | 1,834 (15.0%) | 1,356 (11.1%) | 114 (0.9%) | 1.9% | 8,614 (70.4%) | 4,193 (34.3%) | 3,681 (30.1%) | 547 (4.5%) | 6.0% |
| Belgium | 14,562 | 5,593 (38.4%) | 2,488 (17.1%) | 1,970 (13.5%) | 233 (1.6%) | 4.0% | 9,568 (65.7%) | 5,986 (41.1%) | 5,019 (34.5%) | 867 (6.0%) | 8.3% |
| Belize | 92 | 14 (15.2%) | 25 (27.2%) | 9 (9.8%) | 2 (2.2%) | 12.5% | 60 (65.2%) | 47 (51.1%) | 37 (40.2%) | 4 (4.3%) | 6.3% |
| Benin | 82 | 11 (13.4%) | 6 (7.3%) | 14 (17.1%) | 1 (1.2%) | 8.3% | 35 (42.7%) | 39 (47.6%) | 37 (45.1%) | 5 (6.1%) | 12.5% |
| Bermuda | 12 | 2 (16.7%) | 2 (16.7%) | 3 (25.0%) | 0 (0.0%) | 0.0% | 9 (75.0%) | 6 (50.0%) | 6 (50.0%) | 0 (0.0%) | 0.0% |
| Bhutan | 167 | 23 (13.8%) | 28 (16.8%) | 19 (11.4%) | 0 (0.0%) | 0.0% | 92 (55.1%) | 74 (44.3%) | 59 (35.3%) | 2 (1.2%) | 2.1% |
| Bolivia, Plurinational State of | 1,182 | 255 (21.6%) | 217 (18.4%) | 223 (18.9%) | 9 (0.8%) | 3.4% | 605 (51.2%) | 515 (43.6%) | 505 (42.7%) | 36 (3.0%) | 5.6% |
| Bosnia and Herzegovina | 5,660 | 2,772 (49.0%) | 850 (15.0%) | 630 (11.1%) | 108 (1.9%) | 3.8% | 4,079 (72.1%) | 1,796 (31.7%) | 1,715 (30.3%) | 440 (7.8%) | 9.7% |
| Botswana | 98 | 14 (14.3%) | 12 (12.2%) | 23 (23.5%) | 0 (0.0%) | 0.0% | 53 (54.1%) | 37 (37.8%) | 46 (46.9%) | 1 (1.0%) | 1.9% |
| Brazil | 54,974 | 18,305 (33.3%) | 8,063 (14.7%) | 7,642 (13.9%) | 841 (1.5%) | 4.4% | 35,640 (64.8%) | 20,560 (37.4%) | 19,429 (35.3%) | 3,333 (6.1%) | 8.6% |
| Brunei Darussalam | 42 | 10 (23.8%) | 5 (11.9%) | 10 (23.8%) | 0 (0.0%) | 0.0% | 21 (50.0%) | 13 (31.0%) | 19 (45.2%) | 2 (4.8%) | 8.7% |
| Bulgaria | 5,589 | 2,887 (51.7%) | 531 (9.5%) | 747 (13.4%) | 125 (2.2%) | 4.2% | 4,492 (80.4%) | 1,524 (27.3%) | 2,000 (35.8%) | 617 (11.0%) | 12.1% |
| Burkina Faso | 175 | 18 (10.3%) | 18 (10.3%) | 17 (9.7%) | 0 (0.0%) | 0.0% | 87 (49.7%) | 69 (39.4%) | 59 (33.7%) | 10 (5.7%) | 10.3% |
| Burundi | 28 | 0 (0.0%) | 3 (10.7%) | 0 (0.0%) | 0 (0.0%) | - | 15 (53.6%) | 14 (50.0%) | 7 (25.0%) | 0 (0.0%) | 0.0% |
| Cabo Verde | 71 | 19 (26.8%) | 21 (29.6%) | 2 (2.8%) | 0 (0.0%) | 0.0% | 40 (56.3%) | 42 (59.2%) | 13 (18.3%) | 0 (0.0%) | 0.0% |
| Cambodia | 371 | 110 (29.6%) | 60 (16.2%) | 73 (19.7%) | 6 (1.6%) | 5.2% | 217 (58.5%) | 157 (42.3%) | 152 (41.0%) | 18 (4.9%) | 7.7% |
| Cameroon | 636 | 123 (19.3%) | 59 (9.3%) | 106 (16.7%) | 3 (0.5%) | 2.4% | 355 (55.8%) | 250 (39.3%) | 254 (39.9%) | 13 (2.0%) | 3.5% |
| Canada | 39,719 | 17,391 (43.8%) | 5,903 (14.9%) | 4,501 (11.3%) | 1,277 (3.2%) | 6.8% | 28,334 (71.3%) | 13,815 (34.8%) | 12,329 (31.0%) | 4,653 (11.7%) | 14.1% |
| Cayman Islands | 15 | 5 (33.3%) | 1 (6.7%) | 1 (6.7%) | 0 (0.0%) | 0.0% | 12 (80.0%) | 4 (26.7%) | 4 (26.7%) | 0 (0.0%) | 0.0% |
| Central African Republic | 8 | 1 (12.5%) | 0 (0.0%) | 3 (37.5%) | 0 (0.0%) | 0.0% | 3 (37.5%) | 5 (62.5%) | 3 (37.5%) | 0 (0.0%) | 0.0% |
| Chad | 104 | 16 (15.4%) | 11 (10.6%) | 10 (9.6%) | 4 (3.8%) | 20.0% | 52 (50.0%) | 46 (44.2%) | 33 (31.7%) | 6 (5.8%) | 10.3% |
| Chile | 7,073 | 2,529 (35.8%) | 1,244 (17.6%) | 1,020 (14.4%) | 78 (1.1%) | 3.0% | 4,465 (63.1%) | 2,907 (41.1%) | 2,514 (35.5%) | 336 (4.8%) | 7.0% |
| China | 2,680 | 1,464 (54.6%) | 393 (14.7%) | 287 (10.7%) | 46 (1.7%) | 3.0% | 1,999 (74.6%) | 809 (30.2%) | 745 (27.8%) | 229 (8.5%) | 10.3% |
| Colombia | 10,791 | 2,731 (25.3%) | 2,311 (21.4%) | 2,128 (19.7%) | 163 (1.5%) | 5.6% | 5,644 (52.3%) | 5,043 (46.7%) | 4,652 (43.1%) | 504 (4.7%) | 8.2% |
| Comoros | 15 | 1 (6.7%) | 10 (66.7%) | 0 (0.0%) | 0 (0.0%) | 0.0% | 2 (13.3%) | 15 (100.0%) | 4 (26.7%) | 0 (0.0%) | 0.0% |
| Congo | 121 | 9 (7.4%) | 15 (12.4%) | 20 (16.5%) | 1 (0.8%) | 10.0% | 60 (49.6%) | 57 (47.1%) | 44 (36.4%) | 1 (0.8%) | 1.6% |
| Congo, The Democratic Republic of the | 433 | 74 (17.1%) | 48 (11.1%) | 46 (10.6%) | 3 (0.7%) | 3.9% | 250 (57.7%) | 147 (33.9%) | 153 (35.3%) | 20 (4.6%) | 7.4% |
| Cook Islands | 5 | 0 (0.0%) | 1 (20.0%) | 0 (0.0%) | 0 (0.0%) | - | 5 (100.0%) | 2 (40.0%) | 0 (0.0%) | 0 (0.0%) | 0.0% |
| Costa Rica | 1,433 | 405 (28.3%) | 244 (17.0%) | 201 (14.0%) | 4 (0.3%) | 1.0% | 828 (57.8%) | 623 (43.5%) | 534 (37.3%) | 35 (2.4%) | 4.1% |
| Côte d'Ivoire | 1,115 | 208 (18.7%) | 136 (12.2%) | 213 (19.1%) | 3 (0.3%) | 1.4% | 569 (51.0%) | 437 (39.2%) | 462 (41.4%) | 40 (3.6%) | 6.6% |
| Croatia | 17,600 | 11,069 (62.9%) | 1,367 (7.8%) | 1,543 (8.8%) | 343 (1.9%) | 3.0% | 14,468 (82.2%) | 3,533 (20.1%) | 4,273 (24.3%) | 1,483 (8.4%) | 9.3% |
| Cuba | 1,770 | 440 (24.9%) | 302 (17.1%) | 310 (17.5%) | 21 (1.2%) | 4.6% | 979 (55.3%) | 800 (45.2%) | 770 (43.5%) | 64 (3.6%) | 6.1% |
| Curaçao | 75 | 36 (48.0%) | 4 (5.3%) | 9 (12.0%) | 3 (4.0%) | 7.7% | 59 (78.7%) | 24 (32.0%) | 22 (29.3%) | 5 (6.7%) | 7.8% |
| Cyprus | 1,795 | 750 (41.8%) | 226 (12.6%) | 281 (15.7%) | 24 (1.3%) | 3.1% | 1,217 (67.8%) | 615 (34.3%) | 657 (36.6%) | 89 (5.0%) | 6.8% |
| Czechia | 17,915 | 9,582 (53.5%) | 2,068 (11.5%) | 1,663 (9.3%) | 211 (1.2%) | 2.2% | 13,460 (75.1%) | 5,072 (28.3%) | 4,768 (26.6%) | 1,121 (6.3%) | 7.7% |
| Denmark | 7,944 | 3,876 (48.8%) | 912 (11.5%) | 874 (11.0%) | 159 (2.0%) | 3.9% | 6,129 (77.2%) | 2,306 (29.0%) | 2,381 (30.0%) | 610 (7.7%) | 9.1% |
| Djibouti | 88 | 10 (11.4%) | 8 (9.1%) | 6 (6.8%) | 0 (0.0%) | 0.0% | 37 (42.0%) | 36 (40.9%) | 28 (31.8%) | 1 (1.1%) | 2.6% |
| Dominica | 11 | 4 (36.4%) | 1 (9.1%) | 3 (27.3%) | 2 (18.2%) | 33.3% | 6 (54.5%) | 5 (45.5%) | 6 (54.5%) | 2 (18.2%) | 25.0% |
| Dominican Republic | 1,156 | 302 (26.1%) | 115 (9.9%) | 163 (14.1%) | 9 (0.8%) | 2.9% | 807 (69.8%) | 323 (27.9%) | 489 (42.3%) | 40 (3.5%) | 4.7% |
| Ecuador | 2,229 | 574 (25.8%) | 378 (17.0%) | 362 (16.2%) | 16 (0.7%) | 2.7% | 1,270 (57.0%) | 930 (41.7%) | 834 (37.4%) | 82 (3.7%) | 6.1% |
| Egypt | 7,882 | 1,965 (24.9%) | 1,434 (18.2%) | 1,639 (20.8%) | 48 (0.6%) | 2.4% | 4,252 (53.9%) | 3,455 (43.8%) | 3,541 (44.9%) | 233 (3.0%) | 5.2% |
| El Salvador | 1,071 | 231 (21.6%) | 224 (20.9%) | 258 (24.1%) | 15 (1.4%) | 6.1% | 548 (51.2%) | 514 (48.0%) | 494 (46.1%) | 53 (4.9%) | 8.8% |
| Equatorial Guinea | 24 | 2 (8.3%) | 8 (33.3%) | 3 (12.5%) | 0 (0.0%) | 0.0% | 10 (41.7%) | 18 (75.0%) | 6 (25.0%) | 1 (4.2%) | 9.1% |
| Eritrea | 2 | 0 (0.0%) | 1 (50.0%) | 1 (50.0%) | 0 (0.0%) | - | 1 (50.0%) | 1 (50.0%) | 1 (50.0%) | 0 (0.0%) | 0.0% |
| Estonia | 996 | 510 (51.2%) | 136 (13.7%) | 121 (12.1%) | 10 (1.0%) | 1.9% | 717 (72.0%) | 326 (32.7%) | 300 (30.1%) | 35 (3.5%) | 4.7% |
| Eswatini | 26 | 2 (7.7%) | 5 (19.2%) | 7 (26.9%) | 0 (0.0%) | 0.0% | 11 (42.3%) | 12 (46.2%) | 10 (38.5%) | 1 (3.8%) | 8.3% |
| Ethiopia | 1,550 | 298 (19.2%) | 207 (13.4%) | 256 (16.5%) | 22 (1.4%) | 6.9% | 838 (54.1%) | 581 (37.5%) | 596 (38.5%) | 62 (4.0%) | 6.9% |
| Falkland Islands (Malvinas) | 4 | 4 (100.0%) | 0 (0.0%) | 0 (0.0%) | 0 (0.0%) | 0.0% | 4 (100.0%) | 0 (0.0%) | 1 (25.0%) | 0 (0.0%) | 0.0% |
| Faroe Islands | 26 | 16 (61.5%) | 2 (7.7%) | 0 (0.0%) | 1 (3.8%) | 5.9% | 20 (76.9%) | 12 (46.2%) | 1 (3.8%) | 1 (3.8%) | 4.8% |
| Fiji | 245 | 68 (27.8%) | 56 (22.9%) | 38 (15.5%) | 2 (0.8%) | 2.9% | 148 (60.4%) | 115 (46.9%) | 103 (42.0%) | 14 (5.7%) | 8.6% |
| Finland | 2,795 | 1,394 (49.9%) | 312 (11.2%) | 319 (11.4%) | 36 (1.3%) | 2.5% | 2,097 (75.0%) | 798 (28.6%) | 927 (33.2%) | 174 (6.2%) | 7.7% |
| France | 68,408 | 30,430 (44.5%) | 10,399 (15.2%) | 8,538 (12.5%) | 1,393 (2.0%) | 4.4% | 48,190 (70.4%) | 24,231 (35.4%) | 22,646 (33.1%) | 5,533 (8.1%) | 10.3% |
| French Guiana | 310 | 250 (80.6%) | 12 (3.9%) | 10 (3.2%) | 4 (1.3%) | 1.6% | 284 (91.6%) | 22 (7.1%) | 42 (13.5%) | 25 (8.1%) | 8.1% |
| French Polynesia | 111 | 30 (27.0%) | 29 (26.1%) | 11 (9.9%) | 1 (0.9%) | 3.2% | 61 (55.0%) | 59 (53.2%) | 29 (26.1%) | 7 (6.3%) | 10.3% |
| Gabon | 94 | 10 (10.6%) | 15 (16.0%) | 14 (14.9%) | 1 (1.1%) | 9.1% | 49 (52.1%) | 34 (36.2%) | 32 (34.0%) | 1 (1.1%) | 2.0% |
| Gambia | 71 | 17 (23.9%) | 9 (12.7%) | 7 (9.9%) | 0 (0.0%) | 0.0% | 45 (63.4%) | 24 (33.8%) | 12 (16.9%) | 0 (0.0%) | 0.0% |
| Georgia | 679 | 271 (39.9%) | 81 (11.9%) | 139 (20.5%) | 12 (1.8%) | 4.2% | 448 (66.0%) | 228 (33.6%) | 278 (40.9%) | 31 (4.6%) | 6.5% |
| Germany | 100,285 | 51,000 (50.9%) | 12,027 (12.0%) | 10,413 (10.4%) | 1,935 (1.9%) | 3.7% | 74,353 (74.1%) | 29,473 (29.4%) | 29,825 (29.7%) | 7,686 (7.7%) | 9.4% |
| Ghana | 946 | 182 (19.2%) | 91 (9.6%) | 137 (14.5%) | 9 (1.0%) | 4.7% | 556 (58.8%) | 342 (36.2%) | 347 (36.7%) | 32 (3.4%) | 5.4% |
| Gibraltar | 24 | 15 (62.5%) | 3 (12.5%) | 2 (8.3%) | 0 (0.0%) | 0.0% | 18 (75.0%) | 7 (29.2%) | 8 (33.3%) | 2 (8.3%) | 10.0% |
| Greece | 9,949 | 4,504 (45.3%) | 1,389 (14.0%) | 1,166 (11.7%) | 59 (0.6%) | 1.3% | 7,057 (70.9%) | 3,304 (33.2%) | 3,236 (32.5%) | 432 (4.3%) | 5.8% |
| Greenland | 12 | 2 (16.7%) | 1 (8.3%) | 2 (16.7%) | 0 (0.0%) | 0.0% | 6 (50.0%) | 4 (33.3%) | 3 (25.0%) | 0 (0.0%) | 0.0% |
| Grenada | 9 | 5 (55.6%) | 1 (11.1%) | 0 (0.0%) | 0 (0.0%) | 0.0% | 6 (66.7%) | 3 (33.3%) | 4 (44.4%) | 0 (0.0%) | 0.0% |
| Guadeloupe | 437 | 242 (55.4%) | 40 (9.2%) | 36 (8.2%) | 6 (1.4%) | 2.4% | 328 (75.1%) | 105 (24.0%) | 87 (19.9%) | 34 (7.8%) | 9.4% |
| Guam | 100 | 34 (34.0%) | 9 (9.0%) | 6 (6.0%) | 0 (0.0%) | 0.0% | 66 (66.0%) | 31 (31.0%) | 28 (28.0%) | 1 (1.0%) | 1.5% |
| Guatemala | 1,430 | 269 (18.8%) | 305 (21.3%) | 277 (19.4%) | 10 (0.7%) | 3.6% | 749 (52.4%) | 673 (47.1%) | 640 (44.8%) | 42 (2.9%) | 5.3% |
| Guernsey | 1 | 0 (0.0%) | 0 (0.0%) | 1 (100.0%) | 0 (0.0%) | - | 1 (100.0%) | 0 (0.0%) | 1 (100.0%) | 0 (0.0%) | 0.0% |
| Guinea | 128 | 28 (21.9%) | 7 (5.5%) | 13 (10.2%) | 0 (0.0%) | 0.0% | 81 (63.3%) | 44 (34.4%) | 52 (40.6%) | 4 (3.1%) | 4.7% |
| Guinea-Bissau | 6 | 0 (0.0%) | 1 (16.7%) | 0 (0.0%) | 0 (0.0%) | - | 4 (66.7%) | 2 (33.3%) | 2 (33.3%) | 0 (0.0%) | 0.0% |
| Guyana | 71 | 10 (14.1%) | 7 (9.9%) | 13 (18.3%) | 0 (0.0%) | 0.0% | 38 (53.5%) | 19 (26.8%) | 30 (42.3%) | 2 (2.8%) | 5.0% |
| Haiti | 78 | 17 (21.8%) | 10 (12.8%) | 6 (7.7%) | 0 (0.0%) | 0.0% | 47 (60.3%) | 28 (35.9%) | 22 (28.2%) | 3 (3.8%) | 6.0% |
| Honduras | 846 | 193 (22.8%) | 161 (19.0%) | 150 (17.7%) | 8 (0.9%) | 4.0% | 461 (54.5%) | 378 (44.7%) | 350 (41.4%) | 24 (2.8%) | 4.9% |
| Hong Kong | 1,505 | 732 (48.6%) | 215 (14.3%) | 133 (8.8%) | 17 (1.1%) | 2.3% | 1,055 (70.1%) | 512 (34.0%) | 461 (30.6%) | 91 (6.0%) | 7.9% |
| Hungary | 47,937 | 21,224 (44.3%) | 7,418 (15.5%) | 6,168 (12.9%) | 759 (1.6%) | 3.5% | 32,808 (68.4%) | 16,713 (34.9%) | 16,358 (34.1%) | 3,344 (7.0%) | 9.2% |
| Iceland | 271 | 124 (45.8%) | 47 (17.3%) | 29 (10.7%) | 9 (3.3%) | 6.8% | 199 (73.4%) | 110 (40.6%) | 88 (32.5%) | 25 (9.2%) | 11.2% |
| India | 18,126 | 4,957 (27.3%) | 3,073 (17.0%) | 2,944 (16.2%) | 263 (1.5%) | 5.0% | 10,247 (56.5%) | 7,526 (41.5%) | 6,544 (36.1%) | 915 (5.0%) | 8.2% |
| Indonesia | 6,554 | 1,471 (22.4%) | 1,248 (19.0%) | 1,196 (18.2%) | 57 (0.9%) | 3.7% | 3,382 (51.6%) | 3,022 (46.1%) | 2,708 (41.3%) | 225 (3.4%) | 6.2% |
| Iran, Islamic Republic of | 3,374 | 883 (26.2%) | 566 (16.8%) | 676 (20.0%) | 32 (0.9%) | 3.5% | 1,877 (55.6%) | 1,351 (40.0%) | 1,524 (45.2%) | 107 (3.2%) | 5.4% |
| Iraq | 4,433 | 859 (19.4%) | 781 (17.6%) | 951 (21.5%) | 29 (0.7%) | 3.3% | 2,321 (52.4%) | 1,950 (44.0%) | 2,044 (46.1%) | 106 (2.4%) | 4.4% |
| Ireland | 9,914 | 3,673 (37.0%) | 1,213 (12.2%) | 986 (9.9%) | 106 (1.1%) | 2.8% | 7,108 (71.7%) | 3,666 (37.0%) | 3,064 (30.9%) | 614 (6.2%) | 8.0% |
| Isle of Man | 6 | 4 (66.7%) | 1 (16.7%) | 1 (16.7%) | 0 (0.0%) | 0.0% | 5 (83.3%) | 2 (33.3%) | 2 (33.3%) | 0 (0.0%) | 0.0% |
| Israel | 6,709 | 2,211 (33.0%) | 874 (13.0%) | 978 (14.6%) | 253 (3.8%) | 10.3% | 4,612 (68.7%) | 2,402 (35.8%) | 2,376 (35.4%) | 663 (9.9%) | 12.6% |
| Italy | 151,574 | 79,350 (52.4%) | 16,232 (10.7%) | 15,258 (10.1%) | 3,550 (2.3%) | 4.3% | 117,146 (77.3%) | 41,482 (27.4%) | 42,790 (28.2%) | 13,738 (9.1%) | 10.5% |
| Jamaica | 488 | 83 (17.0%) | 55 (11.3%) | 63 (12.9%) | 7 (1.4%) | 7.8% | 281 (57.6%) | 185 (37.9%) | 157 (32.2%) | 21 (4.3%) | 7.0% |
| Japan | 14,951 | 4,769 (31.9%) | 3,884 (26.0%) | 2,299 (15.4%) | 182 (1.2%) | 3.7% | 8,379 (56.0%) | 7,324 (49.0%) | 5,775 (38.6%) | 718 (4.8%) | 7.9% |
| Jersey | 3 | 2 (66.7%) | 0 (0.0%) | 0 (0.0%) | 0 (0.0%) | 0.0% | 3 (100.0%) | 0 (0.0%) | 1 (33.3%) | 1 (33.3%) | 25.0% |
| Jordan | 2,132 | 476 (22.3%) | 385 (18.1%) | 372 (17.4%) | 13 (0.6%) | 2.7% | 1,130 (53.0%) | 972 (45.6%) | 915 (42.9%) | 52 (2.4%) | 4.4% |
| Kazakhstan | 6,105 | 2,820 (46.2%) | 1,013 (16.6%) | 628 (10.3%) | 56 (0.9%) | 1.9% | 4,246 (69.5%) | 2,175 (35.6%) | 1,796 (29.4%) | 334 (5.5%) | 7.3% |
| Kenya | 2,191 | 369 (16.8%) | 245 (11.2%) | 297 (13.6%) | 17 (0.8%) | 4.4% | 1,216 (55.5%) | 808 (36.9%) | 797 (36.4%) | 75 (3.4%) | 5.8% |
| Kiribati | 4 | 0 (0.0%) | 2 (50.0%) | 0 (0.0%) | 0 (0.0%) | - | 2 (50.0%) | 3 (75.0%) | 0 (0.0%) | 0 (0.0%) | 0.0% |
| Korea, Democratic People's Republic of | 1 | 0 (0.0%) | 1 (100.0%) | 0 (0.0%) | 0 (0.0%) | - | 0 (0.0%) | 1 (100.0%) | 1 (100.0%) | 0 (0.0%) | - |
| Korea, Republic of | 382,602 | 249,395 (65.2%) | 27,069 (7.1%) | 27,570 (7.2%) | 7,957 (2.1%) | 3.1% | 327,939 (85.7%) | 68,845 (18.0%) | 87,568 (22.9%) | 44,524 (11.6%) | 12.0% |
| Kosovo | 22 | 16 (72.7%) | 2 (9.1%) | 1 (4.5%) | 0 (0.0%) | 0.0% | 20 (90.9%) | 4 (18.2%) | 2 (9.1%) | 0 (0.0%) | 0.0% |
| Kuwait | 363 | 84 (23.1%) | 64 (17.6%) | 37 (10.2%) | 1 (0.3%) | 1.2% | 197 (54.3%) | 157 (43.3%) | 121 (33.3%) | 10 (2.8%) | 4.8% |
| Kyrgyzstan | 955 | 292 (30.6%) | 196 (20.5%) | 173 (18.1%) | 7 (0.7%) | 2.3% | 538 (56.3%) | 425 (44.5%) | 383 (40.1%) | 47 (4.9%) | 8.0% |
| Lao People's Democratic Republic | 90 | 14 (15.6%) | 14 (15.6%) | 21 (23.3%) | 0 (0.0%) | 0.0% | 38 (42.2%) | 52 (57.8%) | 38 (42.2%) | 0 (0.0%) | 0.0% |
| Latvia | 2,058 | 985 (47.9%) | 318 (15.5%) | 199 (9.7%) | 24 (1.2%) | 2.4% | 1,499 (72.8%) | 710 (34.5%) | 621 (30.2%) | 148 (7.2%) | 9.0% |
| Lebanon | 1,478 | 370 (25.0%) | 251 (17.0%) | 339 (22.9%) | 6 (0.4%) | 1.6% | 800 (54.1%) | 614 (41.5%) | 722 (48.8%) | 28 (1.9%) | 3.4% |
| Lesotho | 12 | 2 (16.7%) | 2 (16.7%) | 1 (8.3%) | 0 (0.0%) | 0.0% | 7 (58.3%) | 4 (33.3%) | 3 (25.0%) | 0 (0.0%) | 0.0% |
| Liberia | 35 | 9 (25.7%) | 2 (5.7%) | 7 (20.0%) | 0 (0.0%) | 0.0% | 20 (57.1%) | 18 (51.4%) | 14 (40.0%) | 0 (0.0%) | 0.0% |
| Libya | 2,196 | 459 (20.9%) | 435 (19.8%) | 486 (22.1%) | 33 (1.5%) | 6.7% | 1,051 (47.9%) | 1,030 (46.9%) | 1,013 (46.1%) | 91 (4.1%) | 8.0% |
| Liechtenstein | 55 | 14 (25.5%) | 20 (36.4%) | 19 (34.5%) | 0 (0.0%) | 0.0% | 29 (52.7%) | 29 (52.7%) | 34 (61.8%) | 5 (9.1%) | 14.7% |
| Lithuania | 3,006 | 1,668 (55.5%) | 317 (10.5%) | 264 (8.8%) | 35 (1.2%) | 2.1% | 2,389 (79.5%) | 777 (25.8%) | 795 (26.4%) | 157 (5.2%) | 6.2% |
| Luxembourg | 688 | 353 (51.3%) | 76 (11.0%) | 63 (9.2%) | 7 (1.0%) | 1.9% | 556 (80.8%) | 169 (24.6%) | 208 (30.2%) | 34 (4.9%) | 5.8% |
| Macao | 40 | 8 (20.0%) | 8 (20.0%) | 6 (15.0%) | 0 (0.0%) | 0.0% | 16 (40.0%) | 20 (50.0%) | 15 (37.5%) | 2 (5.0%) | 11.1% |
| Madagascar | 174 | 34 (19.5%) | 19 (10.9%) | 18 (10.3%) | 0 (0.0%) | 0.0% | 106 (60.9%) | 61 (35.1%) | 52 (29.9%) | 5 (2.9%) | 4.5% |
| Malawi | 174 | 23 (13.2%) | 10 (5.7%) | 16 (9.2%) | 2 (1.1%) | 8.0% | 112 (64.4%) | 74 (42.5%) | 60 (34.5%) | 9 (5.2%) | 7.4% |
| Malaysia | 2,733 | 688 (25.2%) | 492 (18.0%) | 602 (22.0%) | 20 (0.7%) | 2.8% | 1,487 (54.4%) | 1,189 (43.5%) | 1,237 (45.3%) | 99 (3.6%) | 6.2% |
| Maldives | 96 | 36 (37.5%) | 13 (13.5%) | 9 (9.4%) | 1 (1.0%) | 2.7% | 65 (67.7%) | 35 (36.5%) | 26 (27.1%) | 3 (3.1%) | 4.4% |
| Mali | 176 | 14 (8.0%) | 12 (6.8%) | 17 (9.7%) | 0 (0.0%) | 0.0% | 82 (46.6%) | 71 (40.3%) | 62 (35.2%) | 5 (2.8%) | 5.7% |
| Malta | 690 | 350 (50.7%) | 99 (14.3%) | 97 (14.1%) | 7 (1.0%) | 2.0% | 497 (72.0%) | 220 (31.9%) | 219 (31.7%) | 35 (5.1%) | 6.6% |
| Marshall Islands | 2 | 1 (50.0%) | 0 (0.0%) | 1 (50.0%) | 1 (50.0%) | 50.0% | 2 (100.0%) | 0 (0.0%) | 2 (100.0%) | 1 (50.0%) | 33.3% |
| Martinique | 27 | 7 (25.9%) | 2 (7.4%) | 3 (11.1%) | 0 (0.0%) | 0.0% | 21 (77.8%) | 9 (33.3%) | 8 (29.6%) | 0 (0.0%) | 0.0% |
| Mauritania | 146 | 43 (29.5%) | 13 (8.9%) | 22 (15.1%) | 0 (0.0%) | 0.0% | 98 (67.1%) | 55 (37.7%) | 62 (42.5%) | 1 (0.7%) | 1.0% |
| Mauritius | 437 | 99 (22.7%) | 56 (12.8%) | 60 (13.7%) | 3 (0.7%) | 2.9% | 249 (57.0%) | 188 (43.0%) | 153 (35.0%) | 8 (1.8%) | 3.1% |
| Mayotte | 14 | 8 (57.1%) | 2 (14.3%) | 0 (0.0%) | 0 (0.0%) | 0.0% | 9 (64.3%) | 7 (50.0%) | 3 (21.4%) | 1 (7.1%) | 10.0% |
| Mexico | 11,429 | 3,125 (27.3%) | 1,952 (17.1%) | 1,903 (16.7%) | 114 (1.0%) | 3.5% | 6,747 (59.0%) | 4,739 (41.5%) | 4,528 (39.6%) | 443 (3.9%) | 6.2% |
| Micronesia, Federated States of | 1 | 1 (100.0%) | 0 (0.0%) | 0 (0.0%) | 0 (0.0%) | 0.0% | 1 (100.0%) | 1 (100.0%) | 0 (0.0%) | 0 (0.0%) | 0.0% |
| Moldova, Republic of | 1,741 | 646 (37.1%) | 345 (19.8%) | 222 (12.8%) | 10 (0.6%) | 1.5% | 1,124 (64.6%) | 747 (42.9%) | 565 (32.5%) | 65 (3.7%) | 5.5% |
| Monaco | 168 | 86 (51.2%) | 22 (13.1%) | 15 (8.9%) | 1 (0.6%) | 1.1% | 124 (73.8%) | 48 (28.6%) | 52 (31.0%) | 5 (3.0%) | 3.9% |
| Mongolia | 225 | 33 (14.7%) | 36 (16.0%) | 67 (29.8%) | 0 (0.0%) | 0.0% | 109 (48.4%) | 88 (39.1%) | 134 (59.6%) | 3 (1.3%) | 2.7% |
| Montenegro | 938 | 386 (41.2%) | 145 (15.5%) | 147 (15.7%) | 7 (0.7%) | 1.8% | 591 (63.0%) | 359 (38.3%) | 377 (40.2%) | 65 (6.9%) | 9.9% |
| Montserrat | 1 | 1 (100.0%) | 0 (0.0%) | 1 (100.0%) | 0 (0.0%) | 0.0% | 1 (100.0%) | 0 (0.0%) | 1 (100.0%) | 0 (0.0%) | 0.0% |
| Morocco | 6,602 | 1,572 (23.8%) | 989 (15.0%) | 1,180 (17.9%) | 84 (1.3%) | 5.1% | 3,516 (53.3%) | 2,665 (40.4%) | 2,766 (41.9%) | 325 (4.9%) | 8.5% |
| Mozambique | 555 | 116 (20.9%) | 77 (13.9%) | 90 (16.2%) | 1 (0.2%) | 0.9% | 311 (56.0%) | 223 (40.2%) | 214 (38.6%) | 13 (2.3%) | 4.0% |
| Myanmar | 72 | 17 (23.6%) | 10 (13.9%) | 14 (19.4%) | 0 (0.0%) | 0.0% | 38 (52.8%) | 34 (47.2%) | 31 (43.1%) | 0 (0.0%) | 0.0% |
| Nepal | 1,173 | 236 (20.1%) | 177 (15.1%) | 200 (17.1%) | 3 (0.3%) | 1.3% | 604 (51.5%) | 468 (39.9%) | 497 (42.4%) | 16 (1.4%) | 2.6% |
| Netherlands | 16,454 | 7,242 (44.0%) | 2,320 (14.1%) | 1,916 (11.6%) | 328 (2.0%) | 4.3% | 12,010 (73.0%) | 5,688 (34.6%) | 5,298 (32.2%) | 1,244 (7.6%) | 9.4% |
| New Caledonia | 102 | 31 (30.4%) | 25 (24.5%) | 12 (11.8%) | 4 (3.9%) | 11.4% | 61 (59.8%) | 48 (47.1%) | 40 (39.2%) | 9 (8.8%) | 12.9% |
| New Zealand | 5,031 | 2,537 (50.4%) | 555 (11.0%) | 514 (10.2%) | 111 (2.2%) | 4.2% | 3,853 (76.6%) | 1,456 (28.9%) | 1,453 (28.9%) | 422 (8.4%) | 9.9% |
| Nicaragua | 724 | 157 (21.7%) | 98 (13.5%) | 114 (15.7%) | 2 (0.3%) | 1.3% | 426 (58.8%) | 279 (38.5%) | 281 (38.8%) | 14 (1.9%) | 3.2% |
| Niger | 64 | 17 (26.6%) | 9 (14.1%) | 6 (9.4%) | 0 (0.0%) | 0.0% | 33 (51.6%) | 27 (42.2%) | 24 (37.5%) | 0 (0.0%) | 0.0% |
| Nigeria | 1,491 | 215 (14.4%) | 137 (9.2%) | 163 (10.9%) | 5 (0.3%) | 2.3% | 804 (53.9%) | 555 (37.2%) | 467 (31.3%) | 57 (3.8%) | 6.6% |
| North Macedonia | 2,259 | 955 (42.3%) | 371 (16.4%) | 318 (14.1%) | 27 (1.2%) | 2.7% | 1,539 (68.1%) | 794 (35.1%) | 796 (35.2%) | 136 (6.0%) | 8.1% |
| Northern Mariana Islands | 1 | 0 (0.0%) | 0 (0.0%) | 0 (0.0%) | 0 (0.0%) | - | 0 (0.0%) | 1 (100.0%) | 1 (100.0%) | 0 (0.0%) | - |
| Norway | 1,335 | 676 (50.6%) | 147 (11.0%) | 110 (8.2%) | 14 (1.0%) | 2.0% | 1,038 (77.8%) | 393 (29.4%) | 382 (28.6%) | 67 (5.0%) | 6.1% |
| Oman | 285 | 72 (25.3%) | 30 (10.5%) | 53 (18.6%) | 1 (0.4%) | 1.4% | 163 (57.2%) | 116 (40.7%) | 110 (38.6%) | 8 (2.8%) | 4.7% |
| Pakistan | 7,220 | 1,441 (20.0%) | 1,332 (18.4%) | 1,652 (22.9%) | 53 (0.7%) | 3.5% | 3,662 (50.7%) | 3,235 (44.8%) | 3,414 (47.3%) | 192 (2.7%) | 5.0% |
| Palau | 7 | 0 (0.0%) | 2 (28.6%) | 3 (42.9%) | 0 (0.0%) | - | 1 (14.3%) | 6 (85.7%) | 4 (57.1%) | 0 (0.0%) | 0.0% |
| Palestine, State of | 37 | 12 (32.4%) | 9 (24.3%) | 8 (21.6%) | 1 (2.7%) | 7.7% | 20 (54.1%) | 16 (43.2%) | 21 (56.8%) | 1 (2.7%) | 4.8% |
| Panama | 1,354 | 236 (17.4%) | 197 (14.5%) | 236 (17.4%) | 5 (0.4%) | 2.1% | 701 (51.8%) | 566 (41.8%) | 522 (38.6%) | 21 (1.6%) | 2.9% |
| Papua New Guinea | 216 | 31 (14.4%) | 26 (12.0%) | 33 (15.3%) | 3 (1.4%) | 8.8% | 136 (63.0%) | 76 (35.2%) | 83 (38.4%) | 17 (7.9%) | 11.1% |
| Paraguay | 961 | 230 (23.9%) | 179 (18.6%) | 169 (17.6%) | 7 (0.7%) | 3.0% | 520 (54.1%) | 440 (45.8%) | 407 (42.4%) | 34 (3.5%) | 6.1% |
| Peru | 3,283 | 788 (24.0%) | 518 (15.8%) | 560 (17.1%) | 27 (0.8%) | 3.3% | 1,852 (56.4%) | 1,327 (40.4%) | 1,290 (39.3%) | 97 (3.0%) | 5.0% |
| Philippines | 9,344 | 2,077 (22.2%) | 2,029 (21.7%) | 1,715 (18.4%) | 66 (0.7%) | 3.1% | 4,726 (50.6%) | 4,607 (49.3%) | 3,826 (40.9%) | 258 (2.8%) | 5.2% |
| Pitcairn | 2 | 0 (0.0%) | 0 (0.0%) | 0 (0.0%) | 0 (0.0%) | - | 0 (0.0%) | 1 (50.0%) | 1 (50.0%) | 0 (0.0%) | - |
| Poland | 31,847 | 16,108 (50.6%) | 4,019 (12.6%) | 3,719 (11.7%) | 470 (1.5%) | 2.8% | 23,537 (73.9%) | 9,614 (30.2%) | 9,721 (30.5%) | 2,423 (7.6%) | 9.3% |
| Portugal | 11,593 | 5,601 (48.3%) | 1,391 (12.0%) | 1,480 (12.8%) | 163 (1.4%) | 2.8% | 8,519 (73.5%) | 3,449 (29.8%) | 3,909 (33.7%) | 798 (6.9%) | 8.6% |
| Puerto Rico | 1,115 | 378 (33.9%) | 168 (15.1%) | 140 (12.6%) | 14 (1.3%) | 3.6% | 749 (67.2%) | 428 (38.4%) | 389 (34.9%) | 63 (5.7%) | 7.8% |
| Qatar | 529 | 119 (22.5%) | 85 (16.1%) | 93 (17.6%) | 7 (1.3%) | 5.6% | 295 (55.8%) | 221 (41.8%) | 214 (40.5%) | 17 (3.2%) | 5.4% |
| Réunion | 335 | 132 (39.4%) | 45 (13.4%) | 39 (11.6%) | 1 (0.3%) | 0.8% | 219 (65.4%) | 125 (37.3%) | 105 (31.3%) | 14 (4.2%) | 6.0% |
| Romania | 19,074 | 8,054 (42.2%) | 2,895 (15.2%) | 2,642 (13.9%) | 299 (1.6%) | 3.6% | 12,614 (66.1%) | 6,920 (36.3%) | 6,551 (34.3%) | 1,164 (6.1%) | 8.4% |
| Russian Federation | 64,997 | 17,925 (27.6%) | 13,723 (21.1%) | 10,653 (16.4%) | 619 (1.0%) | 3.3% | 36,585 (56.3%) | 31,816 (48.9%) | 26,173 (40.3%) | 2,162 (3.3%) | 5.6% |
| Rwanda | 100 | 15 (15.0%) | 8 (8.0%) | 15 (15.0%) | 0 (0.0%) | 0.0% | 58 (58.0%) | 34 (34.0%) | 36 (36.0%) | 10 (10.0%) | 14.7% |
| Saint Barthélemy | 3 | 2 (66.7%) | 1 (33.3%) | 0 (0.0%) | 0 (0.0%) | 0.0% | 3 (100.0%) | 1 (33.3%) | 1 (33.3%) | 0 (0.0%) | 0.0% |
| Saint Kitts and Nevis | 15 | 3 (20.0%) | 2 (13.3%) | 2 (13.3%) | 0 (0.0%) | 0.0% | 9 (60.0%) | 7 (46.7%) | 5 (33.3%) | 0 (0.0%) | 0.0% |
| Saint Lucia | 65 | 20 (30.8%) | 8 (12.3%) | 14 (21.5%) | 0 (0.0%) | 0.0% | 48 (73.8%) | 25 (38.5%) | 37 (56.9%) | 1 (1.5%) | 2.0% |
| Saint Pierre and Miquelon | 3 | 0 (0.0%) | 1 (33.3%) | 2 (66.7%) | 0 (0.0%) | - | 1 (33.3%) | 2 (66.7%) | 3 (100.0%) | 0 (0.0%) | 0.0% |
| Saint Vincent and the Grenadines | 9 | 1 (11.1%) | 0 (0.0%) | 3 (33.3%) | 0 (0.0%) | 0.0% | 6 (66.7%) | 4 (44.4%) | 3 (33.3%) | 0 (0.0%) | 0.0% |
| Samoa | 4 | 1 (25.0%) | 3 (75.0%) | 0 (0.0%) | 0 (0.0%) | 0.0% | 1 (25.0%) | 3 (75.0%) | 2 (50.0%) | 0 (0.0%) | 0.0% |
| San Marino | 6 | 4 (66.7%) | 0 (0.0%) | 2 (33.3%) | 0 (0.0%) | 0.0% | 5 (83.3%) | 1 (16.7%) | 4 (66.7%) | 0 (0.0%) | 0.0% |
| Sao Tome and Principe | 2 | 0 (0.0%) | 0 (0.0%) | 0 (0.0%) | 0 (0.0%) | - | 1 (50.0%) | 0 (0.0%) | 1 (50.0%) | 0 (0.0%) | 0.0% |
| Saudi Arabia | 5,714 | 1,252 (21.9%) | 1,207 (21.1%) | 1,504 (26.3%) | 37 (0.6%) | 2.9% | 2,781 (48.7%) | 2,711 (47.4%) | 3,003 (52.6%) | 114 (2.0%) | 3.9% |
| Senegal | 281 | 57 (20.3%) | 37 (13.2%) | 32 (11.4%) | 4 (1.4%) | 6.6% | 156 (55.5%) | 104 (37.0%) | 99 (35.2%) | 12 (4.3%) | 7.1% |
| Serbia | 7,132 | 3,823 (53.6%) | 734 (10.3%) | 725 (10.2%) | 360 (5.0%) | 8.6% | 5,512 (77.3%) | 1,858 (26.1%) | 1,861 (26.1%) | 978 (13.7%) | 15.1% |
| Seychelles | 41 | 3 (7.3%) | 10 (24.4%) | 10 (24.4%) | 0 (0.0%) | 0.0% | 20 (48.8%) | 19 (46.3%) | 17 (41.5%) | 0 (0.0%) | 0.0% |
| Sierra Leone | 60 | 18 (30.0%) | 9 (15.0%) | 7 (11.7%) | 0 (0.0%) | 0.0% | 39 (65.0%) | 24 (40.0%) | 25 (41.7%) | 1 (1.7%) | 2.5% |
| Singapore | 998 | 436 (43.7%) | 154 (15.4%) | 160 (16.0%) | 12 (1.2%) | 2.7% | 652 (65.3%) | 389 (39.0%) | 348 (34.9%) | 58 (5.8%) | 8.2% |
| Slovakia | 8,202 | 2,842 (34.7%) | 1,653 (20.2%) | 1,320 (16.1%) | 93 (1.1%) | 3.2% | 4,846 (59.1%) | 3,740 (45.6%) | 3,235 (39.4%) | 341 (4.2%) | 6.6% |
| Slovenia | 3,247 | 1,953 (60.1%) | 303 (9.3%) | 242 (7.5%) | 63 (1.9%) | 3.1% | 2,651 (81.6%) | 723 (22.3%) | 758 (23.3%) | 254 (7.8%) | 8.7% |
| Solomon Islands | 10 | 1 (10.0%) | 0 (0.0%) | 5 (50.0%) | 0 (0.0%) | 0.0% | 2 (20.0%) | 1 (10.0%) | 8 (80.0%) | 0 (0.0%) | 0.0% |
| Somalia | 164 | 29 (17.7%) | 19 (11.6%) | 22 (13.4%) | 2 (1.2%) | 6.5% | 78 (47.6%) | 69 (42.1%) | 68 (41.5%) | 7 (4.3%) | 8.2% |
| South Africa | 4,603 | 1,507 (32.7%) | 644 (14.0%) | 514 (11.2%) | 103 (2.2%) | 6.4% | 3,017 (65.5%) | 1,775 (38.6%) | 1,451 (31.5%) | 335 (7.3%) | 10.0% |
| South Sudan | 64 | 7 (10.9%) | 8 (12.5%) | 5 (7.8%) | 0 (0.0%) | 0.0% | 37 (57.8%) | 26 (40.6%) | 29 (45.3%) | 0 (0.0%) | 0.0% |
| Spain | 56,347 | 27,667 (49.1%) | 6,210 (11.0%) | 5,383 (9.6%) | 1,142 (2.0%) | 4.0% | 43,188 (76.6%) | 16,334 (29.0%) | 15,662 (27.8%) | 4,317 (7.7%) | 9.1% |
| Sri Lanka | 726 | 177 (24.4%) | 102 (14.0%) | 148 (20.4%) | 5 (0.7%) | 2.7% | 414 (57.0%) | 277 (38.2%) | 297 (40.9%) | 28 (3.9%) | 6.3% |
| Sudan | 585 | 127 (21.7%) | 86 (14.7%) | 74 (12.6%) | 4 (0.7%) | 3.1% | 318 (54.4%) | 233 (39.8%) | 220 (37.6%) | 31 (5.3%) | 8.9% |
| Suriname | 102 | 44 (43.1%) | 15 (14.7%) | 19 (18.6%) | 1 (1.0%) | 2.2% | 79 (77.5%) | 38 (37.3%) | 36 (35.3%) | 4 (3.9%) | 4.8% |
| Sweden | 3,027 | 1,500 (49.6%) | 375 (12.4%) | 303 (10.0%) | 69 (2.3%) | 4.4% | 2,277 (75.2%) | 880 (29.1%) | 900 (29.7%) | 248 (8.2%) | 9.8% |
| Switzerland | 10,876 | 4,646 (42.7%) | 1,547 (14.2%) | 1,267 (11.6%) | 138 (1.3%) | 2.9% | 7,740 (71.2%) | 4,013 (36.9%) | 3,444 (31.7%) | 615 (5.7%) | 7.4% |
| Syrian Arab Republic | 1,358 | 244 (18.0%) | 240 (17.7%) | 307 (22.6%) | 4 (0.3%) | 1.6% | 679 (50.0%) | 659 (48.5%) | 628 (46.2%) | 25 (1.8%) | 3.6% |
| Taiwan, Province of China | 7,529 | 3,224 (42.8%) | 1,127 (15.0%) | 915 (12.2%) | 114 (1.5%) | 3.4% | 5,107 (67.8%) | 2,527 (33.6%) | 2,440 (32.4%) | 383 (5.1%) | 7.0% |
| Tajikistan | 583 | 226 (38.8%) | 107 (18.4%) | 88 (15.1%) | 1 (0.2%) | 0.4% | 403 (69.1%) | 243 (41.7%) | 257 (44.1%) | 49 (8.4%) | 10.8% |
| Tanzania, United Republic of | 893 | 178 (19.9%) | 124 (13.9%) | 135 (15.1%) | 7 (0.8%) | 3.8% | 506 (56.7%) | 325 (36.4%) | 343 (38.4%) | 39 (4.4%) | 7.2% |
| Thailand | 2,371 | 826 (34.8%) | 377 (15.9%) | 303 (12.8%) | 46 (1.9%) | 5.3% | 1,437 (60.6%) | 881 (37.2%) | 806 (34.0%) | 138 (5.8%) | 8.8% |
| Timor-Leste | 43 | 5 (11.6%) | 5 (11.6%) | 10 (23.3%) | 1 (2.3%) | 16.7% | 17 (39.5%) | 21 (48.8%) | 20 (46.5%) | 1 (2.3%) | 5.6% |
| Togo | 252 | 46 (18.3%) | 30 (11.9%) | 25 (9.9%) | 1 (0.4%) | 2.1% | 133 (52.8%) | 98 (38.9%) | 101 (40.1%) | 7 (2.8%) | 5.0% |
| Trinidad and Tobago | 2,003 | 423 (21.1%) | 145 (7.2%) | 260 (13.0%) | 12 (0.6%) | 2.8% | 1,256 (62.7%) | 640 (32.0%) | 807 (40.3%) | 50 (2.5%) | 3.8% |
| Tunisia | 3,275 | 738 (22.5%) | 616 (18.8%) | 586 (17.9%) | 13 (0.4%) | 1.7% | 1,759 (53.7%) | 1,474 (45.0%) | 1,387 (42.4%) | 71 (2.2%) | 3.9% |
| Türkiye | 10,443 | 3,128 (30.0%) | 2,098 (20.1%) | 2,160 (20.7%) | 143 (1.4%) | 4.4% | 5,780 (55.3%) | 4,647 (44.5%) | 4,604 (44.1%) | 520 (5.0%) | 8.3% |
| Turkmenistan | 304 | 83 (27.3%) | 49 (16.1%) | 44 (14.5%) | 3 (1.0%) | 3.5% | 171 (56.3%) | 125 (41.1%) | 113 (37.2%) | 7 (2.3%) | 3.9% |
| Turks and Caicos Islands | 10 | 2 (20.0%) | 0 (0.0%) | 2 (20.0%) | 0 (0.0%) | 0.0% | 6 (60.0%) | 3 (30.0%) | 5 (50.0%) | 0 (0.0%) | 0.0% |
| Uganda | 713 | 112 (15.7%) | 74 (10.4%) | 70 (9.8%) | 3 (0.4%) | 2.6% | 388 (54.4%) | 248 (34.8%) | 266 (37.3%) | 31 (4.3%) | 7.4% |
| Ukraine | 13,288 | 5,550 (41.8%) | 2,333 (17.6%) | 1,645 (12.4%) | 181 (1.4%) | 3.2% | 8,620 (64.9%) | 5,158 (38.8%) | 4,245 (31.9%) | 921 (6.9%) | 9.7% |
| United Arab Emirates | 1,904 | 705 (37.0%) | 316 (16.6%) | 271 (14.2%) | 24 (1.3%) | 3.3% | 1,227 (64.4%) | 693 (36.4%) | 675 (35.5%) | 96 (5.0%) | 7.3% |
| United Kingdom | 54,028 | 23,692 (43.9%) | 6,939 (12.8%) | 6,127 (11.3%) | 1,479 (2.7%) | 5.9% | 39,456 (73.0%) | 17,508 (32.4%) | 17,319 (32.1%) | 5,037 (9.3%) | 11.3% |
| United States | 105,233 | 40,957 (38.9%) | 14,888 (14.1%) | 13,252 (12.6%) | 2,953 (2.8%) | 6.7% | 73,932 (70.3%) | 36,775 (34.9%) | 35,837 (34.1%) | 10,000 (9.5%) | 11.9% |
| United States Minor Outlying Islands | 9 | 5 (55.6%) | 2 (22.2%) | 0 (0.0%) | 0 (0.0%) | 0.0% | 8 (88.9%) | 4 (44.4%) | 1 (11.1%) | 0 (0.0%) | 0.0% |
| Uruguay | 1,635 | 422 (25.8%) | 339 (20.7%) | 216 (13.2%) | 14 (0.9%) | 3.2% | 955 (58.4%) | 785 (48.0%) | 556 (34.0%) | 54 (3.3%) | 5.4% |
| Uzbekistan | 1,909 | 566 (29.6%) | 270 (14.1%) | 367 (19.2%) | 26 (1.4%) | 4.4% | 1,071 (56.1%) | 820 (43.0%) | 792 (41.5%) | 84 (4.4%) | 7.3% |
| Vanuatu | 16 | 9 (56.3%) | 0 (0.0%) | 2 (12.5%) | 0 (0.0%) | 0.0% | 11 (68.8%) | 5 (31.3%) | 3 (18.8%) | 0 (0.0%) | 0.0% |
| Venezuela, Bolivarian Republic of | 3,525 | 847 (24.0%) | 590 (16.7%) | 607 (17.2%) | 75 (2.1%) | 8.1% | 1,977 (56.1%) | 1,459 (41.4%) | 1,399 (39.7%) | 195 (5.5%) | 9.0% |
| Viet Nam | 10,397 | 5,350 (51.5%) | 993 (9.6%) | 1,144 (11.0%) | 94 (0.9%) | 1.7% | 8,547 (82.2%) | 2,461 (23.7%) | 2,975 (28.6%) | 520 (5.0%) | 5.7% |
| Virgin Islands, British | 2 | 1 (50.0%) | 0 (0.0%) | 0 (0.0%) | 0 (0.0%) | 0.0% | 2 (100.0%) | 0 (0.0%) | 0 (0.0%) | 0 (0.0%) | 0.0% |
| Western Sahara | 1 | 0 (0.0%) | 0 (0.0%) | 0 (0.0%) | 0 (0.0%) | - | 0 (0.0%) | 1 (100.0%) | 1 (100.0%) | 0 (0.0%) | - |
| Yemen | 3,053 | 593 (19.4%) | 671 (22.0%) | 673 (22.0%) | 22 (0.7%) | 3.6% | 1,454 (47.6%) | 1,426 (46.7%) | 1,464 (48.0%) | 74 (2.4%) | 4.8% |
| Zambia | 728 | 83 (11.4%) | 82 (11.3%) | 78 (10.7%) | 2 (0.3%) | 2.4% | 340 (46.7%) | 286 (39.3%) | 202 (27.7%) | 19 (2.6%) | 5.3% |
| Zimbabwe | 223 | 45 (20.2%) | 17 (7.6%) | 36 (16.1%) | 3 (1.3%) | 6.3% | 135 (60.5%) | 71 (31.8%) | 78 (35.0%) | 14 (6.3%) | 9.4% |
| *Total* | *1,690,849* | *794,456 (47.0%)* | *214,179 (12.7%)* | *193,980 (11.5%)* | *31,600 (1.9%)* | *3.8%* | *1,229,876 (72.7%)* | *525,046 (31.1%)* | *521,693 (30.9%)* | *136,397 (8.1%)* | *10.0%* |
