## Supplementary figures and images for "Validation of an AI for Skin Diseases in Korea and Global Usage Statistics"

### Fig S1

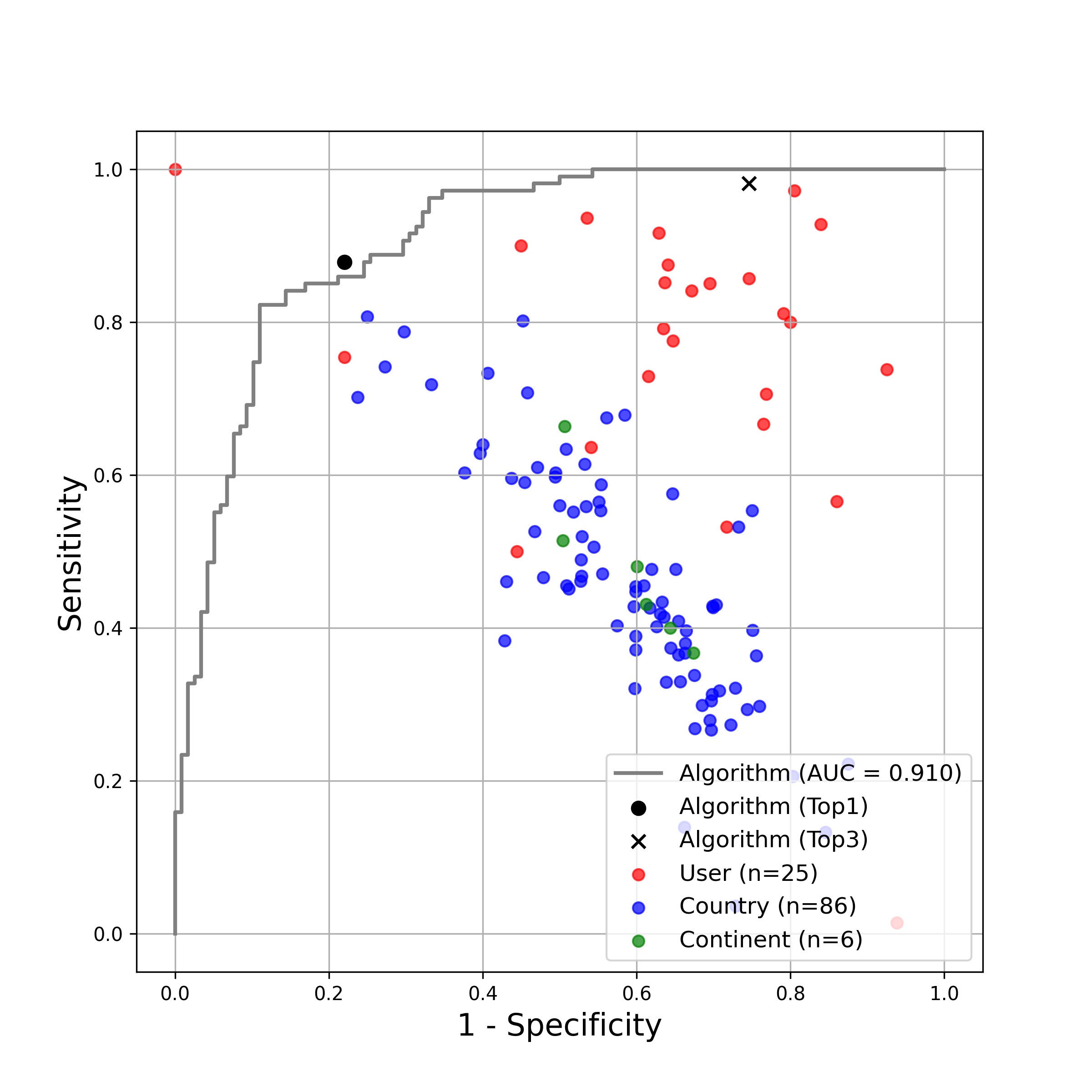

### Fig S2A

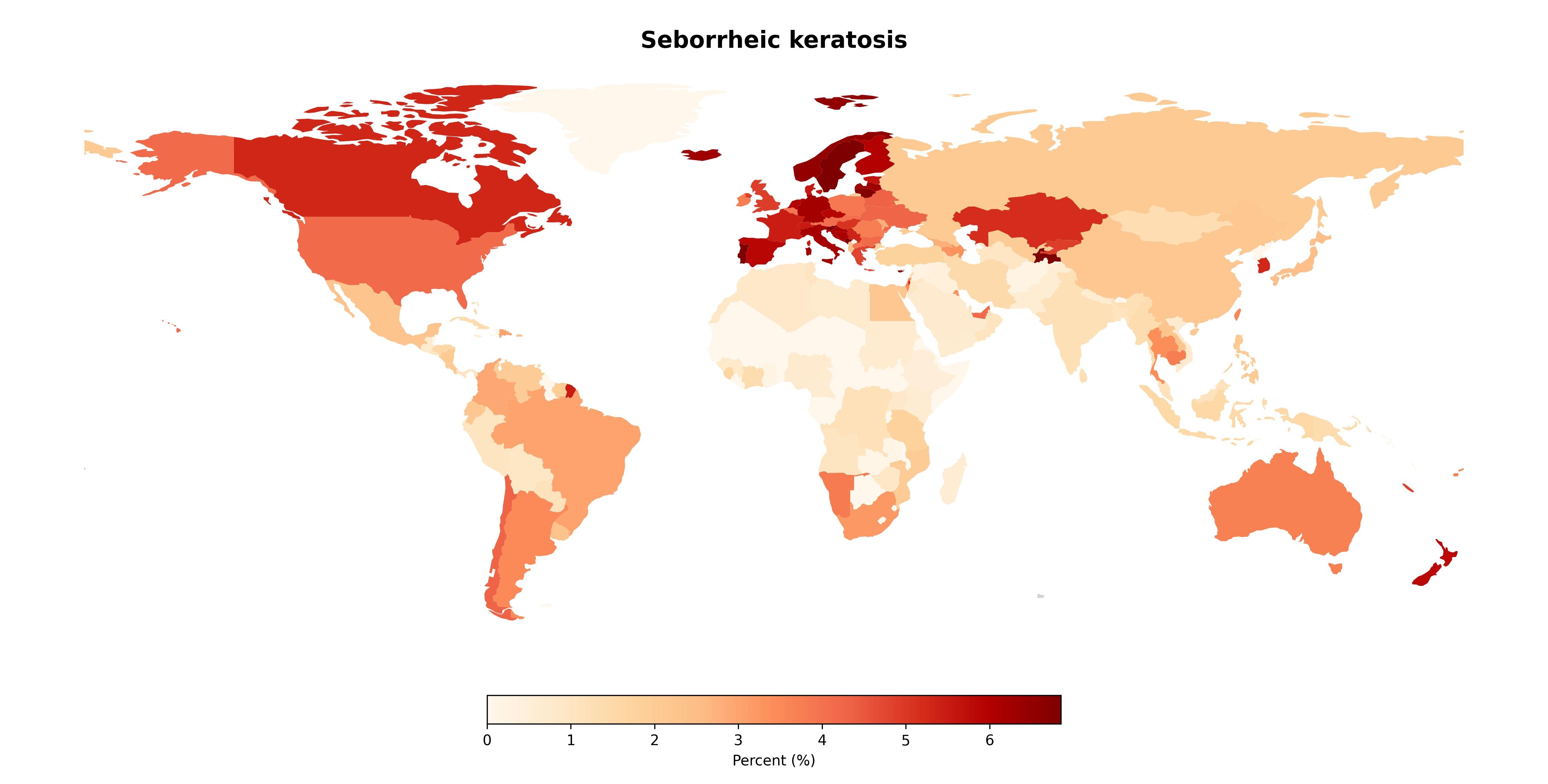

### Fig S2B

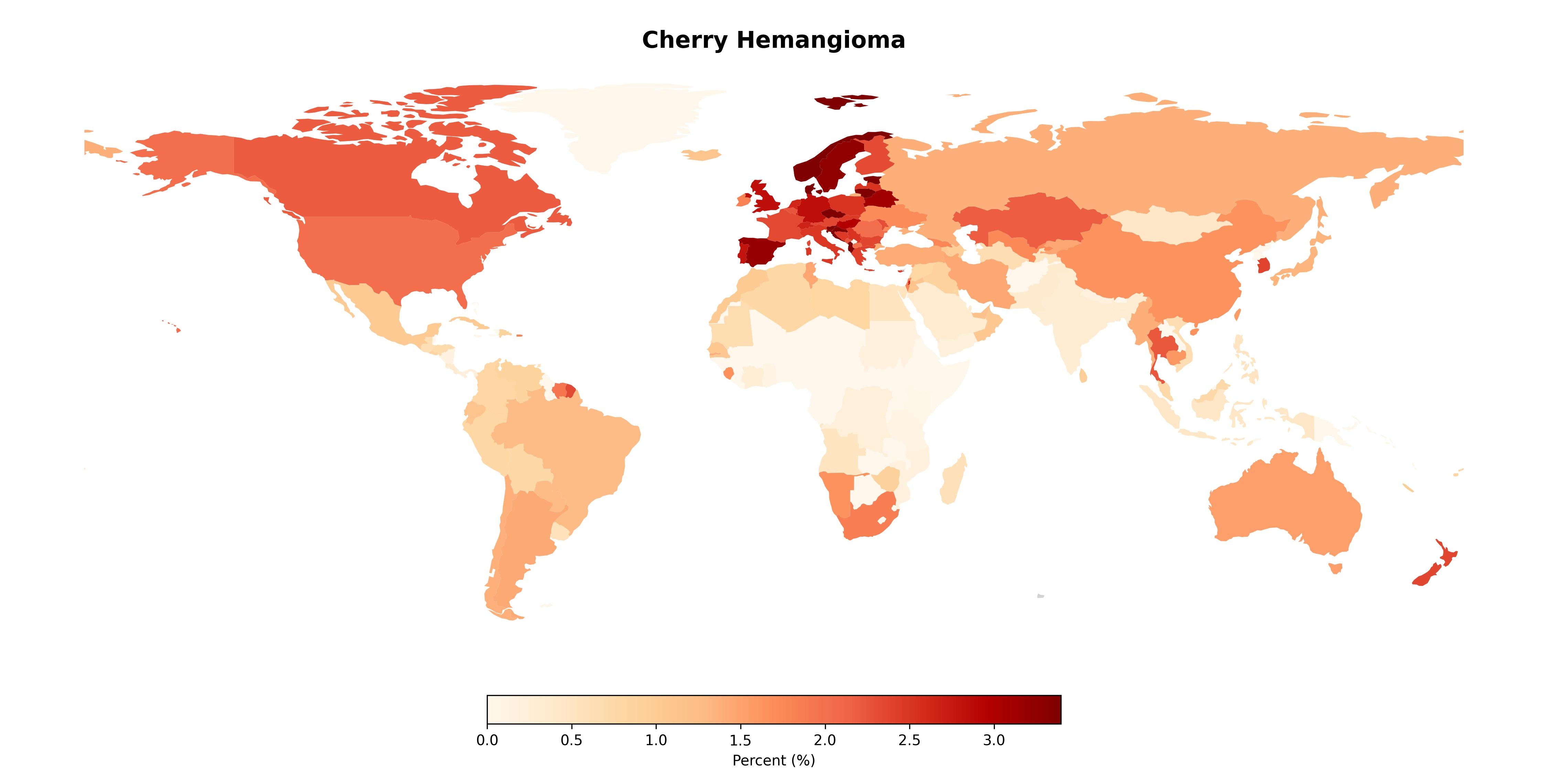

### Fig S2C

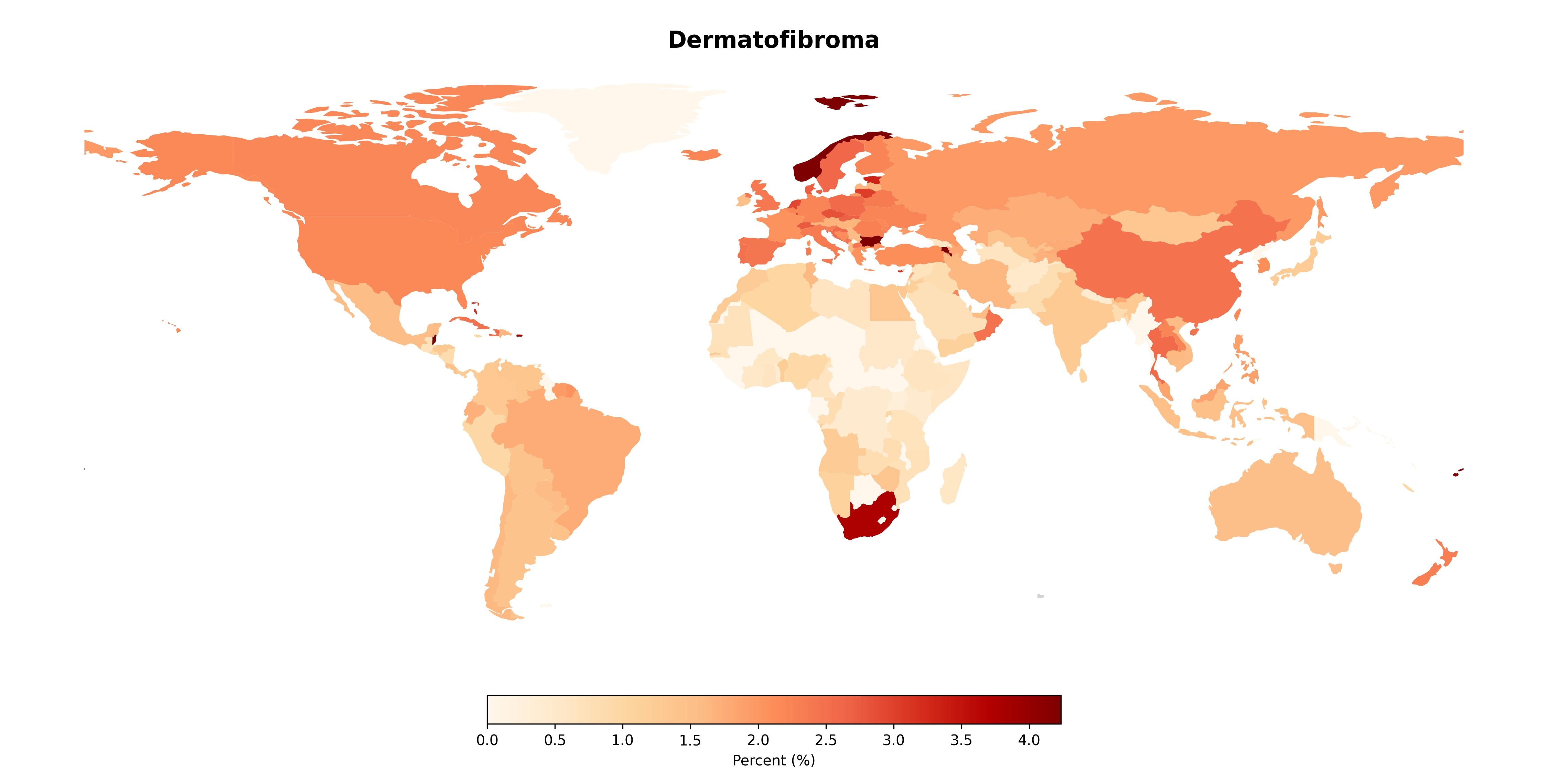

### Fig S2D

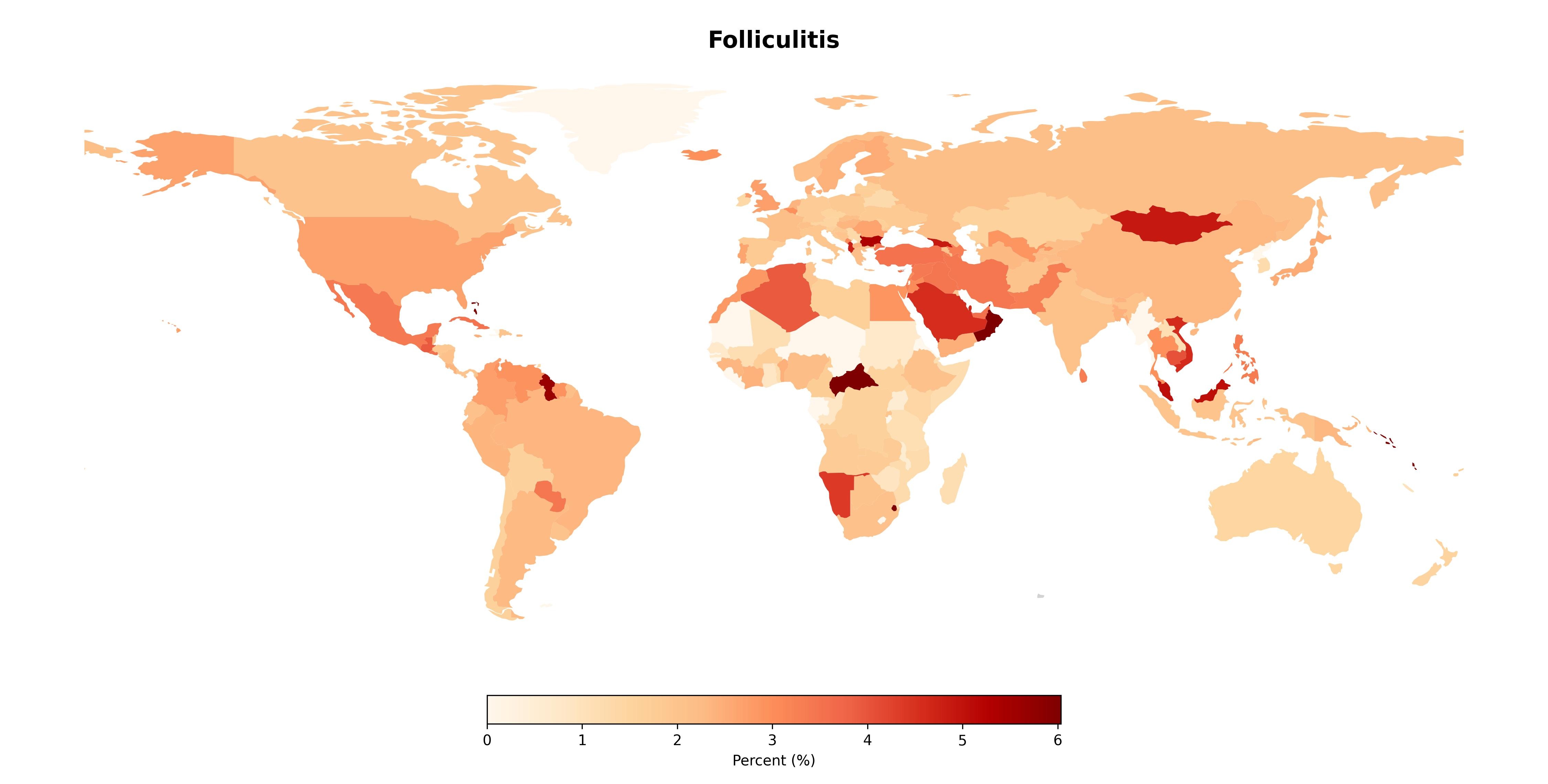

### Fig S2E

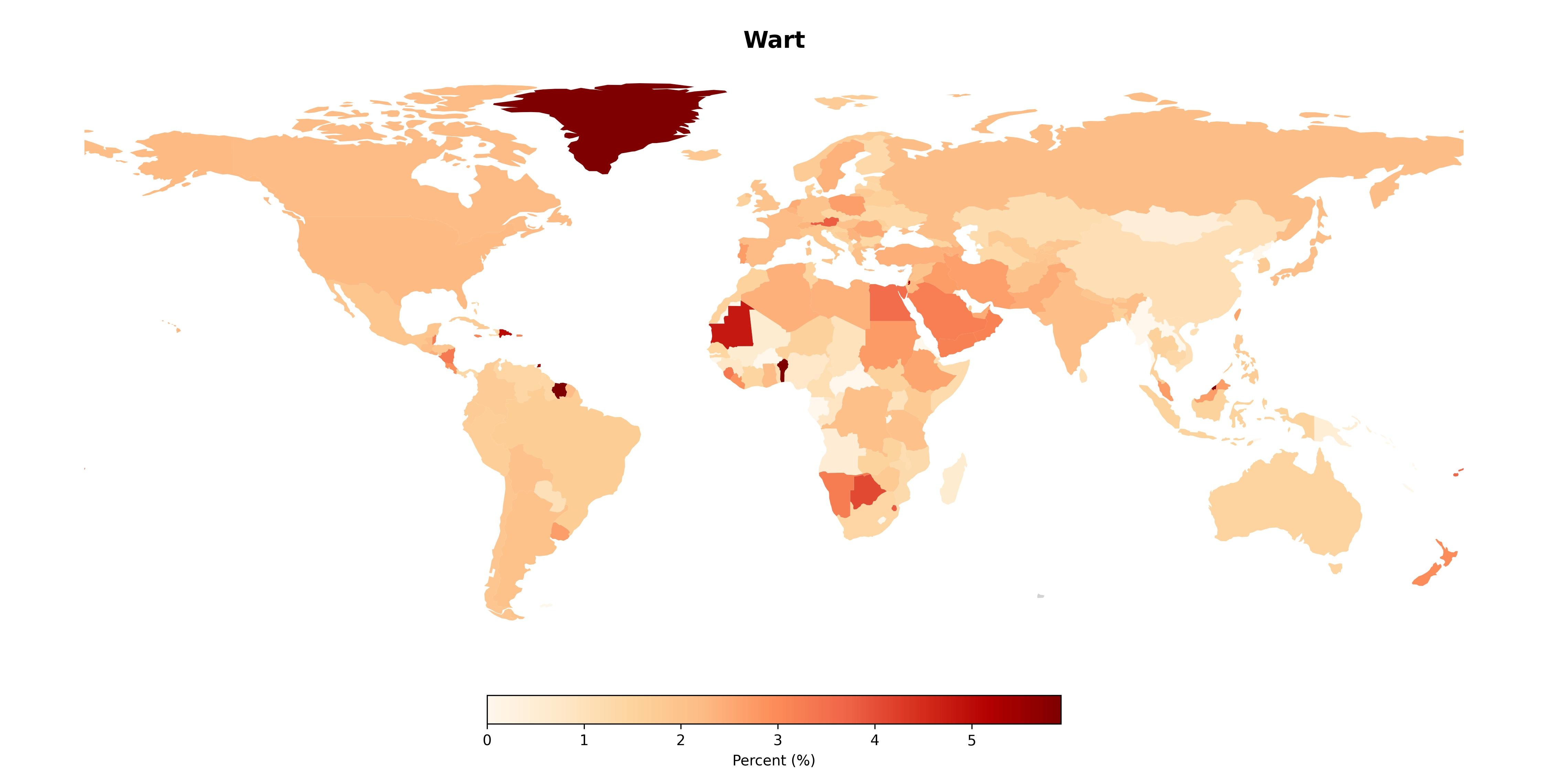

### Fig S2F

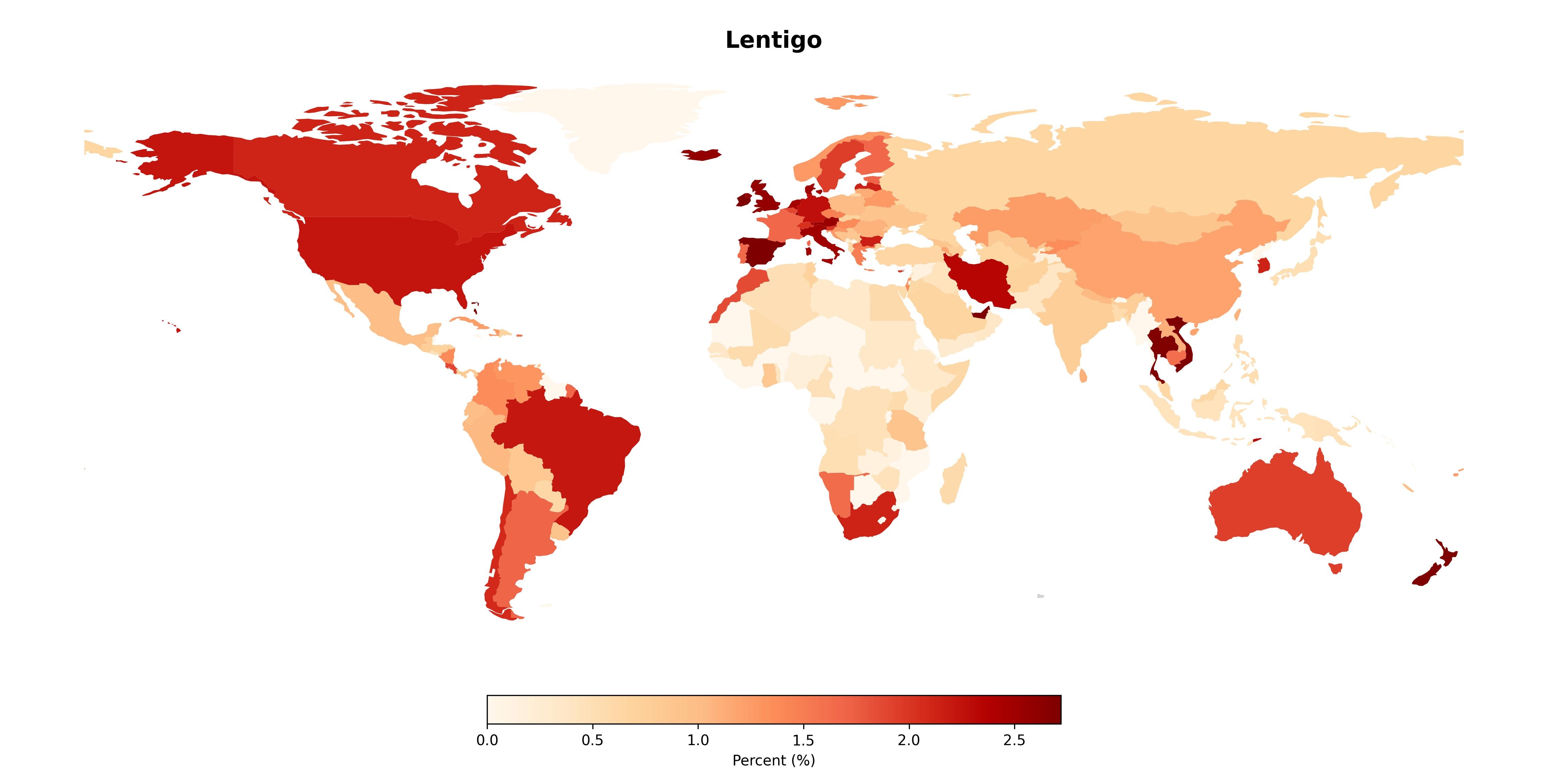

### Fig S2G

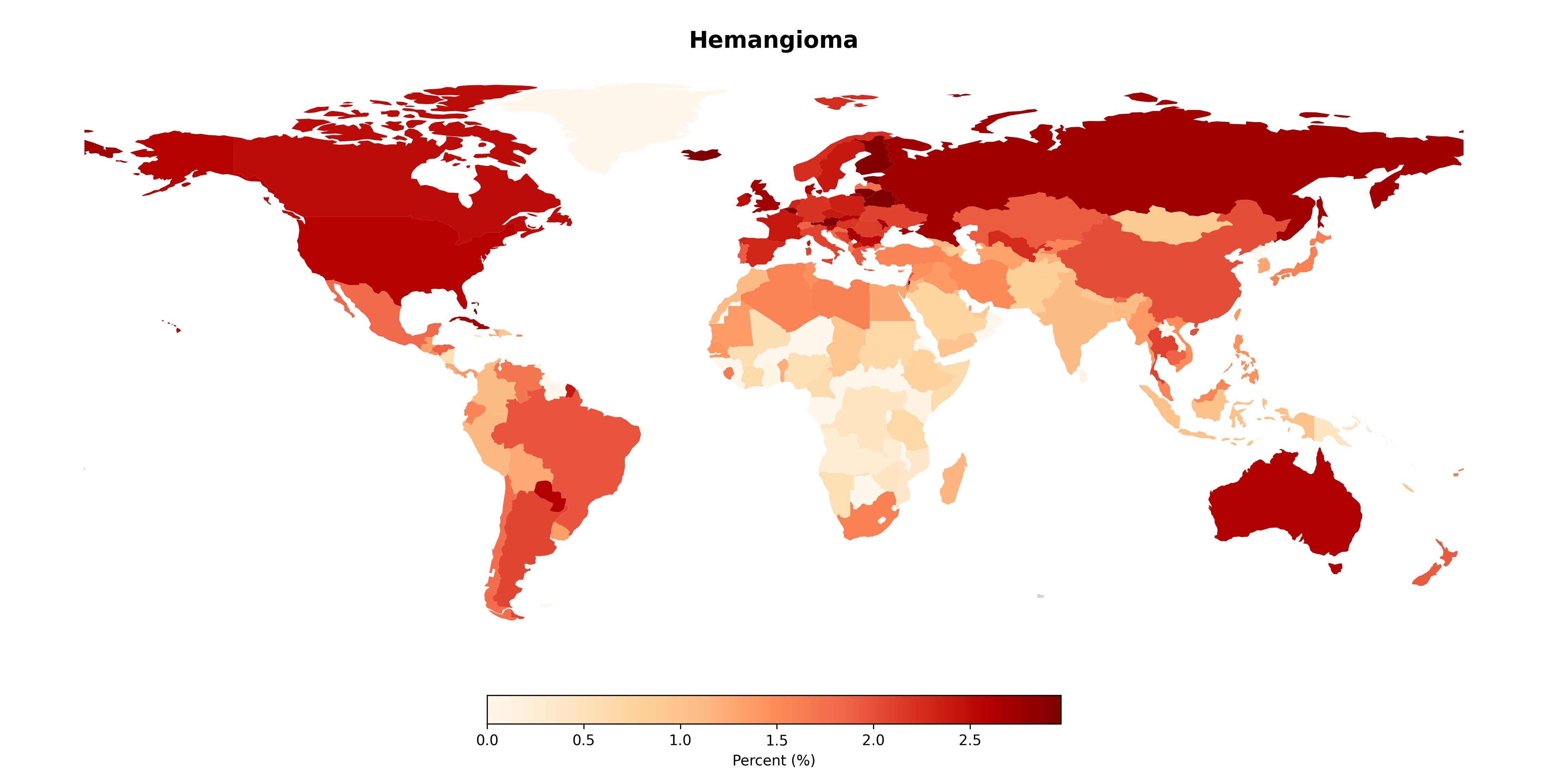

### Fig S2H

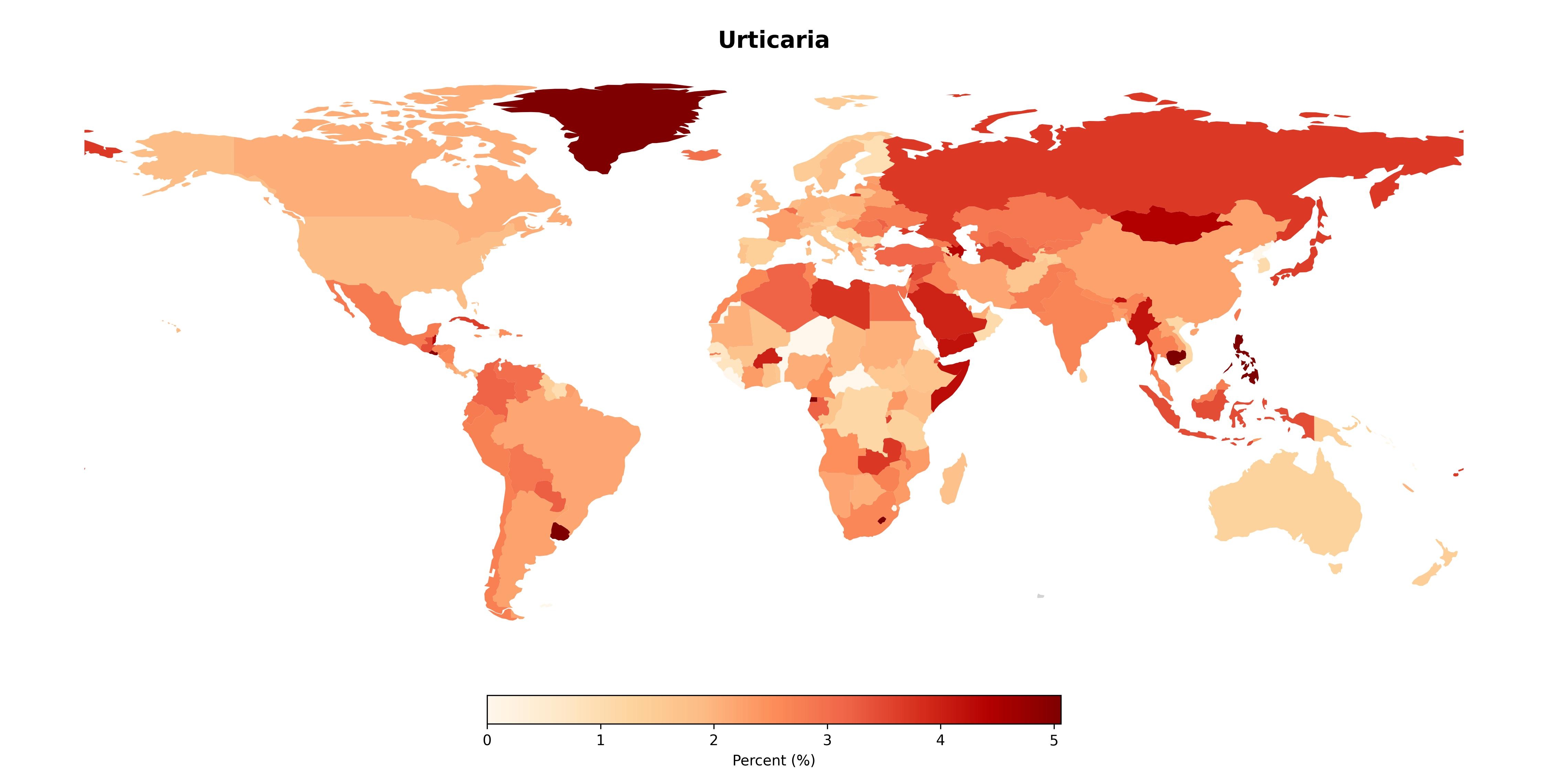

### Fig S2I

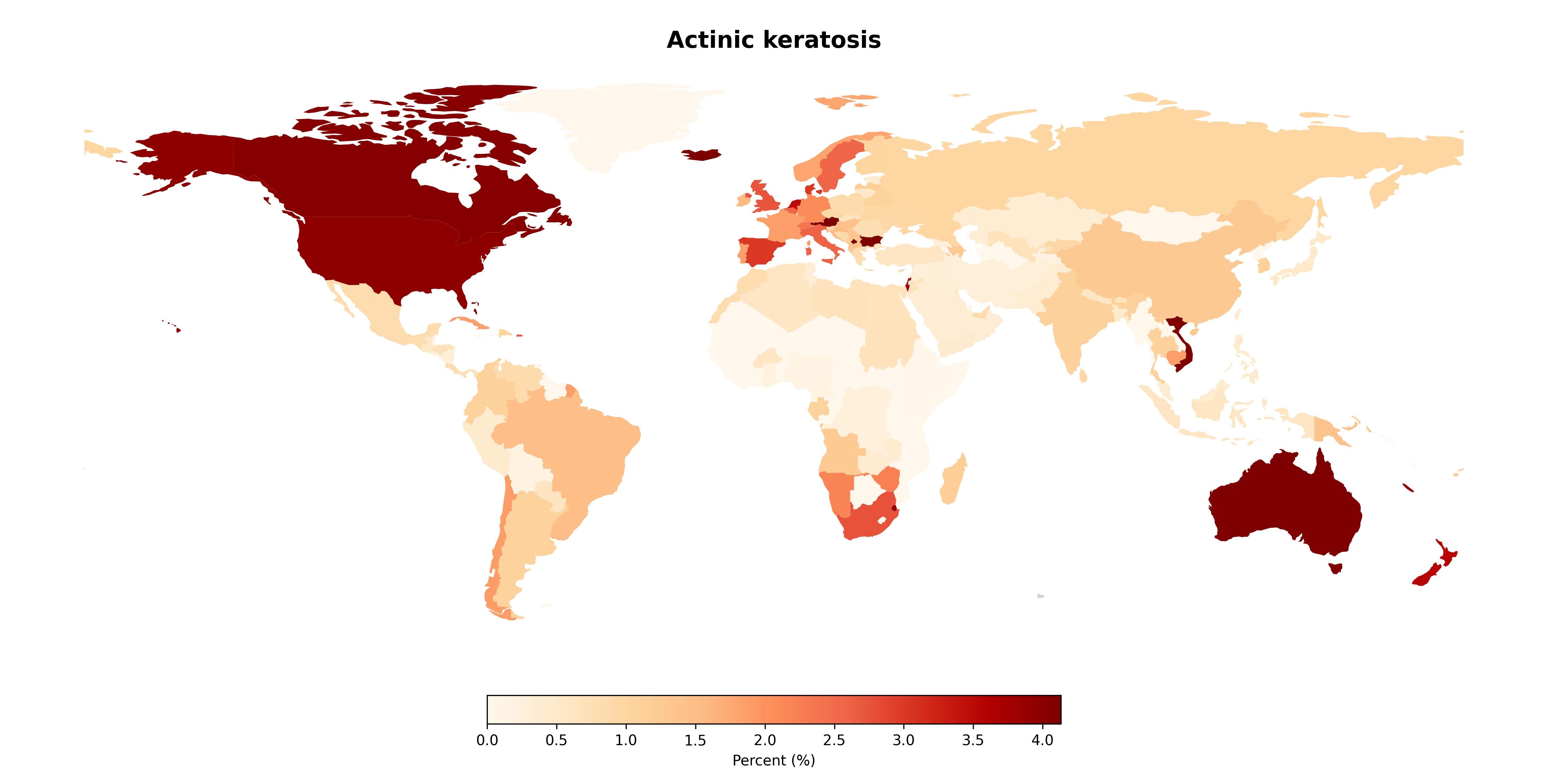
